# Effects of Dish Name, Visible Amount, Persona, and Explicit Instruction on Energy Estimates From Food Photographs by 26 Vision–Language Models: Comparative Evaluation Study

**DOI:** 10.64898/2026.10.09.26365134

**Authors:** Shinichi Nakagawa, Akira Yamamoto

## Abstract

**Background:** Vision–language models (VLMs) are being considered for estimating energy intake from meal photographs. Prompt descriptions such as a Japanese-dietitian persona shift the estimates, suggesting their use for calibration; what the estimates rest on was unknown.

**Objective:** We examined whether prompt descriptions can serve as calibration, at which training stage responses to a nationality persona and to an amount instruction arise, and whether estimates follow the visible amount or one associated with the identified dish.

**Methods:** Eleven commercial and 15 open-weight models were evaluated on NutriImage (887 Japanese single-dish photographs) and SNAPMe (1,134 US meal photographs) under eight prompt conditions (no description, five personas, and smaller- or larger-portion instructions). With one model, images were cropped or replaced by the dish name. Before-and after-meal photographs of 804 meals (ACETADA) were estimated. Pre-trained and post-trained weights of Gemma 3 and InternVL3.5 8B were compared by likelihood scoring. Prompts were compared with numerical calibration.

**Results:** Where the dish was named as in the full image, a 25% crop changed neither the median absolute percentage error (MedAPE; 33.8% vs 34.4%) nor the persona shift (−20 vs −20 kcal); dish names alone gave similar accuracy (MedAPE 31.7%); nearly empty trays still received 39%–75% of the before-meal estimate, often described as already eaten. Between models, the persona shift was related to baseline bias (r = −0.77) and less to instruction following (r = −0.36); the instruction shift showed the reverse (r = −0.14 and −0.95). In Gemma 3, the Japan–US contrast was 2–4 times larger before post-training, and the instruction response 2–4 times larger after it (InternVL3.5 8B: 3 and 1.4 times). Across images, the Japan–US contrast was uniform on US meals and, for Japanese dishes in the commercial models, Gemma 3, and Qwen3-VL 32B, larger where overestimated. In these models, the persona lowered MedAPE for Japanese dishes as much as, or slightly more than, subtracting the mean bias, and no more than dividing by a ratio; on US meals it did not; in open-weight families that responded little, only numerical calibration lowered error. The smaller-portion instruction raised MedAPE in models with little bias (eg, 30.0% to 50.2%).

**Conclusions:** In the model examined most closely, the estimates behaved as if anchored on an amount associated with the identified dish and only partly corrected by the visible amount. Prompt descriptions moved them by amounts depending on model and cuisine, not on the correction needed; a persona is therefore not proposed as a general means of calibration. In a facility, the findings suggest showing the identified dish for correction, calibrating by a ratio measured on reference meals with the deployed model, keeping amount statements out of the prompt, and not deriving intake from before– after-meal differences.

## Introduction

Estimating the nutrient content of a meal from a photograph has been studied as a way of recording what patients eat, in particular for dietary counselling in diabetes and dyslipidaemia [2–7]. Vision– language models (VLMs) now give energy estimates whose rank correlation with reference values is reasonable across public image sets, with a systematic error in absolute value that differs by model [5,6,8–12]. Photographs sent by patients, however, often show the table, tableware, hands, or surroundings, and many medical institutions are not in a position to send them to an external service. For such institutions, an estimate that is produced within the facility and gives an approximate energy value would be of use; the same framework might later be extended to fat and salt, but only energy is considered here.

The models that can be run within a facility are small open-weight models, and their behaviour need not be that of the commercial models on which most reports are based. Before a role description or an instruction about amount is written into an operational prompt, it is necessary to know at which stage of the estimation it acts and on what its effect depends, because this determines how such a system has to be calibrated locally.

The present study is the third of a series. In the first [13], with three models of one vendor on three image sets, the main source of error in estimated energy was the amount and not the composition of the meal, and we expressed the expectation that an object of known size in the photograph would reduce this error. In the second [1], giving six commercial VLMs the persona of a registered dietitian in Japan lowered their energy estimates in one direction on two image sets, whereas the persona of a dietitian in the United States did not; the energy composition of the estimates was unchanged; the lowering was reported to be larger for images that the model overestimated and close to zero for those it underestimated; and explicit instructions to assume a smaller or a larger portion, used as a manipulation check, moved the estimates by a nearly constant amount. Words in the prompt, that is, moved the estimate of amount.

That study left four points open, in its discussion and limitations: whether the persona acts on perception or on inference could not be distinguished; a calibration introduced by post-training could not be excluded as the origin of the response; whether the models use a reference for scale in the photograph was not examined; and the prior itself, the amount that a model associates with a dish, was not measured. The first report had observed that Claude Sonnet and Claude Opus appeared to identify the type of food and to apply nutritional knowledge to it, and had discussed amount as the main source of error. From this observation it was suggested, and in the second study stated as an interpretation, that estimation proceeds in two stages, identification of the dish followed by retrieval of an amount from knowledge (hereafter H1, the two-stage hypothesis), and that the persona replaces the serving that is assumed at the second stage (H2, the replacement hypothesis). Neither was tested.

Related observations exist in neighbouring areas. In image-based nutrient estimation, the estimation of amount has repeatedly been found to be the weak link [10,14–18]. VLMs are known to answer from prior knowledge in preference to visual evidence in some tasks [19–23]. In the evaluation that introduced the ACETADA image set, supplying context as text lowered the error of several models [11]; and in a study that proposed a model trained on paired before- and after-meal images, a commercial VLM asked for the weight consumed showed a percentage error of about 39%, and text input alone gave a lower error than image input alone [24]. An ‘expert’ persona (a nutrition expert or registered dietitian, without nationality) has been evaluated as one of several prompting techniques for reducing error, with results that differ between reports [6,11]. Nationality and cultural personas shift the opinions and values expressed by language models [25–29], and in human observers judgements of amount depend on a reference portion [30,31].

Three questions were asked. (Q1) If descriptions in the prompt move the estimates systematically, can they be used for calibration? (Q2) At which stage of training does the response to a nationality persona arise, and is it the same response as that to an explicit instruction about amount? (Q3) Do the models read the amount in the photograph, or do they identify the dish and take an amount from what they know of it? In addition, we examined whether the size of the persona shift depends on model family, model size, numerical precision, or vision encoder, and what the shifts imply for accuracy.

Predictions were derived from H1 and H2 (Table 1). Some were tested on data already in hand, and others by experiments for which the expected outcome was written down before the run; Table 1 distinguishes the two, and marks for each prediction whether it replicates the previous study, answers a point that study left open, or corrects it. What was already reported there (that the lowering is in one direction, that the nationality in the description carries it, that its effect on accuracy depends on the sign of baseline bias, and that a persona is not recommended in general as a means of calibration) is reported here briefly, as a replication in 26 models.

**Table 1.** Predictions and how they were tested.

|  | Prediction | Test | Outcome | Stated before the data were examined | Relation to the previous study |
| --- | --- | --- | --- | --- | --- |
| P1a | If estimation proceeds by identifying the dish and then retrieving an amount (two-stage hypothesis), then, while identification is retained, the estimate and the persona shift do not depend on how much of the image is visible. | Crop × identification judgment; dish name without image (Sections 3.1 and 3.2) | Consistent. Where the dish was named as in the full image, accuracy and the shift were unchanged at the 25% crop; the dish name alone gave similar accuracy and most of the shift (−13 vs −17 kcal). | Name-only: yes.<br>Crop: no<br>(existing outputs) | Answer to an open point (perception or reasoning; the prior, measured without the image) |
| P1b | Under the same hypothesis, the same dish photographed at two amounts receives similar estimates. | Same-dish size pairs; before/after-meal pairs (Section 3.1) | Partly consistent. The estimates were not the same, against the prediction taken strictly, but they differed by much less than the amounts did: visible amount was read as a direction, reliably by two of five models, but its size was compressed (nearly empty trays, 39–75% of the before-meal estimate in the chat format); the after-meal photographs were, however, often described as a meal that had been eaten, so that this is not a clean test of how amount is read. | Before/after pairs: yes. Size pairs: no (existing outputs) | Answer to an open point (use of a reference for scale); test of the expectation of the first report |
| P1c | The persona does not change identification. | Retention under cropping by persona; names with and without the persona (none-jp, none-us); human rating of 100 pairs (Section 3.2) | Largely consistent. Retention and the correctness of the name did not differ by persona (65% vs 62%). Images whose name under the persona was judged different by the model judge showed a larger shift; by human rating most of these differences were of wording. | None-us naming control: yes. Retention and naming under jp: no (existing outputs) | Answer to an open point (perception or reasoning) |
| P2 | If the persona replaces the assumed serving (replacement hypothesis), the persona shift depends on baseline bias and not on instruction following, and the instruction shift shows the reverse. | Cell-level regressions (Section 3.3) | Consistent in group A; not identifiable in group B. | No (formulated on existing data) | Correction: in the six models of the previous study, baseline bias and instruction following varied together and could not be separated |
| P3 | If the persona channel is installed by post-training, the JP – US contrast is absent in pre-trained weights and present in instruction-tuned weights (the expectation we favoured). | Gemma 3 pt/it, likelihood scoring, NutriImage (Section 3.4) | Not supported; the contrast was larger in pt. | Yes | Answer to an open point (origin in post-training) |
| P4 | If the contrast does not depend on knowledge of particular dishes, it remains in pt on an image set without Japanese dishes. | Gemma 3 pt/it, SNAPMe (Section 3.4) | Contrast remained, at 64–82% of its NutriImage size; this lay between the two outcomes written beforehand. | Yes | Answer to an open point (origin in post-training) |
| P5 | If the size of the persona shift is not a matter of capacity, the slope of the shift against baseline bias does not change with model size, numerical precision, or vision encoder. | Size ladders, FP8, InternVL3.5 (Section 3.6) | Consistent; Qwen3-VL 32B intermediate. | No (formulated on existing data) | New question (not addressed in ref 1) |
| P6 | If the origin observed in Gemma 3 is not specific to that family, then in InternVL3.5 8B the JP – US contrast is negative in the pre-trained weights, smaller in absolute value in the final weights, and larger – smaller is larger in the final weights. | InternVL3.5 8B pre-trained and final weights, likelihood scoring, NutriImage (Section 3.4) | Consistent in sign and ordering (–57 vs –19 kcal; +44 vs +62 kcal). The sizes written beforehand were missed: the pre-trained contrast was larger than the –10 to –40 kcal expected, and the ratio of instruction responses (1.4) was below the 1.5 expected. | Yes | Replication of the Gemma 3 finding in a second family |

The predictions were derived from the two hypotheses (H1, H2) of the previous study [1]. The forms of the contrasts across images (Section 3.5) and the comparison with numerical calibration (Section 3.7) are reported as description and not as predictions.

To our knowledge, how far the estimate of a general-purpose VLM follows the identified dish as against the visible amount has not been examined by manipulating the two separately, nor has it been examined how descriptions in the prompt enter such an estimate. In our previous study, with six models, baseline bias and instruction following varied together across vendors, so that their roles could not be separated; a larger set of models was needed for this (across its 12 cells, baseline bias and larger − smaller correlated at r = 0.60; across the 48 cells here, at 0.20). The aim is a description at the level of behaviour. Internal representations were not examined, and no new method for improving accuracy is proposed.

## Methods

### 2.1 Image sets and reference values

#### NutriImage

NutriImage consists of photographs of single dishes served at the Kyoto University co-op cafeteria, taken in 2002–2005 for a project on health and nutrition information. The reference values were calculated by registered dietitians from the ingredient-level recipe and the total served weight of each dish, with the Standard Tables of Food Composition in Japan (fifth revised edition, 2000). The photographs show prepared food only, and no person appears in them. The data set is maintained by the authors and is archived on Zenodo (concept doi:10.5281/zenodo.21944881). The version used here (v1.1; doi:10.5281/zenodo.21961032) is that of our previous study [1] and differs from the set of our earlier report [13] in three respects: text burned into the photographs was removed by cropping each image to the bounding box of the dish as detected by GPT-4o, instead of by filling a mask; the reference values were rebuilt from the recipe tables; and the evaluation set comprises 899 dish images, of which the 887 with linked reference values were analysed. As a consequence of the cropping, the images differ in pixel size (JPEG; width 161–640 and height 121–480 pixels, median 403 × 303, 0.03–0.31 megapixels; the two most common sizes, 403 × 303 and 640 × 480, account for 182 and 97 images), the dish fills most of the frame in most images, and no object of known size is visible (Figure 1).

**Figure 1.**
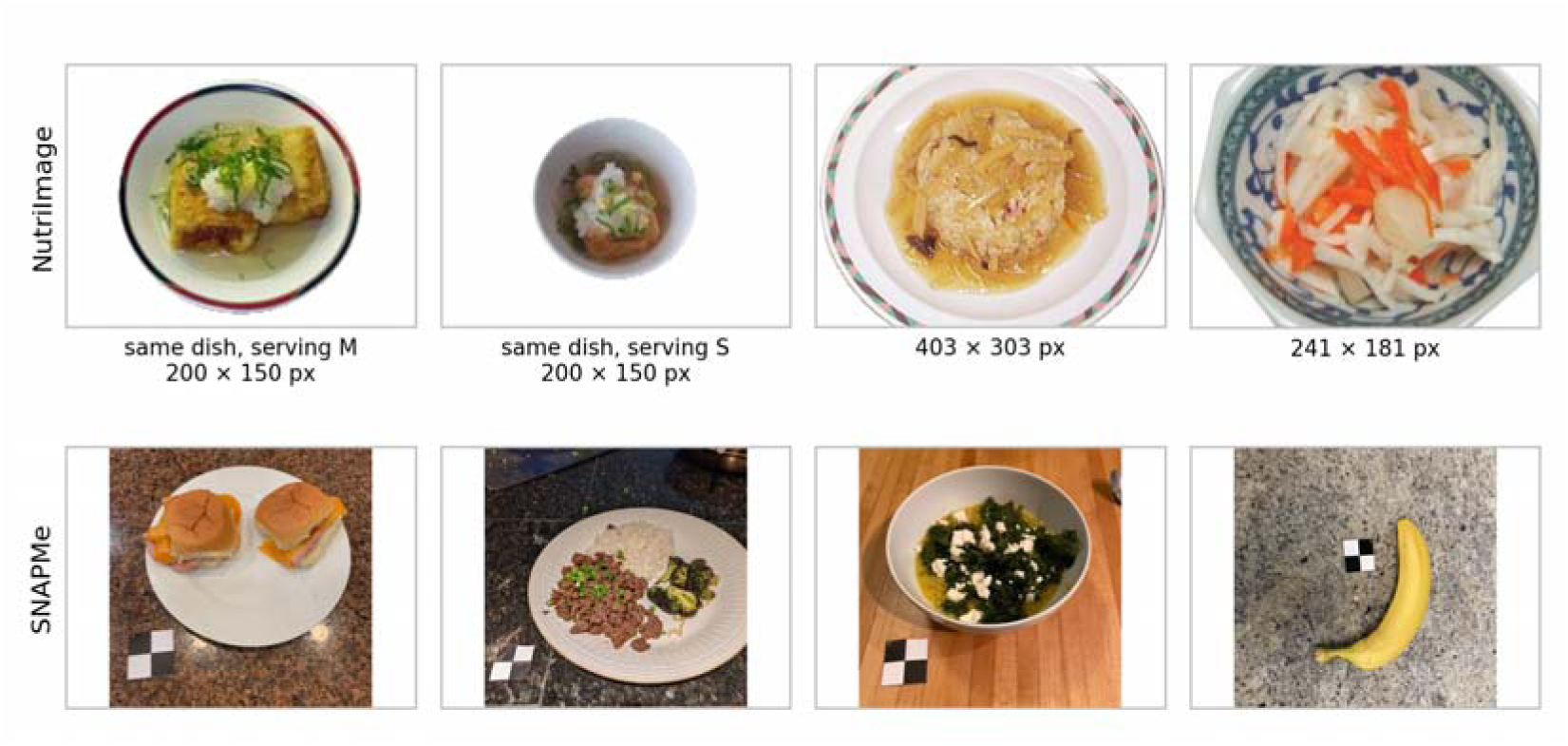
Example images of the two image sets. Top, NutriImage: single dishes cropped to the detected bounding box, with pixel sizes; the two images on the left are one of the same-dish pairs served at two sizes (pair 1). Middle, SNAPMe: meals photographed by free-living participants with a marker of known size. NutriImage photographs are the authors’ own (v1.1, doi:10.5281/zenodo.21961032); SNAPMe images are from the public dataset on Ag Data Commons [32].

#### SNAPMe

SNAPMe [32] consists of photographs of meals taken by free-living adults in the United States with their own phones, each with a marker of known size, and reference values derived from food records that the participants entered on the same day (ASA24). Of the 1,272 ‘before’ images of the previous study, the 1,134 whose native resolution was exactly 800 × 800 pixels (JPEG) were used; the remaining 138 higher-resolution images were excluded to keep input resolution uniform without any resizing. Reference energy distributions of retained and excluded images were comparable (mean 313 vs 319 kcal). Reference energy was the per-image sum of all recorded items, including beverages (images at or below 20 kcal are excluded from the image-level analyses; Section 2.5).

#### Reference tables

Reference energy was taken from one per-image table for each set and used for all models, including the re-aggregation of the 2025 commercial results of the previous study. The numbers of images analysed per cell (Table 2) are slightly below the set sizes where a response could not be parsed or was truncated; no image was excluded for its content. The image sets used only in the analysis of visible amount are described in Section 2.3(e).

#### ACETADA

ACETADA [11] consists of smartphone photographs taken before and after eating in a randomised crossover feeding study in Perth, Western Australia, in which 152 adults were provided with breakfast, lunch, and dinner over three days; every photograph includes a fiducial marker for scale. It was used only in the analysis of visible amount; the pairs analysed and the reference values are described in Section 2.3(e).

### 2.2 Models

Table 3 lists the models. The 2026 commercial models were Claude Haiku 4.5 and Claude Sonnet 5 (Anthropic), GPT-5.6 Sol and GPT-5.6 Luna (OpenAI), and Gemini 3.8 Flash and Gemini 3.1 Pro (Google), accessed through the vendors’ interfaces in September 2026 and chosen as the successors of the six 2025 models of the previous study (Claude Haiku 4.5, Claude Sonnet 4.6, GPT-4o, GPT-4o-mini, Gemini 2.5 Flash, Gemini 2.5 Pro), whose outputs were re-aggregated as a separate group.

**Table 3.** Models and execution settings.

| Group | Model (identifier) | Weights | Runtime | Temperature; thinking; output limit | Run date (2026) |
| --- | --- | --- | --- | --- | --- |
| Commercial 2026 | Claude Haiku 4.5 (claude-haiku-4-5-20251001); Claude Sonnet 5 (claude-sonnet-5) | — | Vendor API | default; disabled; 256 | 09-13 to 09-14 |
|  | GPT-5.6 Sol (gpt-5.6-sol); GPT-5.6 Luna (gpt-5.6-luna) | — | Vendor API | default; reasoning effort none; 256 | 09-13 to 09-14 |
|  | Gemini 3.8 Flash (gemini-3.8-flash); Gemini 3.1 Pro (gemini-3.1-pro-preview) | — | Vendor API | default; budget 0 (Flash), level low (Pro); 2,048 | 09-13 to 09-14 |
| Commercial 2025 (previous study) | Claude Haiku 4.5; Claude Sonnet 4.6; GPT-4o; GPT-4o-mini; Gemini 2.5 Flash; Gemini 2.5 Pro (claude-haiku-4-5-20251001, claude-sonnet-4-6, gpt-4o, gpt-4o-mini, gemini-2.5-flash, gemini-2.5-pro) | — | Vendor API | default; —; 500 | 2026: SN 06-28, NI 07-04 to 07-05; Flash NI re-run 08-15 to 08-16; added conditions 08-15 to 08-19 |
| Open, Qwen3-VL | Qwen/Qwen3-VL-{2B,4B,8B}-Instruct | BF16 | vLLM 0.11, 1 × H100 | 0; —; 256 | 09-08 |
|  | Qwen/Qwen3-VL-32B-Instruct; Qwen/Qwen3-VL-30B-A3B-Instruct (MoE) | BF16 | vLLM 0.11, 8 × H100 | 0; —; 256 | 09-12 |
|  | Qwen/Qwen3-VL-{2B,4B,8B,32B}-Instruct-FP8; Qwen/Qwen3-VL-30B-A3B-Instruct-FP8 (official FP8 weights) | FP8 | vLLM 0.11, H100 | 0; —; 256 | 09-12 (32B: 09-08) |
| Open, Gemma 3 | unsloth/gemma-3-{4b,12b,27b}-it | BF16 | vLLM 0.11, 1 × H100 | 0; —; 256 | 09-12 |
| Open, InternVL3.5 | OpenGVLab/InternVL3_5-{2B,4B,8B,14B}-HF; -30B-A3B-HF (MoE) | BF16 | vLLM 0.11, 1 × H100 | 0; —; 256 | 09-13 |
| Open, Mistral | mistral-community/pixtral-12b; unsloth/Mistral-Small-3.2-24B-Instruct-2506 | BF16 | vLLM 0.11, 1 × H100 | 0; —; 256 | 09-13 |
| Scoring, 2.3(d) | unsloth/gemma-3-{4b,12b,27b}-{pt,it} | BF16 | Transformers 4.57, 1 × H100 | no generation | 09-15 (NI), 09-16 (SN) |
| Scoring, 2.3(d) | OpenGVLab/InternVL3_5-8B-Pretrained; OpenGVLab/InternVL3_5-8B (repository format) | BF16 | Transformers 4.57, 1 × H100 | no generation | 09-21 (NI) |
| Crop and name-only, 2.3(b, c) | Claude Sonnet 4.6; judge, Claude Haiku 4.5 | — | Vendor API | default (not specified); —; 400 | 09-07 to 09-08; name-only 09-19 |
| ACETADA, 2.3(e) | Claude Sonnet 4.6; Qwen3-VL 8B; Gemma 3 27B it (chat); Gemma 3 27B pt, it (scoring) | BF16 (open) | API; vLLM; Transformers | as above | 09-19 to 09-20 |
| 4-bit, S3 | mlx-community/Qwen3-VL-{2B,4B,8B}-Instruct-4bit | MLX 4-bit | mlx-vlm 0.6.17, Apple M4 | 0; —; not reached (longest response | 09-11 to 09-15 |

| Group | Model (identifier) | Weights | Runtime | Temperature;<br>thinking; output<br>limit | Run date (2026) |
| --- | --- | --- | --- | --- | --- |
|  |  |  |  | about 100 tokens) |  |

System placement of the description: system role for Qwen3-VL, InternVL3.5, Mistral Small 3.2, and the commercial models; beginning of the user turn for Gemma 3 and Pixtral-12B. All identifiers of the open-weight chat runs (BF16 and FP8) were read from the model field of the output files.

Claude Haiku 4.5, with the same identifier, belongs to both groups; there are thus 11 distinct commercial models and 26 models in all, and the two runs of Claude Haiku 4.5 are treated as a repeated measurement.

The open-weight models were Qwen3-VL Instruct 2B, 4B, 8B, 32B, and 30B-A3B; Gemma 3 4B, 12B, and 27B (instruction-tuned); InternVL3.5 2B, 4B, 8B, 14B, and 30B-A3B; and Pixtral-12B and Mistral Small 3.2 24B [33–39]. InternVL3.5 uses the Qwen3 language models with a different vision encoder (InternViT) and tiling scheme, which allows the language side and the vision side to be distinguished in the comparison with Qwen3-VL. The two 30B-A3B models are mixture-of-experts models with about 3B active parameters and were analysed separately (Multimedia Appendix 1, S7) [52]. Sizes were compared only within a family. For the comparison of training stages, the pre-trained and instruction-tuned weights of Gemma 3 4B, 12B, and 27B were used (Section 2.3(d)); the Gemma 3 weights were obtained from an ungated mirror (unsloth) described as an unmodified copy of the official release (Section 4.6).

### 2.3 Experimental formats

#### (a) Chat format, eight conditions

The prompt asked the model to estimate the nutritional content of the food shown from its visual appearance and to return a JSON object with energy, protein, fat, carbohydrate, salt, and a one-line basis (Multimedia Appendix 2; identical to Appendix A of the previous study [1]). Eight conditions were formed by a description placed before the prompt. In *none* there was no description. In *jp* and *us* the model was described as a Japanese (kanri-eiyoshi) or a United States registered dietitian ‘deeply familiar with standard single-serving portion sizes’ of everyday Japanese or American meals. In *jp_home* and *us_home* it was described as a Japanese or an American homemaker familiar with the usual serving sizes of home cooking, without formal training in nutrition. In *neutral_dietitian* it was described as a registered dietitian with no national or cultural specialisation; this is the only description that does not mention serving or portion sizes. In *smaller* and *larger* there was no persona, and the model was described as tending to assume smaller or larger single-serving portion sizes than usual. The *jp* condition is the primary persona condition, and *smaller* and *larger* are the explicit portion instructions. The description was given in the system role for Qwen3-VL, InternVL3.5, Mistral Small 3.2, and the commercial models, and at the beginning of the user turn for Gemma 3 and Pixtral-12B, whose templates have no system role. All models in Table 3 were run in this format on both image sets.

#### (b) Central crop and identification judgment

Claude Sonnet 4.6 was given the NutriImage images in full and cropped about the centre to 60%, 40%, and 25% of each side (36%, 16%, and 6.25% of the area), without enlargement. The original images had a median size of 403 × 303 pixels, so the 25% crop was typically 101 × 76 pixels (range 41 × 31 to 160 × 120). The conditions were *none*, *jp*, and *us*, each with and without a statement that the image shows one serving. The persona texts of this experiment were given as system prompts and differed from those of format (a): they describe a Japanese registered dietitian (kanri-eiyoshi) with extensive knowledge of Japanese cuisine, ingredients, and cooking methods, and an American registered dietitian with expertise in Western cuisine and nutrition; neither mentions portions or amounts (Multimedia Appendix 2). The model returned a dish name together with the estimate; under *jp* the name was returned in Japanese in about 99% of responses. For each image and crop, the dish name was compared with the name returned for the full image in the same condition by a second model (Claude Haiku 4.5), which answered SAME or DIFF. The criterion used in the main text required agreement of the main ingredient and the cooking method; a looser criterion requiring the main ingredient only is reported in Multimedia Appendix 1 (S10). The same criterion was used within the *none*, *jp*, and *us* conditions. Temperature was not specified (vendor default), the output limit was 400 tokens, and each request was made once. *Identification retention* is the proportion of images judged SAME. In a further analysis of the same outputs, the name returned under *jp* was compared with that returned without a persona for the same image (full size and 25% crop), with the same strict criterion and a statement that one of the two names may be in Japanese. The same comparison was made between none and *us*, as a control for the variation between two requests at the default temperature; the expected outcome of this control was written down before it was run. For 100 images drawn at random, one author (S.N.), who reads both languages and had the original menu name of each dish, rated whether the name without a persona and the name under *jp* each matched the menu, and whether the two names denoted the same dish, without sight of the judge’s output; since the name under *jp* was in Japanese, the rater was not blind to the condition. For the use of the judge within one language, 100 pairs of the names returned for the full image and for the 25% crop without a persona, drawn at random, were rated by the same author as denoting the same dish or not, without sight of the judge’s output; the criterion given to the judge was provided to the rater, and the original menu name of each dish was in view. Agreement was 84% (κ = 0.57); the rater found the two names the same in 30 pairs and the judge in 18, and in 14 of the 16 disagreements the rater had found them the same and the judge different (Multimedia Appendix 1, S10). The judge was thus the stricter of the two, and the group of images judged SAME is defined conservatively.

#### (c) Dish name without image

The same model was given only the English dish name of the NutriImage reference record, with no image, in the *none*, *jp*, and *us* conditions, using the same system prompts as in (b). Some of the English names were produced by machine translation; a secondary analysis was restricted to the images judged SAME at the 60% crop in (b).

#### (d) Likelihood scoring of pre-trained and instruction-tuned weights

Pre-trained weights do not follow a chat template, so Gemma 3 pt and it were compared in a format that uses neither a template nor generation [53–55]. The input was a short document consisting of one sentence describing the narrator, the image, and the text ‘Energy (kcal):’. Thirty candidate continuations, 50 to 1,500 kcal in steps of 50, were scored by the sum of the log-likelihoods of their tokens (the number followed by a line break), and the scores were normalised over the candidates to give a distribution for each image and condition. The estimate was taken as the median of this distribution; the distributions of pt were flat (entropy 2.4–2.6 nats, against 1.2–1.5 in it), which inflates the expectation toward the centre of the grid, and the mode is reported as a sensitivity analysis (Multimedia Appendix 1, S8). Five conditions were used (Multimedia Appendix 2, A.3): *none*, in which the first sentence only states that what follows is the estimated nutritional content of the dish in the photo; *jp* and *us*, in which the sentence states that a Japanese or an American registered dietitian, familiar with standard single-serving portion sizes, looked at the photo and made the estimate; and *smaller* and *larger*, in which the *none* sentence is followed by a statement that the portion is smaller or larger than it looks. The *smaller* and *larger* documents thus extend the *none* document, whereas the *jp* and *us* documents differ from it in structure; the contrast between nationalities was therefore defined as JP − US. Scoring used Hugging Face Transformers 4.57 with BF16 weights on one H100; no text was generated, so the language of the output does not enter this comparison. The instruction-tuned weights were scored in the same format, and their results were compared with those of the same weights in format (a). After the Gemma 3 results had been examined, the same format was applied to InternVL3.5 8B in the checkpoint released as ‘Pretrained’ and in the final released weights, on NutriImage. The two checkpoints are not the same kind of pair as Gemma 3 pt and it. According to the technical report, the language model of InternVL3.5-8B is initialized from Qwen3-8B (whether from its base or its post-trained release is not stated), and the ‘Pretrained’ checkpoint is the model after the first, multimodal pre-training stage, whose data include multi-turn dialogue with the loss computed on response tokens; the final weights follow supervised fine-tuning and cascade reinforcement learning (offline MPO, then online GSPO). For InternVL3.5, therefore, ‘before’ and ‘after’ post-training in this article mean before and after these last two stages, and the ‘Pretrained’ checkpoint already carries instruction-like training. Because the pre-trained checkpoints of InternVL3.5 are distributed only in the repository’s own format, the vision features were computed with that code and inserted into the token embeddings of the language model (Qwen3), which was then called through Transformers as for Gemma 3; images were preprocessed as in the model card (448-pixel tiles, at most 12 plus a thumbnail). In a pilot, the pre-trained weights continued the number in ways that varied with the condition (a decimal point, a semicolon, a range), so that scoring the number followed by a line break weighted the candidates differently by condition; the estimate for InternVL3.5 was therefore taken from a score that does not depend on how the number ends (the number followed by any token that is not a digit), and the line-break score, identical in definition to that of Gemma 3, is reported alongside. Predictions for this run were written before it (Table 1, P6).

#### (e) Visible amount: same-dish pairs and before/after-meal pairs

Two sets of image pairs in which the amount differs while the dish does not were analysed in the *none* condition. First, NutriImage contains 20 pairs of the same dish served at two sizes (mean ratio of reference energy 1.74); existing outputs of Claude Sonnet 4.6 (chat), Qwen3-VL 8B (chat, MLX 4-bit), and Gemma 3 27B pt and it (scoring) were used. One author (S.N.) classified each pair by whether the difference in amount was apparent in the photographs; this was done after the aggregate results for the 20 pairs had been seen, but without sight of the outputs for individual pairs, and by one rater only. The difference that was apparent in most of the 12 pairs was the number of pieces. Second, ACETADA [11] provides smartphone photographs of 806 meals taken immediately before and after eating in a controlled-feeding study of 152 adults in Perth, Western Australia, in which foods and beverages were weighed to 0.1 g and items, weights, and nutrients were assigned by dietitians. Each photograph shows the meal on a tray with a fiducial marker for scale and standard tableware, so that, unlike NutriImage (Section 2.1), a reference for absolute size is visible in every image. The images were reduced to a long side of 1,024 pixels. The energy and nutrient values of the dataset refer to the amounts consumed, that is, the amount served less what was left, as its authors state [11]; in keeping with this, within each item name, energy divided by consumed weight was constant, whereas energy divided by served weight was not (median coefficient of variation 0.00 against 0.40). The served and the remaining energy of each meal were therefore reconstructed by us as the energy density of each item (from records of the same item with non-zero consumption) multiplied by its served and remaining weight; served minus remaining energy agreed with the recorded consumed energy (median difference 0 kcal). The remaining fraction was defined as remaining over served energy; the fraction by weight, which includes beverages such as water, is reported alongside (Spearman correlation between the two, 0.82). Two of the 806 records pointed to the photographs of another meal of the same participant and were excluded (804 pairs). Twelve meals in which the recorded consumption exceeded the served amount were excluded from the measures that use energy (792 meals) and retained for direction and ratio. Because the labels refer to consumption, the errors reported here for before-meal photographs are not comparable with those of studies that compare before-meal estimates with the consumed amounts [11].

### 2.4 Measures

Energy was read from the JSON field for energy in formats (a)–(c) and from the scored distribution in format (d). Range responses were taken at their midpoint, and responses without a closing brace or a numeric value were excluded; range responses were negligible except in Gemma 3 4B (Multimedia Appendix 1, S4). For each model and image set (a *cell*), the following were computed on the images with valid responses in the conditions compared: *baseline bias*, the mean of (estimate − reference) in the *none* condition; *instruction following*, the mean paired difference larger − smaller; the *persona shift* (JP shift), the mean paired difference JP − none, and the contrast JP − US, each with the proportions of images in which the first estimate was lower, higher, or identical (down/up/tie); and the *instruction shift* (smaller shift), the mean paired difference smaller − none, with the larger shift defined likewise. Accuracy was summarised as the median absolute percentage error (MedAPE) against the reference and Spearman’s ρ between estimate and reference. Two sets of images were used. The cell-level quantities of format (a) (Tables 2 and 4) were computed on all images with a reference value and a valid response (NutriImage: 886 of 887; SNAPMe: 1,131 of the 1,134 listed, of which the 1,128 with reference energy above zero enter MedAPE). The image-level analyses (formats (b)–(d), the size pairs, and the stratifications) were restricted to images with reference energy above 20 kcal (NutriImage: 859, the 28 excluded being items such as tea and vegetable sticks; 853–859 by condition in the crop experiments, and the 853 common to all nine conditions in Table 5; SNAPMe: 1,075), because the absolute percentage error is unstable at small reference values. For the pairs in (e), the measures were the proportion of pairs in which the smaller amount received the lower estimate, the ratio of the two estimates, and, for ACETADA, the rank correlation between this ratio and the weighed remaining fraction.

**Table 4.**
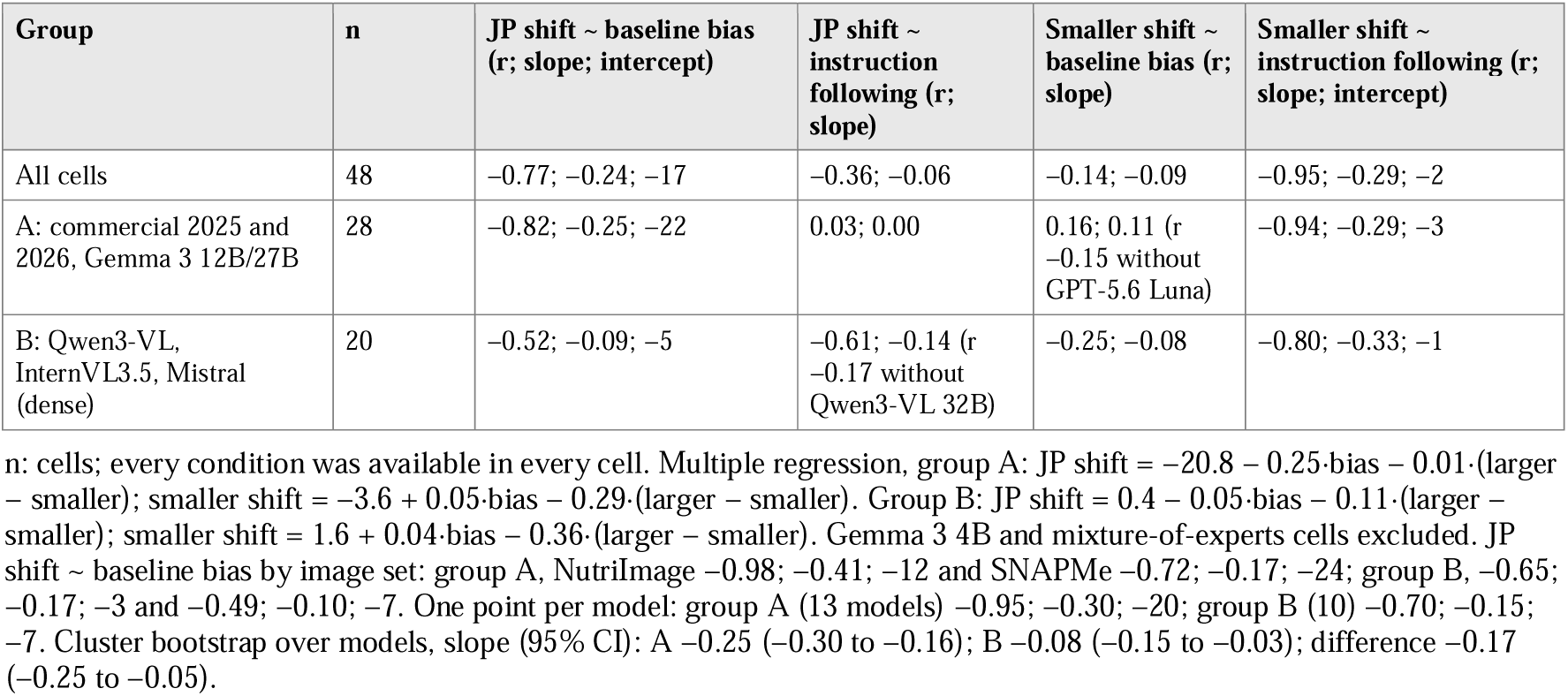
Regressions of each shift on baseline bias and on instruction following (kcal per kcal).

| Group | n | JP shift ~ baseline bias (r; slope; intercept) | JP shift ~ instruction following (r; slope) | Smaller shift ~ baseline bias (r; slope) | Smaller shift ~ instruction following (r; slope; intercept) |
| --- | --- | --- | --- | --- | --- |
| All cells | 48 | –0.77; –0.24; –17 | –0.36; –0.06 | –0.14; –0.09 | –0.95; –0.29; –2 |
| A: commercial 2025 and 2026, Gemma 3 12B/27B | 28 | –0.82; –0.25; –22 | 0.03; 0.00 | 0.16; 0.11 (r –0.15 without GPT-5.6 Luna) | –0.94; –0.29; –3 |
| B: Qwen3-VL, InternVL3.5, Mistral (dense) | 20 | –0.52; –0.09; –5 | –0.61; –0.14 (r –0.17 without Qwen3-VL 32B) | –0.25; –0.08 | –0.80; –0.33; –1 |

n: cells; every condition was available in every cell. Multiple regression, group A: JP shift = $-20.8 - 0.25 \cdot \text{bias} - 0.01 \cdot (\text{larger} - \text{smaller})$ ; smaller shift = $-3.6 + 0.05 \cdot \text{bias} - 0.29 \cdot (\text{larger} - \text{smaller})$ . Group B: JP shift = $0.4 - 0.05 \cdot \text{bias} - 0.11 \cdot (\text{larger} - \text{smaller})$ ; smaller shift = $1.6 + 0.04 \cdot \text{bias} - 0.36 \cdot (\text{larger} - \text{smaller})$ . Gemma 3 4B and mixture-of-experts cells excluded. JP shift ~ baseline bias by image set: group A, NutriImage –0.98; –0.41; –12 and SNAPMe –0.72; –0.17; –24; group B, –0.65; –0.17; –3 and –0.49; –0.10; –7. One point per model: group A (13 models) –0.95; –0.30; –20; group B (10) –0.70; –0.15; –7. Cluster bootstrap over models, slope (95% CI): A –0.25 (–0.30 to –0.16); B –0.08 (–0.15 to –0.03); difference –0.17 (–0.25 to –0.05).

**Table 5.** Estimates and persona shift of Claude Sonnet 4.6 on NutriImage under three inputs (853 images common to the three; energy, kcal).

| Measure | Full image | Dish name only | Central 25% crop |
| --- | --- | --- | --- |
| Baseline bias (none) | +17.5 | +23.4 | -16.3 |
| MedAPE, none (%) | 34.4 | 31.7 | 56.1 |
| $\rho$ with reference, none | 0.75 | 0.77 | 0.34 |
| $\rho$ with full-image estimate, none | — | 0.68 | 0.44 |
| JP – none | -17 | -13 | -44 |
| down / up / tie | 0.49 / 0.18 / 0.33 | 0.41 / 0.17 / 0.42 | 0.62 / 0.18 / 0.20 |
| JP – US | -16 | -20 | -42 |
| US – none | -2 | +7 | -2 |
| MedAPE, JP (%) | 33.0 | 28.6 | 58.0 |
| JP – none by stratum of US-condition error, lowest to highest (slope) | -4, -14, -31, -45 (-0.11) | -3, -2, -22, -40 (-0.11) | -26, -30, -70, -83 (-0.11) |
| Size pairs (12): larger > smaller / equal | 0.58 / 0.33 | 0.25 / 0.58 | 0.50 / 0.08 |
| Size pairs (12): mean ratio of estimates (reference 1.85) | 1.37 | 1.01 | 1.25 |

Strata are by the error of the estimate under the US persona, which does not enter JP – none. Strata: error < 0, 0–50, 50–150, > 150 kcal; all rows on the 853 images common to the three inputs.

Most open models returned energy on a coarse grid (multiples of 50 kcal in up to 98% of responses), so a mean paired difference can be small while the proportion of images that moved is informative; sign proportions are therefore reported with the means. Means, not medians, are used for bias and shifts so that they are on one scale in the regressions. The Wilcoxon signed-rank test was computed for each cell for reference only (Multimedia Appendix 1, Table S1b) and is not used in the text. The coarse grid and the proportion of ties are described in Multimedia Appendix 1, S6.

### 2.5 Analysis

*Predictions.* The predictions in Table 1 were derived from the two hypotheses of the previous study. For the experiments run for this study after the ladder had been aggregated (formats (c) and (d) and the ACETADA pairs), the expected outcome and the outcome that would count against it were written down before the run; these are reported with the results, including those that were not met. The remaining predictions (crop, size pairs, P2, and P5) were formulated on data already in hand, and Table 1 marks the distinction.

*Cell-level regressions.* Each shift was regressed across cells on baseline bias and on instruction following by ordinary least squares, with Pearson’s r; a multiple regression on both variables was fitted as a check. Regressions were run on all cells and within two groups defined after inspection of Figure 2: (A) commercial models of both generations and Gemma 3 12B/27B, and (B) Qwen3-VL, InternVL3.5 (dense models), and the two Mistral models. The grouping is descriptive, and no test of the difference between slopes is reported. Gemma 3 4B and the two mixture-of-experts models were excluded from the regressions. Cells lacking the *smaller* condition or instruction following enter only the regressions that do not require them, so n differs between regressions (Table 4). Under H2, if baseline bias arose entirely from the serving a model assumes and the persona replaced that serving by a correct one, the shift would equal minus the bias (slope −1, intercept 0) and would not depend on how strongly the model follows instructions; a partial replacement gives a slope between 0 and −1.

**Figure 2.**
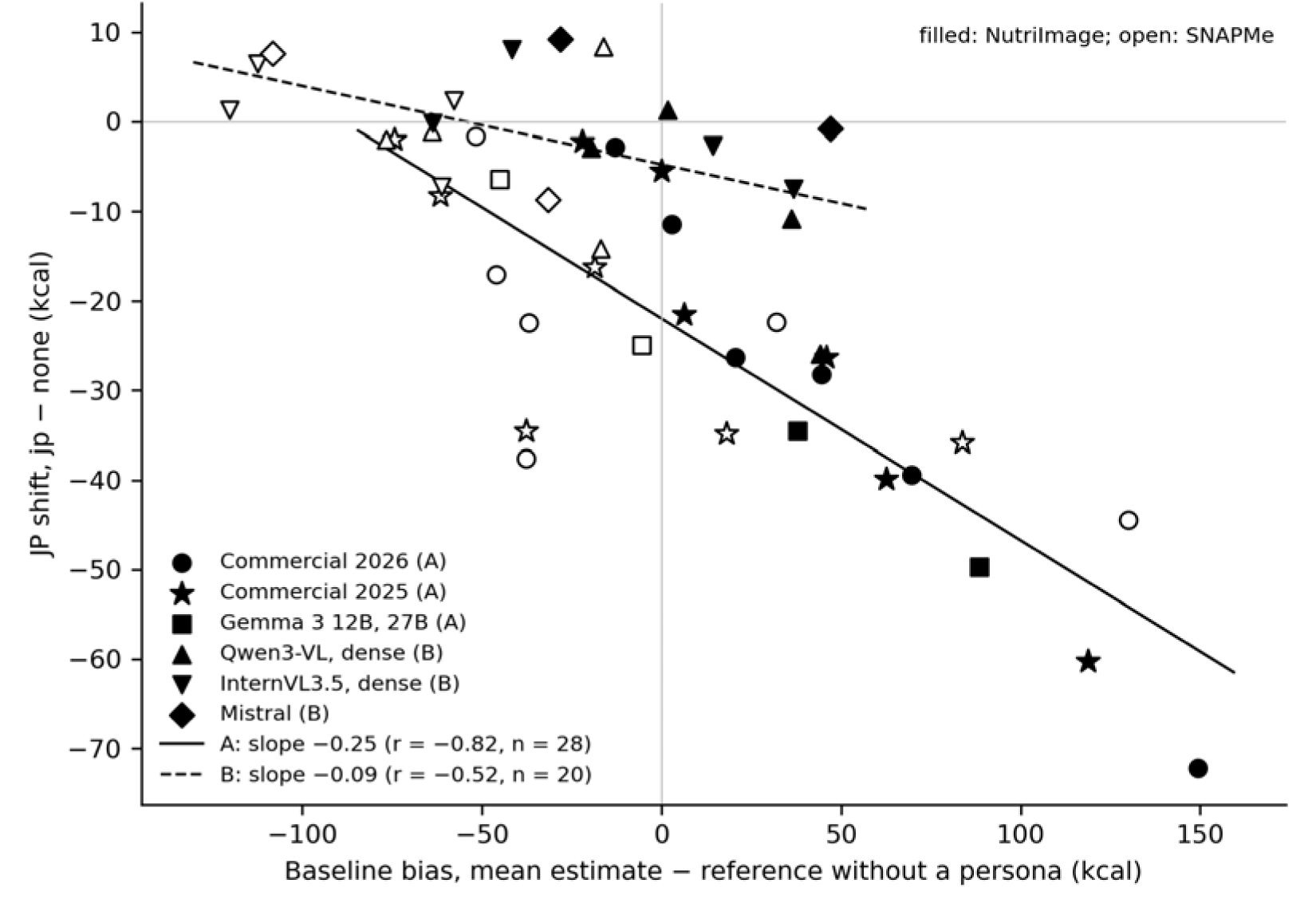
JP shift against baseline bias in 48 cells, by group. Each point is one cell (model × image set); filled, NutriImage; open, SNAPMe. Group A (28 cells): the commercial models of 2025 and 2026 and Gemma 3 12B and 27B; group B (20 cells): the dense models of Qwen3-VL, InternVL3.5, and Mistral. Lines are ordinary least-squares fits within each group (Table 4). Gemma 3 4B and the two mixture-of-experts models are not shown (Section 2.5). Values: master_cells_energy_v2_20260921.csv; script: fig1.py.

The alternative considered, that the persona is followed as a weak instruction about amount, predicts a shift related to instruction following and to the smaller shift, and unrelated to bias. This formulation was made on the data in hand (Table 1). Because each model contributes two cells, regressions were also run on one point per model and by image set, and intervals for slopes and their difference were obtained by resampling models with replacement (5,000 samples). Because bias and shift are computed on the same images, each image set was also split at random into halves, bias computed on one half and the shift on the other, 200 times (Multimedia Appendix 1, S1).

#### Pre-trained against post-trained weights

The change with post-training was assessed within images: for each image, (JP − US) and (larger − smaller) in the post-trained weights minus the same quantities in the pre-trained weights, with the mean of these paired differences and a bootstrap interval over images (2,000 resamples). This was done for each size and image set of Gemma 3 and for InternVL3.5 8B.

#### Stratification by identification

In format (b), the JP shift at the 25% crop was computed separately for images judged SAME and DIFF, and compared with the shift of the same images at full size.

#### Form of a response across images

To describe how the response to a description varies with the error of the individual estimate, contrasts between two conditions were stratified by the error of the estimate in a condition that enters neither term of the contrast. The contrasts were JP − US, jp_home − us_home, smaller − US, and larger − US, and the stratifying condition was *none* (estimate without a persona minus reference), in four strata (< 0, 0–50, 50–150, > 150 kcal). Two kinds of apparent slope arise otherwise. (a) If the stratifying estimate is one term of the difference, as when persona − none is set against the error without a persona (Figure 3 of the previous study), the variation between two requests for the same image produces a negative slope by itself. (b) The estimate without a persona also carried deviations specific to the image that recurred between runs, so that a difference from *none*, stratified by the error of *none*, showed a slope in conditions whose mean shift was zero, even when the error was taken from a separate run (Multimedia Appendix 1, S12). A contrast between two conditions other than *none*, stratified by the error of *none*, carries neither.

**Figure 3.**
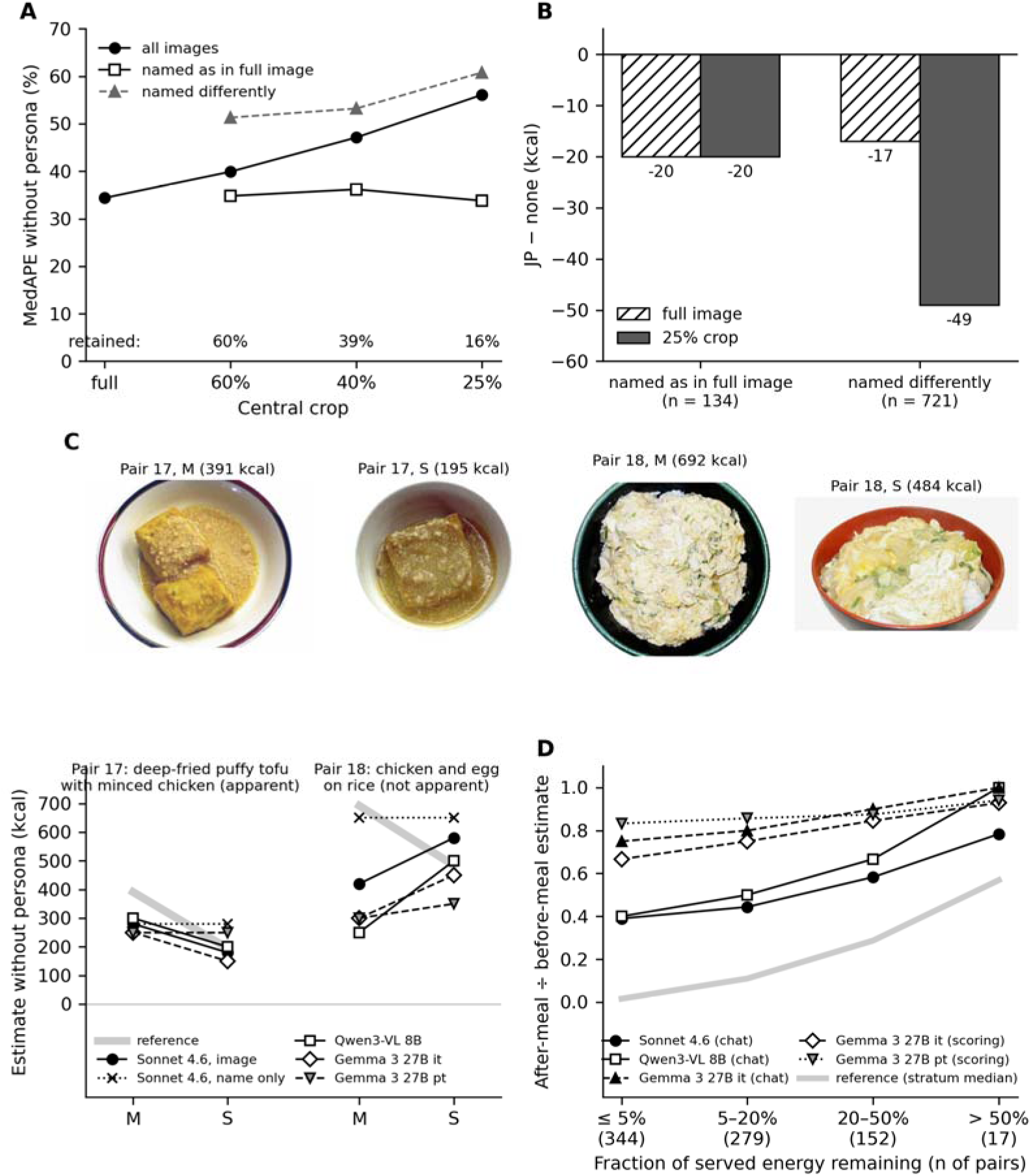
Locus of the persona shift. (A) MedAPE of Claude Sonnet 4.6 without a persona at each central crop, for all images and by whether the dish was named as in the full image; percentages, identification retention. (B) JP shift at full size and at the 25% crop, by identification at the 25% crop. (C) Photographs (top; NutriImage 0490, 0491, 0540, 0541, with reference energy) and estimates without a persona (bottom left) for two same-dish pairs served at two sizes (M, S): pair 17, difference apparent in the photographs (two pieces against one); pair 18, not apparent (a bowl filling the frame); grey, reference values. Photographs are the authors’ own (NutriImage v1.1, doi:10.5281/zenodo.21961032). (D) ACETADA: ratio of the after-meal to the before-meal estimate by the fraction of served energy remaining; grey, the reference fraction (stratum median).

For the chat format, images with reference energy above 20 kcal and with estimates above zero in both terms of the contrast and in *none* were used. The contrast was taken as the mean of log2 of the ratio of the two estimates, and is given alongside in kcal and as the proportion of images lowered, which does not depend on the choice of scale. The *step* was defined as the mean of the three strata with positive error minus the stratum with negative error, so that a negative step indicates a larger lowering where the model overestimated; strata with fewer than 30 images were not used. The form was described only for cells in which the mean contrast was −10 kcal or beyond. Intervals are percentile 95% intervals from 2,000 resamples of the images within a cell (commercial models of 2026 and open-weight models; point estimates only for the six models of the previous study). The way of reading these forms was settled on the outputs of the six models of the previous study and then applied unchanged to all models; the forms are reported as description and not as tests of predictions. For Gemma 3 under scoring (Section 3.4), the same stratification is given in kcal, with the slope of the per-image difference on the error and r; no inference is drawn from these beyond description. The crop and name-only experiments have only the *none*, *jp*, and *us* conditions; there JP − none was stratified by the error under *us*, which does not enter that difference, and this is retained as a secondary description (Section 3.2), the main analysis of form being JP − US stratified by the error of *none*.

#### Comparison with numerical calibration

The images of each cell (reference energy above 20 kcal) were split at random into halves 200 times. On one half a correction was determined, either (i) the mean bias, to be subtracted, or (ii) the median ratio of estimate to reference, by which the estimates are divided; MedAPE after each correction was evaluated on the other half, alongside MedAPE under *none*, *jp*, and *smaller* on the same images. Over the 200 splits, each MedAPE is reported as the median with the 2.5th and 97.5th percentiles, and differences between methods are computed within the same split and summarized in the same way; a difference is described as holding across splits when its percentile interval excludes zero.

Analyses used Python 3.11 with NumPy, pandas, and SciPy. No correction for multiple comparisons was applied; *P* values are given for reference.

### 2.6 Execution settings

Open-weight models were run in format (a) with vLLM 0.11 on NVIDIA H100 GPUs with BF16 weights, at temperature 0, with an output limit of 256 tokens and one image per prompt; Qwen3-VL was also run with the official FP8 weights (Multimedia Appendix 1, S2), and Qwen3-VL 2B, 4B, and 8B with MLX 4-bit weights on Apple silicon (Multimedia Appendix 1, S3). Image preprocessing followed each model’s default (dynamic resolution in Qwen3-VL, fixed 896 pixels in Gemma 3, tiling in InternVL3.5). The 2026 interfaces of two vendors did not accept a temperature of 0 for the models used, so all six commercial models were run at the vendors’ default temperature with one repetition, as in the previous study. Extended thinking was set to the lowest option each interface accepted (Anthropic: disabled; OpenAI: reasoning effort ‘none’; Gemini 3.8 Flash: thinking budget 0; Gemini 3.1 Pro: thinking level ‘low’). For the two Gemini models, thinking tokens counted against the output limit and truncated the JSON at 256 tokens, so the limit was raised to 2,048 for these two models only; the visible JSON was about 100 tokens in all cases. Responses without a closing brace were treated as truncated and requested again, and remaining failures were excluded. The 2025 results had been obtained with an output limit of 500 tokens and, for SNAPMe, on 1,272 images, with the *smaller* and *larger* conditions on a subset. Differences between environments are summarised in Table 3.

### 2.7 Ethical considerations

Ethical review and approval were waived for this study because it used only pre-existing, de-identified image sets and involved no new collection of data from human participants, as in our previous study [1]: SNAPMe is openly available, NutriImage is available for academic research under restricted access, and ACETADA is distributed by its authors for non-commercial research. No person is shown in the photographs of NutriImage. Informed consent was not applicable.

## Results

### 3.1 What the estimate rests on: identification, dish name, and visible amount (P1a, P1b)

#### Under cropping, accuracy followed identification

In the crop experiment (Claude Sonnet 4.6, NutriImage), without a persona, retention fell from 60.0% at the 60% crop to 39.4% at 40% and 15.6% at 25%, and MedAPE rose from 34.4% (full image) to 39.9%, 47.1%, and 56.1%; within the images named identically, MedAPE stayed near the full-image value at every crop (34.8%, 36.2%, 33.8%), and the loss of accuracy was confined to those named differently (51.3%, 53.2%, 60.8%) (Figure 3A; n = 857 images with reference energy above 20 kcal). Over all images, the 25% crop lowered Spearman’s ρ against the reference from 0.75 to 0.34 (Table 5).

#### The dish name alone gave similar estimates

When the image was withheld and only the English dish name of the reference record was given, the estimates of the same model correlated with its image-based estimates at ρ = 0.68 (0.71 in the 512 images named identically at the 60% crop); we had expected a value above 0.7. Baseline bias was +23.4 kcal (image, +17.5 kcal), ρ against the reference 0.77 (0.75), and MedAPE 31.7% (34.4%). The name given was that of the reference record, not the model’s own identification, so this condition corresponds to identification without error; the small difference in accuracy is not an argument for withholding the image.

#### The same dish at two amounts

NutriImage contains 20 pairs of the same dish served at two sizes (mean reference energy ratio 1.74). One author (S.N.) classified the pairs by whether the difference in amount was apparent in the photographs, after seeing the aggregate proportions for the 20 pairs but not the outputs for individual pairs: apparent in 12 (reference ratio 1.85; in most, two pieces against one), not apparent in 7 (bowls filling the frame), and apparently reversed in 1. In the 12 pairs, the larger serving received the higher estimate in 0.58 of pairs with Claude Sonnet 4.6 (mean ratio of estimates 1.37), 0.75 with Qwen3-VL 8B (MLX 4-bit; 1.30), and 0.42 with Gemma 3 27B, both instruction-tuned (1.17) and pre-trained (1.12) (Multimedia Appendix 1, S9). Figure 3C shows one pair of each kind: in the pair with an apparent difference (two pieces against one), three of the four image-based estimates were lower for the smaller serving, that of Gemma 3 27B pt was the same, and the name-only estimates were identical; in the pair without one (a rice bowl filling the frame), all four image-based estimates were higher for the smaller serving. With the dish name alone, the corresponding values were 0.25 and 1.01; the names given for the two servings of these 12 pairs were identical or differed only in typography, except in one pair in which the name of the smaller serving carried a size letter (Multimedia Appendix 1, S9). In the 7 pairs without an apparent difference the proportion was 0.29 for every model. Twelve pairs do not allow any of these proportions to be distinguished from one half; the ratios of 1.1–1.4 against a reference ratio of 1.85 suggest that visible amount entered the estimate weakly and to a degree that differed by model.

#### Before and after the meal

In the 804 ACETADA pairs, the median fraction of the served energy that remained after the meal was 0.07 (0.08 by weight), and 169 pairs had 20% or more remaining. The after-meal estimate was lower than the before-meal estimate in 0.97 of pairs with Claude Sonnet 4.6 and with Qwen3-VL 8B, 0.74 with Gemma 3 27B it in the chat format (equal in 0.20), 0.87 with the same weights under likelihood scoring, and 0.78 with Gemma 3 27B pt (Table 6; Figure 3D). The size of the decrease was much smaller than the reference: in the 344 pairs in which 5% or less of the served energy remained (median 1.6%), the after-meal estimate was 0.39 (Sonnet 4.6), 0.40 (Qwen3-VL 8B), 0.75 and 0.67 (Gemma 3 27B it, chat and scoring), and 0.83 (Gemma 3 27B pt) of the before-meal estimate. The ratio rose with the remaining fraction in the two models that read the direction reliably (Qwen3-VL 8B, 0.40 to 1.00 across strata; Sonnet 4.6, 0.39 to 0.78) and changed little in Gemma 3 27B pt (0.83 to 0.94; with the mode of the scored distribution in place of its median, Multimedia Appendix 1, S11); its rank correlation with the remaining fraction was 0.29, 0.61, 0.30, 0.39, and 0.14 in the order of Table 6 (by weight: 0.24, 0.48, 0.21, 0.29, 0.07). What the models wrote about the after-meal photographs differed. In the 344 pairs with 5% or less remaining, Claude Sonnet 4.6 gave the after-meal photograph a name such as ‘finished meal remnants’ in 87% (298 of 342 with both estimates) and still returned a median of 280 kcal for these (100 kcal or more in 78%), its stated basis often being the meal that the residue suggested; Qwen3-VL 8B stated that the tray was empty in 15% (51 of 343), and then returned 0 kcal in 82% of these, and otherwise described the residue as a meal; Gemma 3 27B it seldom mentioned that the food had been eaten (Multimedia Appendix 1, S11). The task prompt asks for ‘the nutritional content of this meal’, so that the after-meal estimates reflect in part an estimate of the meal that had been served and not of what remained, and the ratios above cannot be read as a measure of how visible amount is read. Under scoring, the distribution of the pre-trained weights was nearly flat (median entropy 3.1 against a maximum of 3.4 for 30 candidates), so that its median moves little between images and the ratio of 0.83 depends on the format; the lower limit of the candidates (50 kcal) was reached in 1% of after-meal photographs or fewer. These photographs include a fiducial marker and standard tableware, although its dimensions were not given to the models. Of the expectations written before these runs, those for the direction in Claude Sonnet 4.6 and Qwen3-VL 8B and for the ratio in Gemma 3 27B pt were met; the direction in Gemma 3 27B it was read more often, and the correlations were lower, than expected (Multimedia Appendix 1, S11). Against the served energy (median 725 kcal), the before-meal estimates were low by 130–237 kcal in all five cases (MedAPE 21–30%; ρ 0.67–0.80).

**Table 6.**
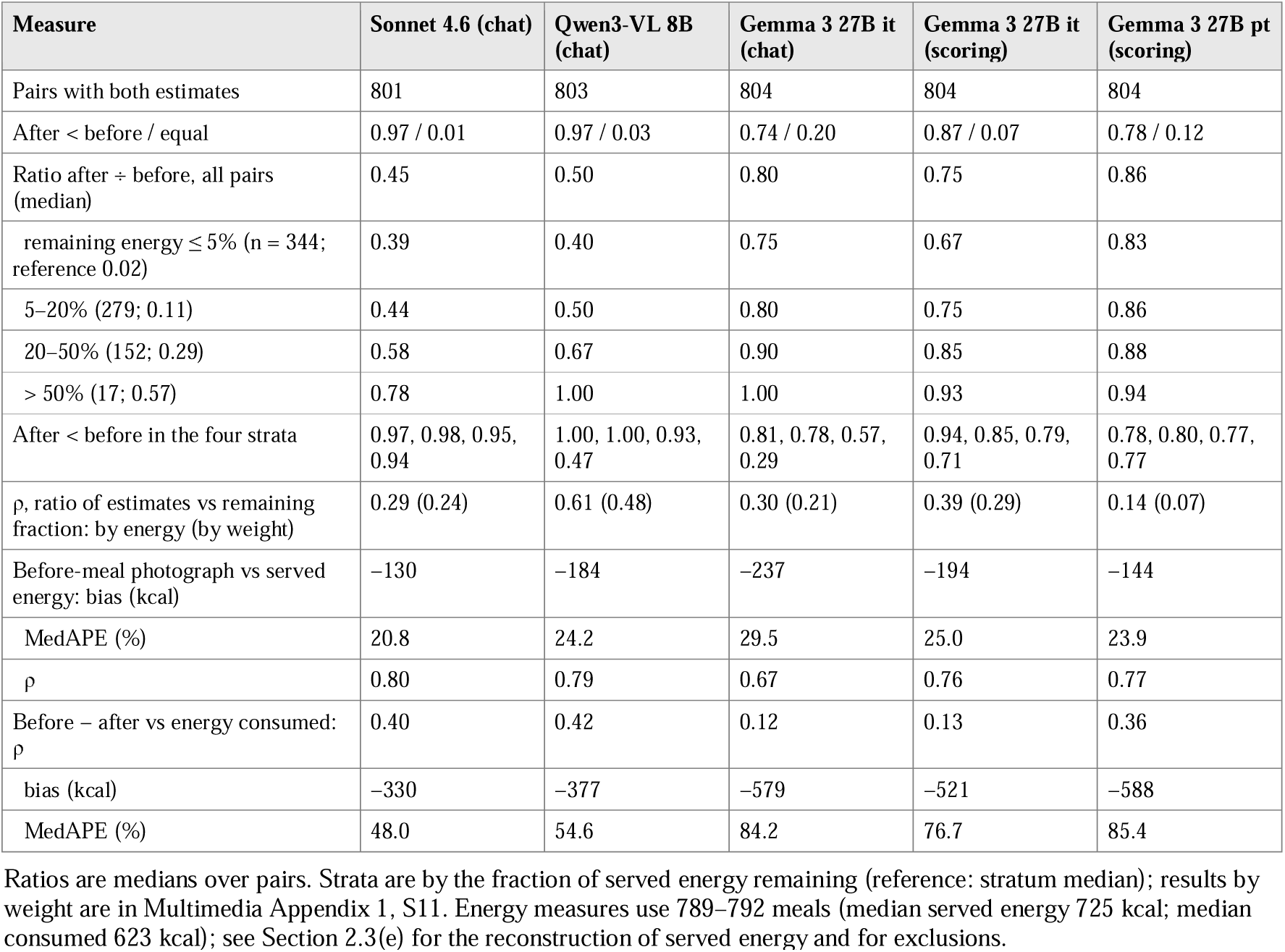
Before- and after-meal photographs of the same tray (ACETADA, 804 pairs; no persona).

These results are consistent with the two-stage account in that the estimate appeared to be anchored on the amount associated with a dish name and to be corrected only in part by the visible amount: the direction of a change in amount was read, reliably by two of the models, but its size was compressed toward the anchor in the same-dish pairs; in the before/after pairs the compression cannot be separated from the models’ estimating the meal that had been eaten.

### 3.2 Where the persona acts: identification and naming under the persona (P1c)

#### Identification retention did not differ by persona

In the crop experiment (Claude Sonnet 4.6, NutriImage), the dish named from the central 25% of the image was judged to be the same as that named from the full image in 15.6% of images without a persona (134 of 857), 17.1% under the JP persona, and 16.7% under the US persona. Retention was judged within each condition and does not show whether the persona changed the name itself; this is examined below.

#### Where identification was retained, the persona shift did not depend on image extent

In the 134 images named identically at the 25% crop, the JP shift was −20 kcal (95% CI −31 to −9; bootstrap), close to that of the same images at full size (−20 kcal; −28 to −13). In the 721 images named differently, the JP shift at the 25% crop was −49 kcal (−57 to −41), against −17 kcal (−21 to −12) at full size (Figure 3B); the looser criterion gave −22 and −50 kcal at the 25% crop (Multimedia Appendix 1, S10). The first result is what P1a predicts, within the precision that 134 images allow: the within-image difference between the shift at the 25% crop and at full size was 0 kcal (95% CI −11 to +12) in these images, against −32 kcal (−41 to −24) in the 721 others. The second was not predicted. Over all images, the JP shift at the 25% crop was −44 kcal (Table 5); at full size, when images were stratified by the error of the estimate under the US persona, which does not enter JP − none (a secondary description; the main analysis of form is that of Section 3.5), the shift was smallest where the model underestimated and largest where it was overestimated by more than 150 kcal (−4, −14, −31, and −45 kcal across the four strata; slope against the error −0.11). At the 25% crop the slope was the same (−0.11) but the shift was already −26 kcal in the lowest stratum (Table 5; Multimedia Appendix 1, S10), that is, the dependence on error was retained and a roughly constant lowering was added. We report this as an observation and return to it in the Discussion.

#### With the dish name alone, the persona shift was similar

The JP shift was −13 kcal with the name alone (down/up/tie 0.41/0.17/0.42) and −17 kcal with the image (0.49/0.18/0.33); JP − US was −20 and −16 kcal. The dependence on baseline error, stratified as above, had the same form (−3, −2, −22, and −40 kcal across the strata, slope −0.11, against −4, −14, −31, and −45 kcal with the image). The images that moved were not the same under the two inputs: the per-image JP shifts correlated at 0.04.

#### Names given with and without the persona

The name returned for an image under a persona was compared with that returned without a persona. By the model judge, the two names denoted different dishes in 20% of the full images under the US persona (35% at the 25% crop), which gives the variation to be expected between two requests at the default temperature, and in 42% (73%) under the JP persona. Before the none–us comparison was run, we had expected a disagreement of 20–30% at full size and a US shift within ±5 kcal whether or not the name differed; both were met (−3 kcal, n = 683, and +3 kcal, n = 175). The excess under the JP persona, however, was largely a matter of wording across languages. In 100 full-image pairs rated by one author against the original menu, the name without a persona was correct in 62 and the name under *jp* in 65 (discordant pairs 3 and 6; exact McNemar *P*=.51), and the two names denoted different dishes in 22 (95% CI 15–31; taken as different where only one name was correct, as the same where both were correct, and as rated where neither was), against 46 of the same 100 by the model judge (agreement 68%, κ = 0.33). By human rating, then, the JP persona neither improved identification nor renamed dishes more often than is seen between two requests, and the model judge was not valid across languages. The strata formed by the judge nevertheless differed in the JP shift: −9 kcal where it found the names the same (n = 494; lower in 0.43 of images) and −28 kcal where it found them different (n = 364; 0.58); at the 25% crop, −16 kcal (n = 228) and −55 kcal (n = 628); the difference was also present among images whose name was stable under *us* (full size, −25 kcal, n = 257, against −9 kcal, n = 426) (Multimedia Appendix 1, S10). Since most of the judged differences were not different dishes, the 96 rated pairs with reference energy above 20 kcal were divided into three groups. Where the two names denoted the same dish and the judge too had found them the same (n = 47), the estimate under *jp* was lower than that without a persona in 0.30 of pairs and higher in 0.32 (mean +3 kcal; 95% CI −9 to 15; median 0). Where they denoted the same dish but the judge had found them different, that is, where the wording differed (for example, sautéed greens without a persona and ohitashi under *jp*; n = 27), it was lower in 0.59 and higher in 0.07 (mean −28 kcal; −45 to −13; median −10). Where they denoted different dishes (n = 22), it was lower in 0.50 and higher in 0.14 (mean −37 kcal; −70 to −7; median −5); this group was underestimated without a persona (mean error −41 kcal), so its lowering is not accounted for by dependence on baseline error. The means of the last two groups were influenced by a few large differences, and the US shift was −6, −1, and −9 kcal in the three groups. In this sample, then, a lowering in one direction was seen where the description of the dish changed under the persona, in wording or in identity, and not where it was unchanged. The sample is small and was rated by one author, and in the whole set the stratum in which the judge found the names the same still showed −9 kcal.

With respect to P1c, the persona did not alter how well identification survived cropping nor how often the name was correct, and with the name held fixed (the name-only condition) a shift of −13 kcal was obtained, against −17 kcal with the image. In the rated sample the lowering accompanied a change in the description of the dish under the persona, in wording or identity, and was absent where the description was unchanged; this suggests that the persona may act through how the dish is described, the description serving as the key with which an amount is retrieved, more than through a different amount for an unchanged description. The name-only condition does not contradict this, since there the input name was fixed but the model still returned its own name under the persona: in 22% of name-only responses under *jp* the returned name differed from the one given (in 30% it contained Japanese script), against 9% without a persona and 11% under *us*, where the difference was mostly a corrected spelling. The identification itself was correct in about two thirds of the rated images. The crop and name-only experiments used one model; the chat format of the ladder does not return a dish name.

### 3.3 Channel: relation of the persona shift and the instruction shift to baseline bias and instruction following (P2)

Table 2 gives baseline bias, the shifts, and instruction following for all 54 cells. In the 48 cells analysed, baseline bias ranged from −120 kcal (InternVL3.5 4B, SNAPMe) to +149 kcal (GPT-5.6 Luna, NutriImage). Instruction following (larger − smaller) ranged from near zero in the smallest open models to several hundred kcal in commercial models and increased with model size within every open family (NutriImage: +4, +22, +44, and +120 kcal in Qwen3-VL 2B to 32B; +4, +9, +15, and +19 kcal in InternVL3.5 2B to 14B; +28, +134, and +211 kcal in Gemma 3 4B to 27B). GPT-5.6 Luna was the single model with a reversed sign (−15 and −36 kcal; Multimedia Appendix 1, S4). Across the 48 cells, baseline bias and instruction following were only weakly correlated (r = 0.20).

#### The JP shift depended on baseline bias and not on instruction following

Across 48 cells the JP shift was related to baseline bias (r = −0.77, slope −0.24 kcal per kcal); it was less closely related to instruction following (r = −0.36, slope −0.06), and not at all within group A (r = 0.03) (Figure 4A, B; Table 4). In the multiple regression within group A, the coefficient on baseline bias was −0.25 and that on instruction following −0.01. Gemini 3.1 Pro, with the largest instruction following of any model (+403 kcal, NutriImage), shifted by −11 kcal under the JP persona at a baseline bias of +3 kcal; GPT-5.6 Luna, whose instruction following was reversed, shifted by −72 kcal at a baseline bias of +149 kcal.

**Figure 4.**
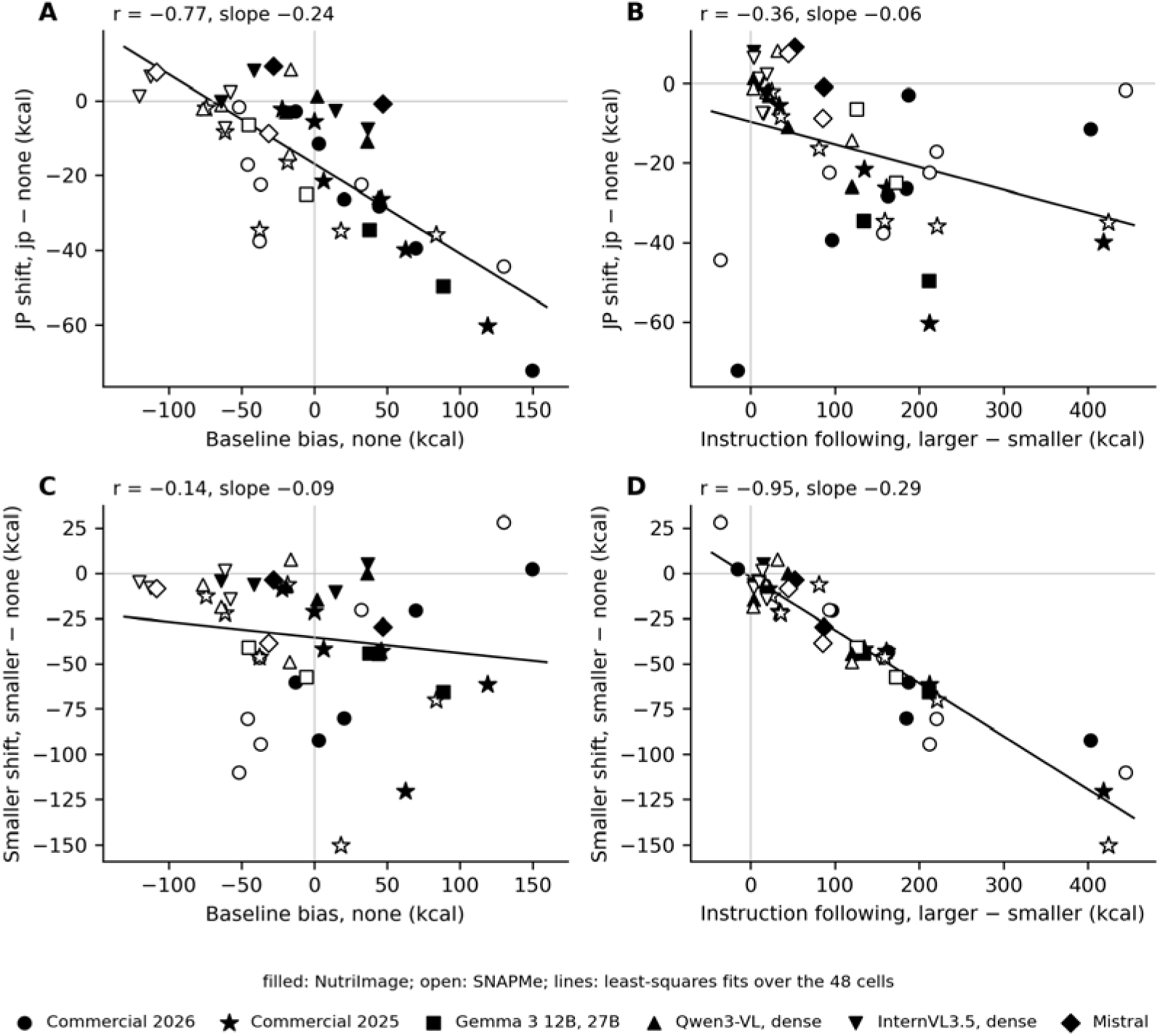
The JP shift and the smaller shift against baseline bias and against instruction following, 48 cells.(A) JP shift against baseline bias; (B) JP shift against instruction following (larger − smaller); (C) smaller shift against baseline bias; (D) smaller shift against instruction following. Each point is one cell; filled, NutriImage; open, SNAPMe; markers as in Figure 2. Lines and the r and slope shown are least-squares fits over all 48 cells (Table 4, first row). Values: master_cells_energy_v2_20260921.csv; script: fig3.py. *The smaller shift depended on instruction following and not on baseline bias.* Across the 48 cells, the smaller shift was related to instruction following (r = −0.95, slope −0.29) and not to baseline bias (r = −0.14) (Figure 4C, D). Gemini 3.1 Pro, with a baseline bias near zero, shifted by −92 kcal (NutriImage) and −110 kcal (SNAPMe) under the instruction. The larger shift behaved likewise (r = 0.99 with instruction following, 0.22 with baseline bias). Within group A the smaller shift was not related to baseline bias (r = 0.16; −0.15 without GPT-5.6 Luna); in no analysis was the smaller shift related to baseline bias in the direction shown by the JP shift.

#### The two shifts were only moderately related to each other

Across cells the JP shift and the smaller shift correlated at r = 0.35 (Spearman 0.48), and, in the cells in which the smaller shift was at least 10 kcal in size, their ratio ranged from near zero (Gemini 3.1 Pro, SNAPMe: −2 against −110 kcal; Gemini 3.8 Flash, NutriImage: −3 against −60 kcal) to 1.9 (GPT-5.6 Sol, NutriImage: −39 against −21 kcal). Within images, the rank correlation between the two paired differences was 0.10 to 0.51 in group A and 0.30 to 0.71 in group B, where most estimates lie on a grid of 50 kcal (median over all cells, 0.43; Multimedia Appendix 1, S5). In InternVL3.5, instruction following increased fivefold from 2B to 14B while the JP shift stayed between −7 and +8 kcal.

#### Models and image sets

The cells are not independent: each model contributes two cells, and baseline bias was largely a property of the model (group A: r = 0.92 between the two image sets). With one point per model (means over the two sets), the relation in group A was as close as with cells (13 models; slope −0.30, r = −0.95). Fitted separately by image set, the group-A slope was −0.41 on NutriImage (r = −0.98, intercept −12 kcal) and −0.17 on SNAPMe (r = −0.72, intercept −24 kcal); the pooled value of −0.25 lies between the two. Within models, the lower baseline bias on SNAPMe (by 52 kcal on average) was not accompanied by a smaller shift (slope of the change in shift on the change in bias, −0.03; r = −0.05; for example, Claude Haiku 4.5: bias +44 and −38 kcal, shift −28 and −38 kcal). The relation of the shift to baseline bias is therefore a relation between models: models that tend to overestimate gave lower estimates under the persona, on both image sets.

#### The contrast JP – US

Because the estimate without a persona enters both baseline bias and the JP shift, the same regression was made for JP − US, which does not contain it. Across the 48 cells JP − US was related to baseline bias at r = −0.66 (slope −0.17), and in group A at r = −0.71 (slope −0.19; 28 cells); in group B the relation was weak (r = −0.24, slope −0.04). GPT-5.6 Luna was lowered by every persona, including the US persona and the dietitian without nationality (US shift −32 and −19 kcal; neutral dietitian −20 and −11 kcal); without its two cells, the values in group A were r = −0.78 and slope −0.25, and the US shift was unrelated to baseline bias (r = 0.10 in 26 cells, against −0.38 in all 28). The conclusion drawn from the JP shift thus held for a contrast that does not share a term with baseline bias. *Control conditions.* In group A, the mean shift across the 28 cells was −26 kcal under *jp* and −20 kcal under *jp_home*, against −1 kcal under *us*, +4 kcal under *us_home*, and −3 kcal under *neutral_dietitian* (Table 2; Multimedia Appendix 1, Table S1). The lowering thus accompanied the Japanese descriptions whether or not the persona was a dietitian, and did not accompany the descriptions of a dietitian without the nationality; the US descriptions mention portion or serving sizes in the same words as the Japanese ones.

#### Group B

Within Qwen3-VL, InternVL3.5, and Mistral (20 cells) all JP shifts were small (−26 to +9 kcal) and the associations correspondingly weak: r = −0.52 with baseline bias and r = −0.61 with instruction following, the latter carried by the two Qwen3-VL 32B cells (−0.17 without them). The dissociation is therefore seen in group A, where the shifts are large enough to separate the two variables; in group B the variable on which the persona shift depends could not be identified.

#### Description

The regression of the JP shift on baseline bias had an intercept of −22 kcal in group A (−17 kcal over all cells; −5 kcal in group B): at zero baseline bias the commercial models and Gemma 3 still gave estimates about 20 kcal lower under the persona (cluster bootstrap over models, 95% CI −27 to −18 kcal). A part of the persona contrast that is uniform across images was seen directly when the contrast was stratified by the error without a persona (Section 3.5): the lowering was small, but not zero, where the model underestimated, and on SNAPMe it was of similar size in all strata; for Japanese dishes, in some models, a further part was added in overestimated images. The intercept is in keeping with the uniform part. An account of the intercept from the form of the shift across images alone would apply only to NutriImage, since on SNAPMe the contrast showed no step.

### 3.4 Training stage: pre-trained and post-trained weights of Gemma 3 (NutriImage and SNAPMe) and InternVL3.5 8B (NutriImage) (P3, P4, P6)

#### NutriImage (P3)

Gemma 3 4B, 12B, and 27B were scored in the narrator format as pre-trained (pt) and instruction-tuned (it) weights (Table 7; Figure 5). Relative to the document without a narrator, both narrators raised the median estimate on NutriImage (pt: JP +28 to +81 kcal, US +125 to +135 kcal; it: JP +10 to +61 kcal, US +44 to +83 kcal). Because the document without a narrator differs in structure from those with one, the contrast between nationalities is read from JP − US, and the design does not determine how much of it is a lowering under JP and how much a raising under US.

**Figure 5.**
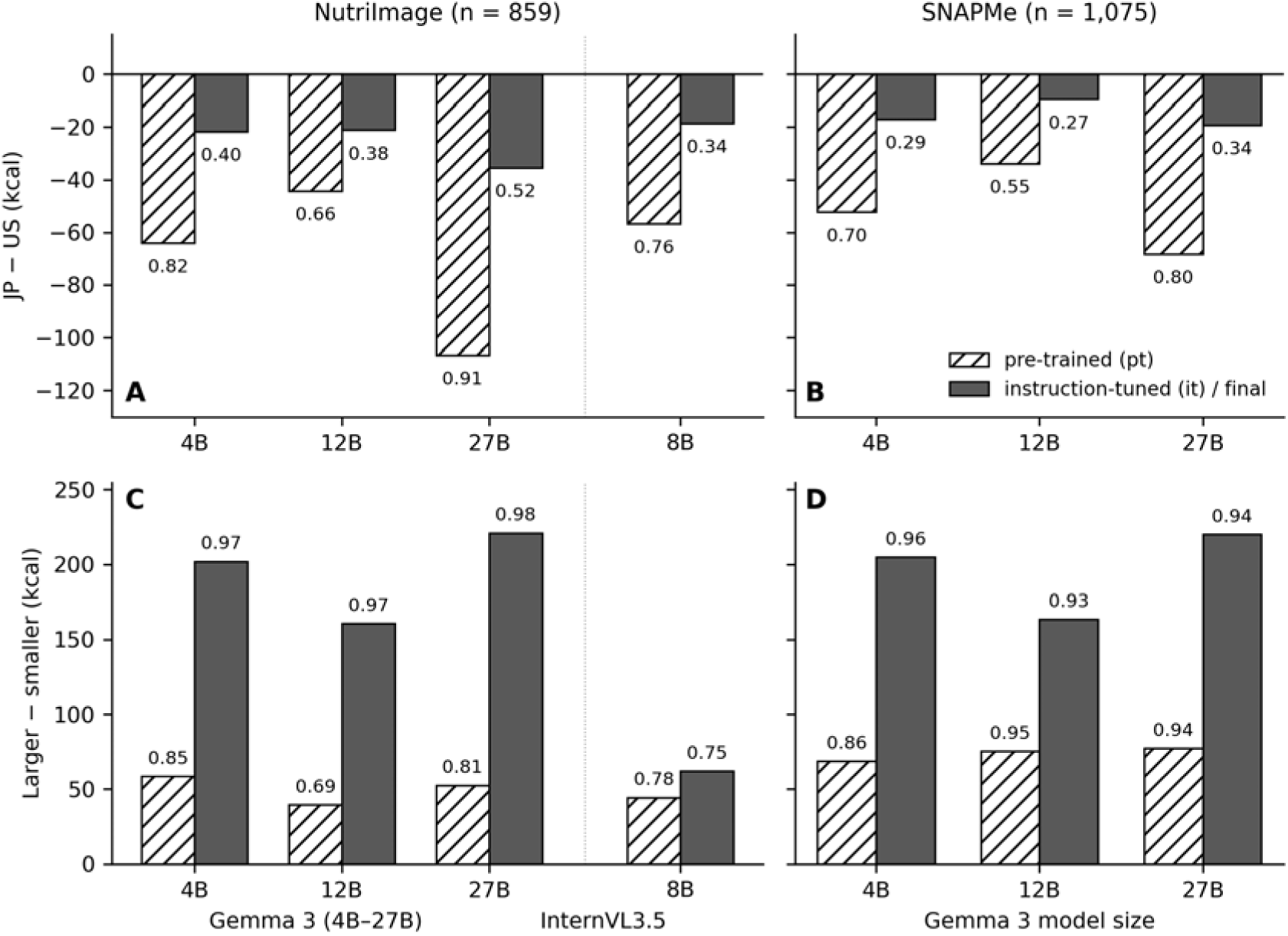
Pre-trained and post-trained weights of Gemma 3 and of InternVL3.5 8B under likelihood scoring. (A, B) Mean paired difference JP − US; (C, D) larger − smaller; NutriImage (left) and SNAPMe (right). Numbers at the bars are the proportions of images with JP < US (A, B) and larger > smaller (C, D). Estimates are medians of the scored distribution over 30 candidate values; for InternVL3.5 8B (A, C, right of the dotted line; NutriImage only) the score is the one that does not depend on how the number ends (Section 2.3(d)).

**Table 7.** Pre-trained (pt) and post-trained (it, final) weights of Gemma 3 and InternVL3.5 8B under likelihood scoring (energy, kcal; median of the scored distribution).

| Model | Set | n | Bias (none) | MedAPE (%) | $\rho$ | JP – none | US – none | JP – US | JP<US / JP>US | Larger – smaller | L>S |
| --- | --- | --- | --- | --- | --- | --- | --- | --- | --- | --- | --- |
| 4B pt | NI | 859 | –5 | 40.5 | 0.71 | +67 | +131 | –64 | 0.82 / 0.01 | +59 | 0.85 |
| 4B it | NI | 859 | +56 | 46.2 | 0.68 | +61 | +83 | –22 | 0.40 / 0.12 | +202 | 0.97 |
| 12B pt | NI | 859 | –13 | 41.8 | 0.63 | +81 | +125 | –44 | 0.66 / 0.05 | +40 | 0.69 |
| 12B it | NI | 859 | +85 | 55.1 | 0.71 | +23 | +44 | –21 | 0.38 / 0.15 | +161 | 0.97 |
| 27B pt | NI | 859 | +42 | 44.0 | 0.72 | +28 | +135 | –107 | 0.91 / 0.02 | +53 | 0.81 |
| 27B it | NI | 859 | +74 | 51.0 | 0.71 | +10 | +46 | –35 | 0.52 / 0.14 | +221 | 0.98 |
| 4B pt | SN | 1075 | –43 | 34.5 | 0.72 | +41 | +93 | –52 | 0.70 / 0.02 | +69 | 0.86 |
| 4B it | SN | 1075 | –10 | 32.5 | 0.75 | +42 | +59 | –17 | 0.29 / 0.14 | +205 | 0.96 |
| 12B pt | SN | 1075 | +30 | 38.8 | 0.74 | +22 | +56 | –34 | 0.55 / 0.06 | +75 | 0.95 |
| 12B it | SN | 1075 | +24 | 34.3 | 0.74 | +25 | +34 | –9 | 0.27 / 0.16 | +164 | 0.93 |
| 27B pt | SN | 1075 | +38 | 35.6 | 0.76 | –24 | +45 | –68 | 0.80 / 0.01 | +77 | 0.94 |
| 27B it | SN | 1075 | +11 | 31.0 | 0.73 | +14 | +33 | –19 | 0.34 / 0.13 | +220 | 0.94 |
| InternVL3.5 8B pt | NI | 859 | –27 | 41.9 | 0.67 | +95 | +152 | –57 | 0.76 / 0.01 | +44 | 0.78 |
| InternVL3.5 8B final | NI | 859 | +11 | 43.4 | 0.65 | +58 | +76 | –18 | 0.34 / 0.26 | +62 | 0.75 |

NI, NutriImage; SN, SNAPMe. Bias, mean (estimate – reference) without a narrator. JP – none, US – none, JP – US, larger – smaller: mean paired differences. JP<US / JP>US and L>S: proportions of images. Images with reference energy above 20 kcal (Section 2.4). InternVL3.5 8B rows: score that does not depend on how the number ends (Section 2.3(d)); 'final' denotes the released post-trained weights.

Before the run, we had expected the contrast to be absent in pt and present in it. The result was the reverse in size. On NutriImage (859 images), JP − US was −64 kcal (95% CI −67 to −61) in 4B pt and −22 kcal (−26 to −17) in 4B it, −44 (−47 to −41) and −21 (−25 to −17) kcal at 12B, and −107 (−112 to −102) and −35 (−40 to −31) kcal at 27B; the intervals of pt and it did not overlap at any size. Within images, the contrast was smaller in it than in pt by 42 kcal (paired difference; 95% CI 38 to 47) at 4B, 23 (19 to 27) at 12B, and 71 (66 to 77) at 27B on NutriImage, and by 35 (29 to 41), 24 (21 to 28), and 49 (45 to 53) kcal on SNAPMe; larger − smaller was larger in it by 143, 121, and 168 kcal on NutriImage and 136, 88, and 143 kcal on SNAPMe (all intervals excluding zero; Multimedia Appendix 1, S8, Table S8d). The contrast was smaller in it in 0.48–0.75 of images on NutriImage (4B 0.59, 12B 0.48, 27B 0.75) and 0.48–0.68 on SNAPMe; at 12B, then, the mean difference of +23 kcal arose with the contrast smaller in fewer than half of the images, so that the mean change reflects an asymmetry in the size of the changes, with large reductions in a part of the images, rather than a reduction shared by most images. In pt, the JP estimate was lower than the US estimate in 0.82, 0.66, and 0.91 of images and higher in 0.01, 0.05, and 0.02; in it, lower in 0.40, 0.38, and 0.52 and higher in 0.12, 0.15, and 0.14. Taking the mode of the distribution instead of its median gave the same signs and ordering (pt −51, −30, −81 kcal; it −21, −20, −33 kcal; Multimedia Appendix 1, S8).

Instruction following showed the opposite pattern. Larger − smaller was +59, +40, and +53 kcal in pt (higher under *larger* in 0.85, 0.69, and 0.81 of images) and +202, +161, and +221 kcal in it (0.97– 0.98), a three-to fourfold difference on NutriImage.

Accuracy without a narrator was similar in the two sets of weights (MedAPE 40–44% in pt and 46– 55% in it; ρ 0.63–0.72 and 0.68–0.71), so the differences above are not explained by a general inability of the pre-trained weights to perform the task. For the instruction-tuned weights, JP − US under scoring (−22, −21, −35 kcal) had the same sign and ordering across sizes as in the chat format (−40, −46, −54 kcal; Table 2); the absolute values depended on the format.

P3 was therefore not supported. In Gemma 3, the contrast between a Japanese and a US dietitian was already present after pre-training and was smaller after post-training, whereas the response to explicit portion instructions was several times larger after post-training.

#### SNAPMe (P4)

The same scoring was applied to SNAPMe (1,075 images), which consists of meals of participants in the United States. Two outcomes had been written before the run: a contrast of similar or greater size than on NutriImage if the narrator changes the serving assumed for a population, and less than half if it changes servings dish by dish. In pt, JP − US was −52 kcal (95% CI −55 to −50) at 4B (NutriImage −64), −34 kcal (−37 to −31) at 12B (−44), and −68 kcal (−72 to −65) at 27B (−107), that is, 82%, 77%, and 64% of the NutriImage values, with the JP estimate lower in 0.70, 0.55, and 0.80 of images and higher in 0.02, 0.06, and 0.01. In it, the contrast was −17 (−22 to −12), −9 (−13 to −6), and −19 (−23 to −16) kcal. The contrast thus remained on US meals, as P4 states, at a size between the two outcomes written beforehand. Larger − smaller was +69, +75, and +77 kcal in pt and +205, +164, and +220 kcal in it, as on NutriImage (Table 7).

#### InternVL3.5 8B (P6)

The pre-trained and the final weights of InternVL3.5 8B were scored on NutriImage after the Gemma 3 results had been seen, with the outcome written down beforehand (Section 2.3(d); Table 7; Figure 5). In the pre-trained weights, JP − US was −57 kcal (95% CI −60 to −54), with the JP estimate lower in 0.76 of images and higher in 0.01; in the final weights it was −18 kcal (−24 to −13; lower in 0.34, higher in 0.26). Larger − smaller was +44 kcal (42 to 46) before and +62 kcal (58 to 66) after the SFT and RL stages, a ratio of 1.4. Within images, the contrast was smaller in the final weights by 38 kcal (paired difference; 95% CI 32 to 44; smaller in 0.63 of images), and larger − smaller was larger by 18 kcal (14 to 21). Accuracy without a narrator was similar (MedAPE 41.9% and 43.4%; ρ 0.67 and 0.65). The line-break score, defined as for Gemma 3, gave the same signs and ordering (JP − US −53 and −29 kcal; larger − smaller +44 and +65 kcal).

Stratified by the error of the estimate without a narrator, the contrast in the pre-trained weights was −51, −51, −66, and −92 kcal across the four strata (slope −0.05, r −0.17), and in the final weights −2, −2, −8, and −95 kcal (slope −0.18, r −0.32; 140 images in the highest stratum), that is, close to uniform before post-training and confined to the overestimated images after it. The sign of the contrast in the pre-trained weights, its reduction after post-training, and the larger instruction response after post-training were as written beforehand; the size of the pre-trained contrast exceeded the −10 to −40 kcal expected, and the ratio of the instruction responses fell below the 1.5 expected (Table 1). The probability that the scored distribution placed on the 30 candidates was small (median 0.03 in the pre-trained and 0.10 in the final weights; Multimedia Appendix 1, S8), because these weights often continue the number in forms that are not on the grid (for example with decimals or above 1,500 kcal); the medians rest on the part of the distribution that the grid covers (Section 4.6). The intermediate checkpoints (Instruct, MPO) and SNAPMe were not scored. Because the ‘Pretrained’ checkpoint of InternVL3.5 already includes multimodal training on dialogue data (Section 2.3(d)), what this comparison shows is that the contrast was present before the SFT and RL stages and was reduced by them, not that it was present before any instruction-like training.

#### Contrast across strata of baseline error (description)

Images were stratified by the error of the estimate without a narrator, a condition that enters neither term of the contrast (Section 2.5), into four strata (< 0, 0–50, 50–150, > 150 kcal; Figure 6; Multimedia Appendix 1, S8). In pt, the contrast was of similar size in the three lower strata and somewhat larger in the highest: for 27B, −105, −103, −105, and −117 kcal on NutriImage and −68, −58, −63, and −84 kcal on SNAPMe. The slope against the error was between −0.05 and 0.00 in the six pt cells (r between −0.15 and 0.01), and the contrast was already −34 to −105 kcal where the model underestimated. In it, the contrast where the model underestimated was −6 to −16 kcal in all six cells. At 27B it increased with overestimation on both image sets (NutriImage: −16, −33, −39, −54 kcal, slope −0.10; SNAPMe: −14, −6, −19, −49 kcal, slope −0.06), as at 12B on NutriImage (−6, −9, −23, −37 kcal; slope −0.09), the form reported previously for commercial models in the chat format; at 4B, and at 12B on SNAPMe, it changed little across strata (slopes −0.02). After pre-training the contrast was thus close to uniform across images; after post-training it was smaller throughout and, in the larger models, depended on baseline error.

**Figure 6.**
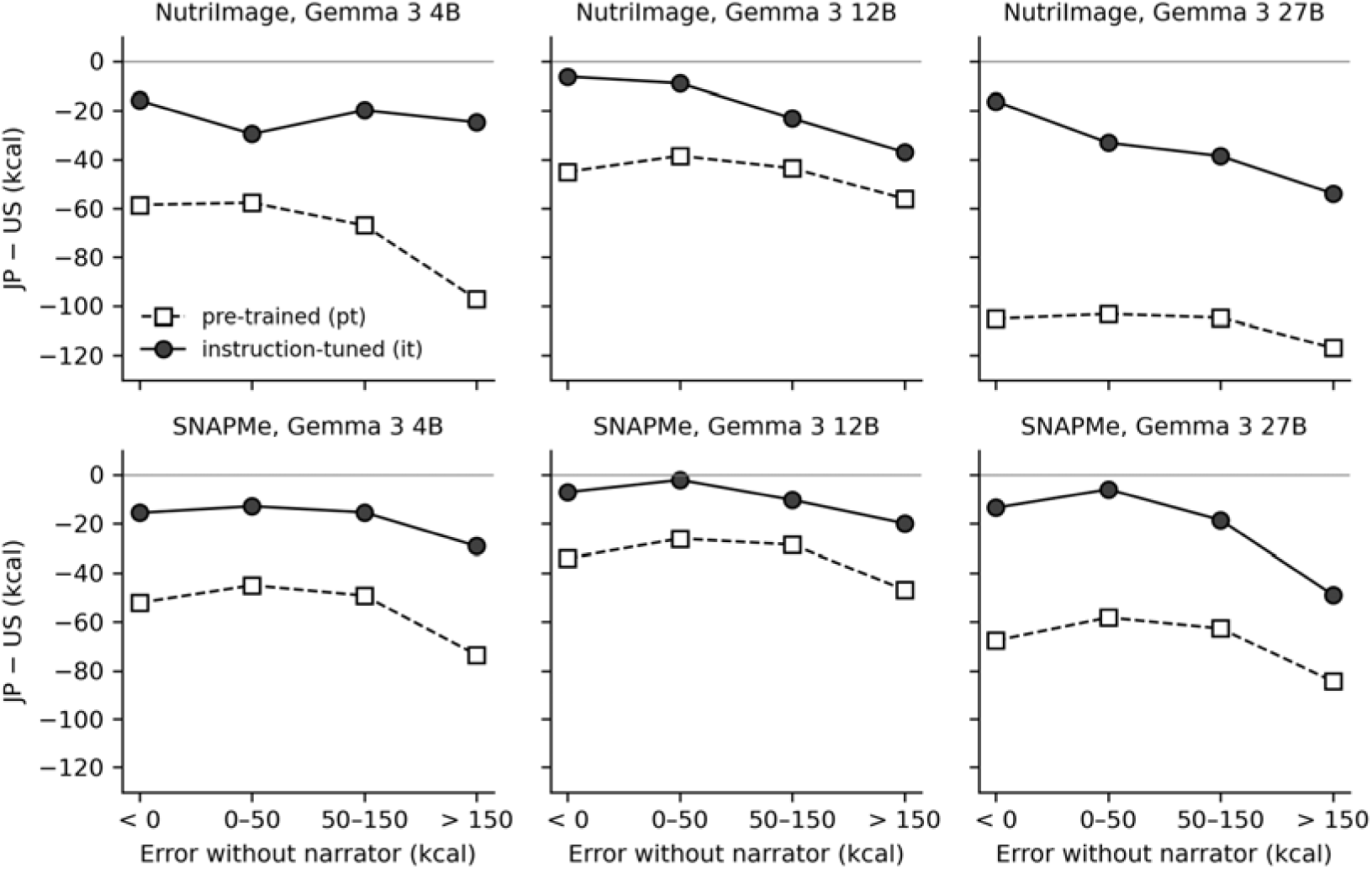
JP − US contrast by stratum of baseline error, Gemma 3 pre-trained and instruction-tuned weights. Mean paired difference JP − US within strata of the error of the estimate without a narrator (estimate − reference), which enters neither term of the contrast. Stratum sizes are given in Multimedia Appendix 1, S8.

The correlations at the image level were weak in every case (|r| ≤ 0.21). A regression of the contrast of 27B on the estimate without a narrator and on the reference value together is given in Multimedia Appendix 1, S8. We describe these forms here without interpretation.

### 3.5 Form of the contrasts across images (description)

When contrasts between conditions were stratified by the error of the estimate without a persona, which enters neither term, the Japan–US contrast was of similar size across strata on SNAPMe in all commercial models (about 0.85–0.95 of the US estimate in the models that responded). On NutriImage it was of that size where the model underestimated and larger where it overestimated in the commercial models of both years, in Gemma 3 12B and 27B, and in Qwen3-VL 32B; in Qwen3-VL 4B and 8B, InternVL3.5 8B, and Mistral Small 3.2 it was small and of similar size across strata (Table 8; Figure 7; all cells in Multimedia Appendix 1, Table S14). The contrast between the *smaller* instruction and the US persona changed little across strata. These forms suggest that the persona shift has a part that is uniform across images and, for Japanese dishes in some models, a further part confined to images that the model overestimated, that is, a step between the stratum with negative error and the others and not a gradient across all strata. The way of reading these forms was settled on the six models of the previous study and then applied unchanged to all models.

**Figure 7.**
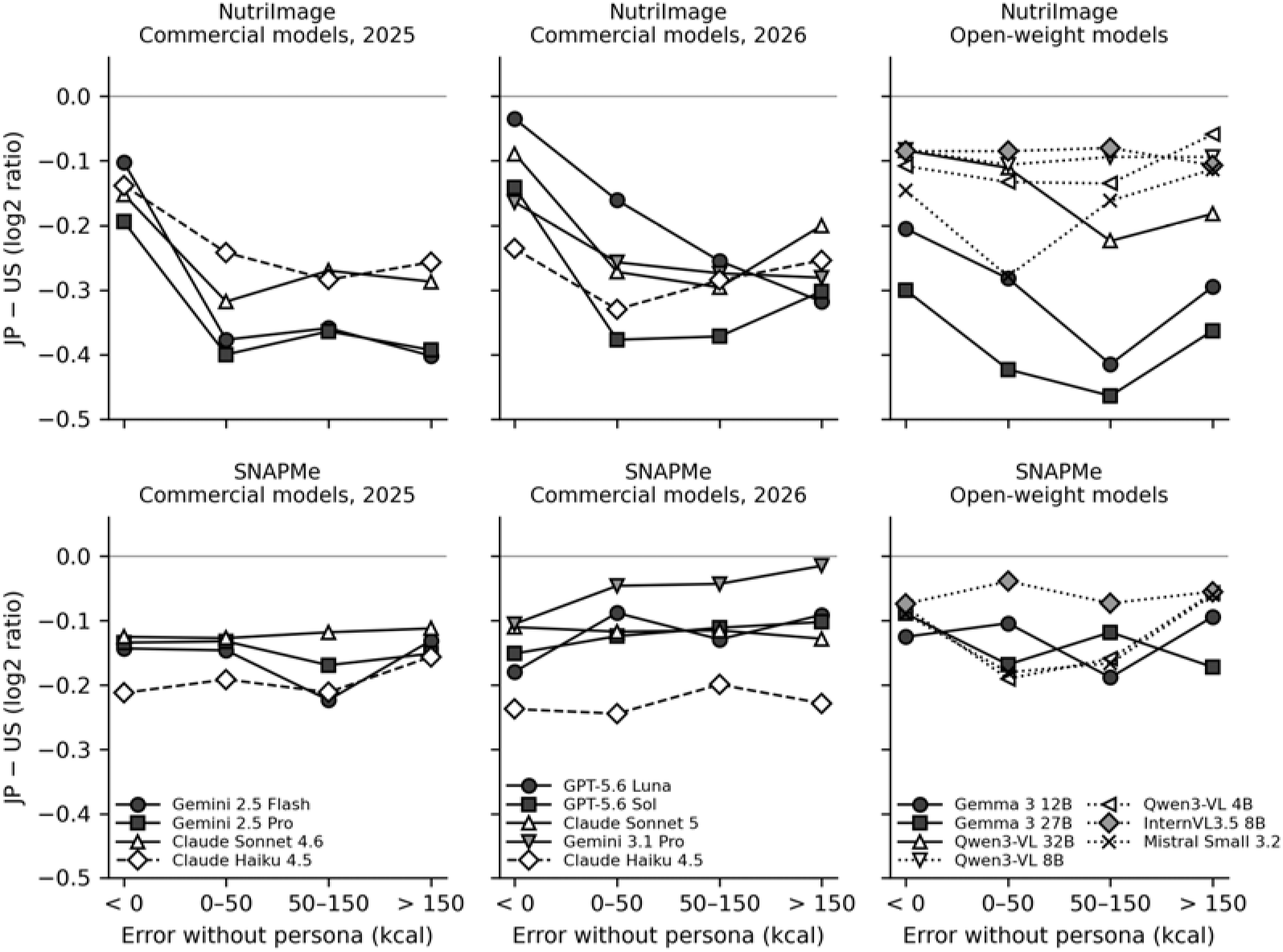
JP − US contrast by stratum of the error without a persona, chat format. Mean of log2 of the ratio of the estimate under the JP persona to that under the US persona, within strata of the error of the estimate without a persona (estimate − reference), which enters neither term. Top, NutriImage; bottom, SNAPMe. Cells in which the mean contrast did not reach −10 kcal are not drawn (NutriImage: GPT-4o, GPT-4o-mini, Gemini 3.8 Flash; SNAPMe: the same two GPT models, Qwen3-VL 8B and 32B; Gemini 3.8 Flash is omitted on both sets). Dotted lines, open-weight models without a step on NutriImage. Values and intervals: Table 8 and Multimedia Appendix 1, Table S14.

**Table 8.**
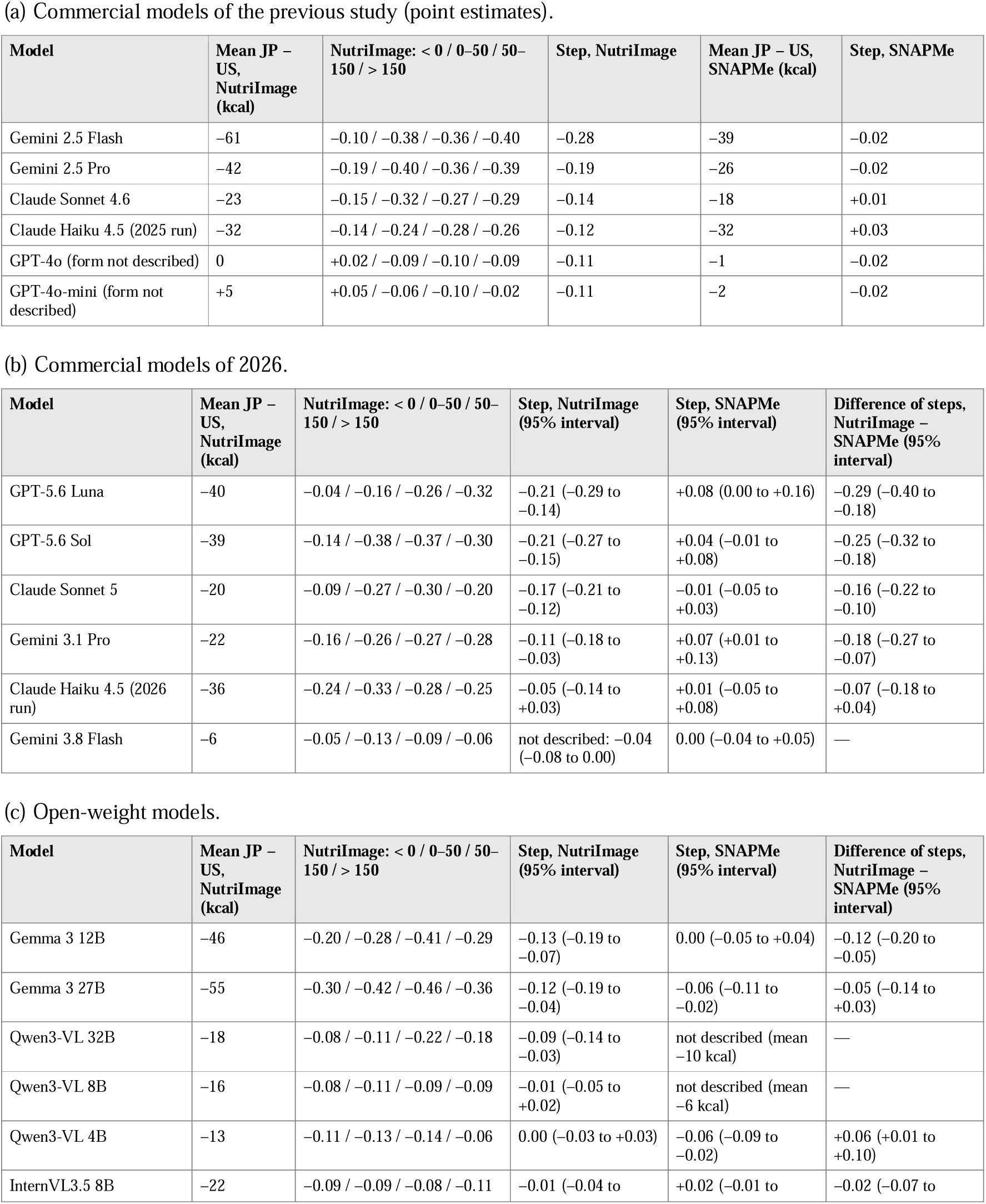
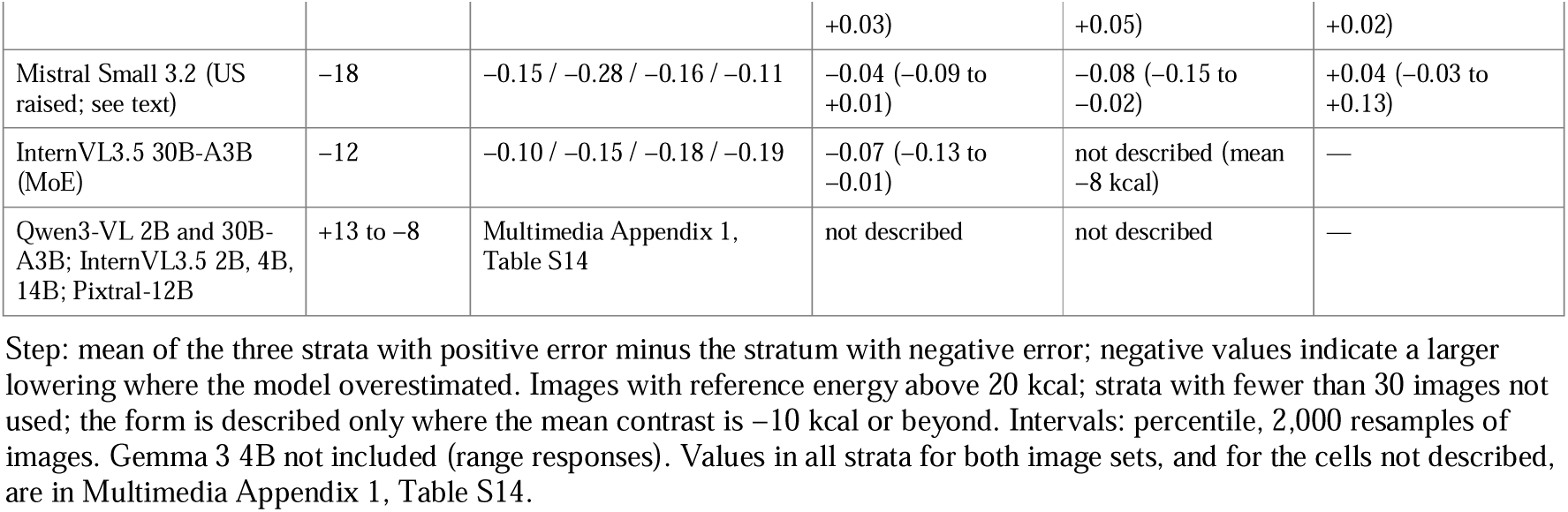
Step of the JP − US contrast across strata of the error without a persona (log2 of the ratio of estimates; chat format).

#### Step of the JP − US contrast

In the six models of the previous study the step on NutriImage (log2) was −0.28 in Gemini 2.5 Flash, −0.19 in Gemini 2.5 Pro, −0.14 in Claude Sonnet 4.6, −0.12 in Claude Haiku 4.5, and on SNAPMe between −0.02 and +0.03 in these four; in GPT-4o and GPT-4o-mini the mean contrast did not reach −10 kcal (−0.3 and +5 kcal on NutriImage), and by the rule of Section 2.5 no form is described for them (their values are shown in Table 8a for reference). In the commercial models of 2026 it was −0.21 (95% interval −0.29 to −0.14) in GPT-5.6 Luna, −0.21 (−0.27 to −0.15) in GPT-5.6 Sol, −0.17 (−0.21 to −0.12) in Claude Sonnet 5, and −0.11 (−0.18 to −0.03) in Gemini 3.1 Pro; in the 2026 run of Claude Haiku 4.5 it was −0.05, with an interval that included zero (−0.14 to +0.03), and on NutriImage Gemini 3.8 Flash hardly responded to the persona (mean JP − US, −6 kcal; on SNAPMe, −18 kcal, uniform). On SNAPMe the step was between −0.01 and +0.08, and the difference between the two image sets had an interval that excluded zero in four of the five models (Table 8b). Among the open-weight models, the step on NutriImage was −0.13 (−0.19 to −0.07) in Gemma 3 12B, −0.12 (−0.19 to −0.05) in Gemma 3 27B, and −0.09 (−0.14 to −0.03) in Qwen3-VL 32B. In Qwen3-VL 4B and 8B, InternVL3.5 8B, and Mistral Small 3.2, JP − US was negative (−13 to −22 kcal) but the step was between 0.00 and −0.04 with intervals that included zero, that is, a lowering of similar proportion in all strata (to about 0.93 of the US estimate) (Table 8c). On SNAPMe, negative steps were small or absent (Gemma 3 27B, −0.06; Qwen3-VL 4B, −0.06; Mistral Small 3.2, −0.08); the difference between the two image sets had an interval that excluded zero in Gemma 3 12B (−0.12; −0.20 to −0.05) and included it in Gemma 3 27B (−0.05; −0.14 to +0.03). The models with a step on NutriImage were those of group A in Section 3.6, together with Qwen3-VL 32B, which had lain nearer the group-A line.

#### Other measures of the same form

The proportion of images lowered showed the same form (NutriImage, from the stratum with negative error to the strata with positive error: GPT-5.6 Luna 0.38 to 0.72, GPT-5.6 Sol 0.58 to 0.77, Claude Sonnet 5 0.47 to 0.73; flat on SNAPMe). The contrast jp_home − us_home had the same form (NutriImage step −0.10 to −0.18 in the five commercial models of 2026 that responded, with the interval of the difference between image sets excluding zero in all five; Gemma 3 12B and 27B, −0.09; Qwen3-VL 32B, −0.12). Where the model underestimated, the JP − US contrast on NutriImage was small but not zero (log2 −0.04 to −0.30, about 0.97 to 0.81 of the US estimate, in the cells for which a form was described), and this was about the level of the uniform contrast on SNAPMe (about 0.9). The step of smaller − US was small (commercial 2026, −0.01 to −0.06; Gemma 3 12B 0.00, 27B +0.07; Qwen3-VL 32B −0.01); its difference from the step of JP − US had an interval excluding zero in GPT-5.6 Sol, Claude Sonnet 5, Gemma 3 12B and 27B, and Qwen3-VL 32B, and including zero in Gemini 3.1 Pro and Claude Haiku 4.5. The instruction contrast was thus nearly uniform, with a step smaller than that of the persona contrast. In GPT-5.6 Luna the *smaller* instruction raised the estimates (smaller − US, +35 and +49 kcal), in agreement with the reversed instruction following noted in Section 3.3.

#### Cells read differently or not read

In Mistral Small 3.2, JP − US was negative because the US persona raised the estimates and not because the JP persona lowered them (against *none*, JP +9 and US +28 kcal on NutriImage), so this model is not read in the same frame as the others. Gemma 3 4B is not included because of its range responses (Multimedia Appendix 1, S4), and the two mixture-of-experts models are given in Multimedia Appendix 1 (InternVL3.5 30B-A3B: mean JP − US −12 kcal, step −0.07, −0.13 to −0.01). In Qwen3-VL 2B and 30B-A3B, InternVL3.5 2B, 4B, and 14B, and Pixtral-12B the mean contrast (+13 to −8 kcal) did not reach −10 kcal, and no form was described.

#### Repeated runs

Runs of the same conditions on two occasions were available for Claude Haiku 4.5 (the run of the previous study, July 2026 for NutriImage and June 2026 for SNAPMe with the added conditions of August 2026, and that of September 2026; NutriImage, 853 images; SNAPMe, 1,063) and for Gemini 2.5 Flash (a run of July 2026 and a re-run of 15–16 August 2026, both for the previous study; NutriImage, 714 images valid in both) (Multimedia Appendix 1, S12). Without a persona, the within-image SD between runs was 33 and 45 kcal for Claude Haiku 4.5 and 70 kcal for Gemini 2.5 Flash, against between-image SDs of 145–238 kcal. Cell means were reproduced within 1–8 kcal, whereas the change of the individual image correlated between runs at only 0.34–0.57 under the descriptions that moved the estimates and at 0.33 and 0.12–0.19 under *us*; on SNAPMe, Pearson’s r for Claude Haiku 4.5 was higher (JP 0.73, smaller 0.66, us 0.67), but the rank correlations were of the same order as on NutriImage (0.42, 0.40, and 0.31). With the error of *none* taken from the same run, the US persona, whose mean shift was near zero, showed a gradient across strata (Claude Haiku 4.5, NutriImage, log2: +0.10 to −0.08), which disappeared when the error was taken from the other run; about four tenths of the JP gradient was of this kind. The second kind of apparent slope of Section 2.5, and the JP shift where the model underestimated with the error taken from the other run, are given in Multimedia Appendix 1, S12.

### 3.6 Slope of the shift against baseline bias by family, model size, precision, and vision encoder (P5)

Figure 2 plots the JP shift against baseline bias with the two groups distinguished. In group A the slope was −0.25 (n = 28, r = −0.83); the 24 commercial cells alone gave −0.24 (r = −0.81), and Gemma 3 12B and 27B lay within ±6 kcal of the commercial line on both image sets. In group B the slope was −0.09 (n = 20, r = −0.52). Resampling models with replacement (5,000 samples; each model with both of its cells), the slope was −0.25 (95% CI −0.30 to −0.16) in group A and −0.08 (−0.15 to −0.03) in group B, and their difference −0.17 (−0.25 to −0.05); the groups had been defined after inspection of the figure, so this interval describes the difference and does not test a prior hypothesis. The difference was seen on both image sets (NutriImage: −0.41 and −0.17; SNAPMe: −0.17 and −0.10). A split-half check, in which baseline bias and the shift were computed on disjoint halves of the images, gave the same coefficients as the full image sets (medians over 200 splits: group A, r = −0.82, slope −0.25; group B, r = −0.51, slope −0.09; Multimedia Appendix 1, S1). Leave-one-out prediction of the JP shift from baseline bias gave the correct sign in 28 of 28 group-A cells, with a mean absolute error of 8.6 kcal (SD of the shifts, 18 kcal).

#### Model generation

Fitted separately, the 2025 and 2026 commercial cells gave nearly the same line (2025: slope −0.25, intercept −21 kcal, r = −0.81; 2026: −0.23, −22 kcal, −0.80), although baseline bias changed between generations within a vendor (Multimedia Appendix 1, S15).

#### Model size and vision encoder

In InternVL3.5 (2B–14B) the within-family slope was −0.06; the 8B and 14B cells, with baseline biases of +37 and +14 kcal on NutriImage, lay within 3 kcal of the group-B line and 23–24 kcal from the group-A line. InternVL3.5 uses a vision encoder and tiling scheme different from those of Qwen3-VL, so the vision front end alone does not appear to account for the difference from group A. In Qwen3-VL (2B–32B) the 2B and 4B cells shifted within ±3 kcal (tie 0.6–0.8), the 8B cells lay near the group-B line, and the two 32B cells lay nearer the group-A line (residuals +7 and +4 kcal from the group-A line, −17 and −11 kcal from the group-B line); the within-family slope was −0.14. Whether the position of the 32B model reflects size or a property of that model could not be determined. The two Mistral models gave a within-family slope of −0.05; Pixtral-12B, at a baseline bias of +47 kcal on NutriImage, shifted by −1 kcal (down 0.14, up 0.13).

#### Numerical precision

For the ten Qwen3-VL cells run in both BF16 and FP8, the sign of JP − US agreed in every pair and the difference was at most 3 kcal (Multimedia Appendix 1, S2). With MLX 4-bit weights of Qwen3-VL 8B on Apple silicon, accuracy without a persona was unchanged from BF16 (NutriImage: bias +39 against +36 kcal, MedAPE 46.5% against 46.6%, ρ 0.78 against 0.80), but the JP shift differed (+3.6 against −10.8 kcal) (Multimedia Appendix 1, S3); the runtime also differed, so this is not a comparison of precision alone.

#### Placement of the description

The description was placed at the beginning of the user turn for Gemma 3 (group A) and Pixtral-12B (group B) and in the system role for all other models of both groups, so the placement does not coincide with the grouping.

#### Cells not analysed

Gemma 3 4B was excluded because of range responses (Multimedia Appendix 1, S4); the two mixture-of-experts models did not fall on either line and are reported without interpretation (Multimedia Appendix 1, S7).

### 3.7 Consequences for accuracy

Because the JP shift followed baseline bias, it moved estimates toward the reference where bias was positive and was small where it was not; the smaller shift lowered estimates irrespective of bias (Multimedia Appendix 1, Table S1). In all 15 cells with a baseline bias above +30 kcal among the 48 analysed, MedAPE was lower under the JP persona than without it (e.g., Gemini 2.5 Pro, NutriImage: 41% to 30%; GPT-5.6 Sol: 51% to 35%; Gemma 3 27B: 57% to 40%; Qwen3-VL 32B: 49% to 39%), with smaller gains in group B (Pixtral-12B: 50.4% to 48.8%). In 24 of the 26 cells with negative baseline bias, MedAPE under the persona was within 3 points of the value without it; the exceptions were Claude Haiku 4.5 on SNAPMe (36% to 39%) and GPT-4o on NutriImage (37% to 32%). Under the smaller instruction, MedAPE worsened in models with little bias (Claude Sonnet 5, SNAPMe: 30% to 41%; Gemini 3.1 Pro, SNAPMe: 31% to 50%) and improved only where baseline bias was large and positive. The larger instruction raised MedAPE wherever instruction following was large (Claude Sonnet 5, NutriImage: 41% to 77%; Gemini 3.1 Pro: 35% to 167%) and hardly at all in group B. Spearman’s ρ without a persona lay between 0.60 and 0.86 in all cells, a narrow range compared with that of baseline bias; differences in MedAPE between models were largely differences in bias.

#### Comparison with numerical calibration

With the correction determined on a random half of the images and evaluated on the other half (200 splits; Table 9, median and 2.5th–97.5th percentiles over splits), subtracting the mean bias lowered MedAPE where baseline bias was large and positive, and by less than dividing by the median ratio of estimate to reference (GPT-5.6 Luna, NutriImage: 92.8% without a persona, 46.7% after subtraction, 39.5% after division; Gemma 3 27B: 55.2%, 46.2%, 38.7%); elsewhere subtraction changed MedAPE little or raised it (Gemini 3.1 Pro, SNAPMe: 29.8% to 35.2%). Division lowered MedAPE where baseline bias was large and left it nearly unchanged elsewhere. On NutriImage, in the commercial models of 2026, MedAPE under the JP persona was lower than after subtraction of the mean bias in four models, with the interval of the within-split difference excluding zero (GPT-5.6 Sol −7.5 (−10.7 to −4.1); Gemini 3.1 Pro −5.2 (−7.4 to −2.8); Claude Haiku 4.5 −5.1 (−9.1 to −0.7); Claude Sonnet 5 −5.3 (−7.4 to −2.7) points), and not distinguishable from it in GPT-5.6 Luna and Gemini 3.8 Flash; against division, the persona was lower in Claude Sonnet 5 (−3.8 (−6.0 to −1.6)) and Gemini 3.1 Pro (−3.4 (−5.9 to −1.4)), not distinguishable in three models, and higher where baseline bias was very large (GPT-5.6 Luna, +10.4 (+3.6 to +14.9)). The same held for Gemma 3 12B and 27B and Qwen3-VL 32B against subtraction (−5.5 (−8.6 to −1.1), −7.3 (−11.1 to −3.5), −5.1 (−7.9 to −1.9)) and was not distinguishable against division. On SNAPMe, MedAPE under the JP persona was within 3 points of the value without a persona in five of the six commercial models, higher in two of them with the interval excluding zero (Gemini 3.8 Flash +2.1 (+0.4 to +3.7); Claude Haiku 4.5 +2.6 (+0.7 to +4.3)); the exception was GPT-5.6 Luna, with a large bias (49.4% to 41.4%; after division, 32.5%). In the open-weight families that responded little to the persona (Qwen3-VL 8B, InternVL3.5, Pixtral-12B), MedAPE under the persona was within the split-to-split range of that without it, and it was numerical calibration, not the persona, that lowered it where baseline bias was large (division: by 4–7 points in Qwen3-VL 8B, InternVL3.5 8B and 14B, and Pixtral-12B on NutriImage, and by 4–6 points in InternVL3.5 2B and 4B on SNAPMe, intervals excluding zero; Multimedia Appendix 1, Table S13). The *smaller* instruction raised MedAPE considerably in models with little bias (Gemini 3.1 Pro, SNAPMe: 29.8% to 50.2%); only in Pixtral-12B on NutriImage (baseline bias +45 kcal, with a response to the instruction) was it the lowest column. The split-to-split percentiles span 2–5 points for most cells, so differences of a few points between methods are within this range unless the within-split difference is reported. That the persona was as low as division, or lower, in cells with little bias is in keeping with the further part of the contrast described in Section 3.5, which lowers overestimated images more than others and so reduces dispersion as well as bias. Whether a persona lowered error thus depended on the cuisine and on the model.

**Table 9.**
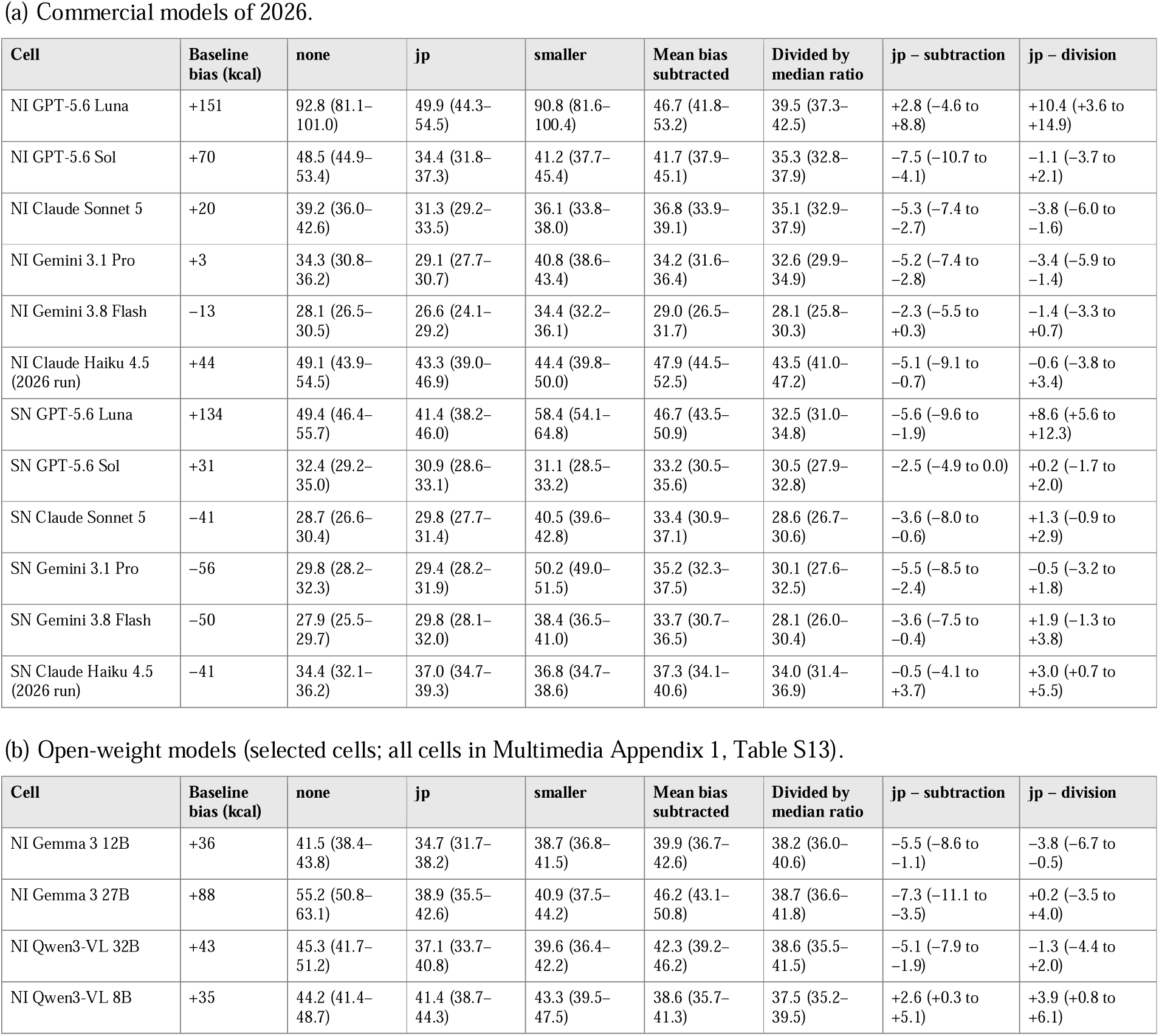
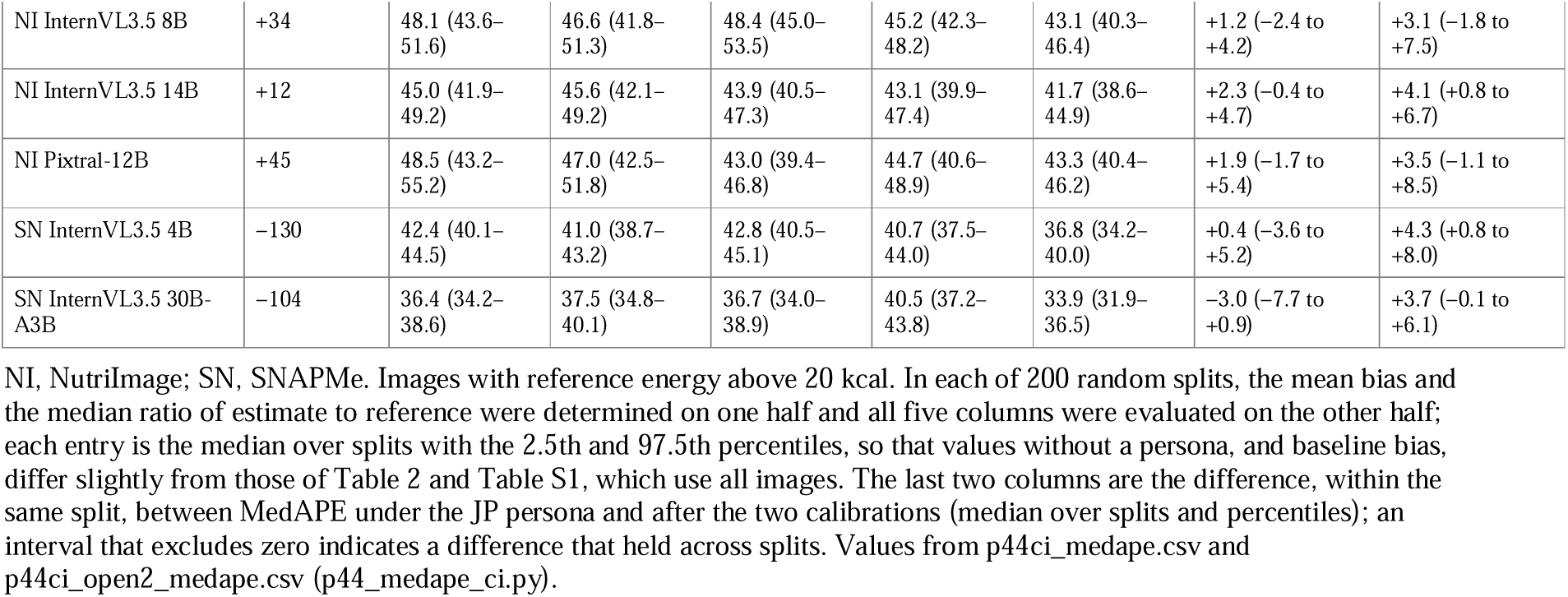
MedAPE (%) under prompt conditions and after numerical calibration determined on a random half of the images and evaluated on the other half: median over 200 random splits (2.5th–97.5th percentiles).

In the other formats, for Claude Sonnet 4.6 on NutriImage (crop experiment, full image; baseline bias +17 kcal) MedAPE was 34.5% without a persona, 35.2% after subtraction of the mean bias, and 32.9% under the JP persona. For Gemma 3 it under scoring, subtraction lowered MedAPE more than the JP narrator did (NutriImage, 27B: 50.4% to 45.2% against 50.3%; 12B: 56.7% to 41.6% against 61.7%), although in the scoring format the narrator sentence itself raises the estimates, so this comparison is unfavourable to the persona.

## Discussion

### 4.1 Principal findings

The energy estimates of Claude Sonnet 4.6, the model examined most closely, behaved as if they rested mainly on an amount associated with the identified dish and were corrected only in part by the amount visible in the photograph: where identification was retained, neither accuracy nor the persona shift depended on how much of the image remained; the dish name without the image gave estimates of similar accuracy; and photographs of trays that were nearly empty received 39–75% of the before-meal estimate in the chat format, often with a description of the meal that had been eaten. These observations are consistent with a two-stage account rather than a demonstration of it: the name-only condition supplied the reference dish name rather than the name the model itself would have produced, and the subset with retained identification was selected by a model judge after the fact; conditions that re-enter the model’s own name, or change only the name while holding the image fixed, would be needed to separate the two stages directly. Two kinds of description in the prompt moved this value in different ways. A description stating Japanese nationality lowered it without a change in how often the dish was correctly named, by an amount that was related, between models, to baseline bias and not to instruction following; an explicit instruction about amount showed the reverse dependence. Across images, the contrast between the Japanese and the US persona had a part that was uniform and, for Japanese dishes in the commercial models, Gemma 3, and Qwen3-VL 32B, a further part confined to overestimated images; where this further part was present, the persona lowered error as much as, or by a few points more than, subtracting the mean bias, and no more than dividing by a ratio (within-split intervals in Table 9); elsewhere it did not. In Gemma 3, the contrast between the Japanese and the US narrator was already present in the released pre-trained weights and was two to four times smaller after post-training, while the response to explicit instructions was two to four times larger [47,49]; in InternVL3.5 8B, run afterwards on NutriImage with the outcome written beforehand, the contrast was three times smaller and the instruction response 1.4 times larger after the SFT and RL stages, and its form across images changed in the same direction as in Gemma 3; because the InternVL3.5 ‘Pretrained’ checkpoint already includes instruction-like multimodal training, the two families place the change at different depths of training, and agree only that the last stages of post-training reduce the contrast and enlarge the instruction response. The slope of the persona shift against baseline bias, a relation between models, differed between two groups of families; within families the comparisons available (size ladders, FP8 against BF16, and InternVL3.5 against Qwen3-VL) did not account for it, with Qwen3-VL 32B as a possible exception [62–64]. The two groups were defined after inspection of Figure 2, and the difference between their slopes is described with an interval, not tested.

### 4.2 Comparison with the hypotheses

#### Consistent with H1

Most observations in Sections 3.1 and 3.2 are those expected under the two-stage hypothesis (H1): where identification was retained, neither accuracy nor the persona shift depended on how much of the image remained; the dish name without the image gave similar estimates and a similar shift; and a more than tenfold difference in the amount on the plate was read as a direction but compressed to a ratio of 0.4–0.8. That the estimate behaves as if anchored on the amount associated with the identified dish, and corrected incompletely by what is visible, is the observation on which the rest of the study turns, because it places the persona shift on the side where a name is turned into an amount; as noted in Section 4.1, the observations are consistent with this account and do not demonstrate it. The prediction on identification (P1c) needed closer examination than retention under cropping. A model judge suggested that the persona renamed many dishes, but human rating of 100 pairs showed that names were correct equally often with and without the persona and were different dishes in about one fifth, no more than between two requests without the JP persona. The larger shift in images whose names the judge found different therefore does not indicate re-identification; if the amount is retrieved with the name as the key, the wording of the key under the persona (the same food described as a Japanese dish) may matter. The rated sample was in keeping with this: a lowering in one direction was seen where the description changed under the persona and not where it was unchanged. This observation is offered as hypothesis-generating: it rests on 96 pairs rated by one author for one model, and the model judge was not valid across languages (Section 3.2). If confirmed in a larger sample, with independent blinded raters, and in other models, H2 would need to be restated: the persona would replace the description under which the amount is retrieved, more than the amount assumed for a given description. The dependence of the shift on baseline bias and its independence from instruction following (Section 3.3) are what the replacement hypothesis (H2) predicts, and P4 and P5 were consistent with the data, with the reservations noted for Qwen3-VL 32B.

#### Not supported

We had expected the persona channel to be introduced by post-training, in which role descriptions are common [60,61]. In Gemma 3 the result was the reverse: the contrast was two to four times larger before post-training. The expectation we favoured before the run was therefore wrong.

For InternVL3.5 8B, run afterwards, we wrote down the Gemma 3 pattern as the expectation, and its sign and ordering were reproduced, although the sizes we expected were not (Table 1, P6); the family difference in Section 3.6 cannot be attributed simply to the presence or absence of post-training.

#### What in the description carries the effect

The persona texts of the chat format mention standard single-serving portion sizes, so the lowering might be read as a response to the mention of amount. Two observations argue against this. The mention was not sufficient: the US descriptions use the same words and did not lower the estimates, and *smaller* and *larger*, which mention amount most directly, showed a different dependence (Section 3.3). Nor was it necessary: the persona text of the crop and name-only experiments describes knowledge of Japanese cuisine, ingredients, and cooking methods without any reference to amount, and the JP shift of Claude Sonnet 4.6 was −17 kcal, close to the −22 kcal of the same model in the chat format on the same images. The occupation was likewise neither necessary (the homemaker description lowered estimates by a similar amount) nor sufficient (the dietitian without nationality did not). What the effective descriptions share is the nationality. This reading has a limit that should be stated with it: under the Japanese persona text of the crop experiments the model answered in Japanese, so in those experiments the persona and the language of the output are not separated (Section 4.6). In the chat format, four of the six commercial models of 2026 and the open-weight models answered in English under the Japanese persona, whereas GPT-5.6 Sol answered in Japanese (Section 4.6); the shift and the contrast were present in the former as well, and the likelihood-scoring format involves no generation. An effect of output language alone therefore does not seem sufficient to explain the results, but its contribution in GPT-5.6 Sol and Luna and in the crop and name-only experiments remains open.

#### Not predicted

Three observations share the feature of a lowering that does not depend on baseline error: the roughly constant lowering added at the 25% crop, where most dishes were no longer identified (intercept −46 against −15 kcal, with the same slope); the nearly uniform contrast in pre-trained Gemma 3; and the part of the JP − US contrast in the chat format that was uniform across images, which was seen in every family that responded to the persona and on both image sets (about 0.9 of the US estimate; Section 3.5). We place them side by side without claiming that they are the same thing. (The intercept of the cell-level regression in group A is in keeping with the third; Section 3.3.) For the 25% crop, the added lowering coincided with a higher proportion of names judged different under the persona (Section 3.2), but the judge was not valid across languages, and the nature of this coincidence is not established. One possible reading is that, when no dish-specific amount can be retrieved, or before post-training has made the adjustment conditional, the persona changes a default amount; this was not tested. The reading of the contrast as a uniform part with, for Japanese dishes in some models, a further part that depends on baseline error was formed after the results of the six earlier models had been seen, and goes no further than a suggestion.

### 4.3 Relation to previous work

#### Our previous studies

Four statements of the previous study [1] were reproduced here in 26 models and are not developed further: the lowering under the Japanese persona was in one direction; the nationality in the description, and not the occupation or the mention of portions, carried it; its effect on accuracy depended on the sign of baseline bias; and a persona is not recommended in general as a means of calibration. Two of its figures need correction. In its Figure 3, with six models, baseline bias and instruction following varied together by vendor (both small in the OpenAI models and both large in the Google models), so that a relation of the persona shift to the one could not be distinguished from a relation to the other (across its 12 cells, baseline bias and larger − smaller correlated at r = 0.60, and the JP shift at −0.81 with the former and −0.73 with the latter; computed from the per-image outputs of that study with its own reference values); in the present set the two were only weakly correlated (r = 0.20), and the two shifts separated (Section 3.3). In its Figure 4, the estimate without a persona entered both the horizontal axis (error without a persona) and the vertical axis (persona − none), and the slopes included the two apparent components described in Section 2.5; about four tenths of the JP gradient was of the first kind (Section 3.5; Multimedia Appendix 1, S12), and the lowering where the model underestimated, reported there as close to zero, was small but not zero. Its third conclusion, that the persona acts as a conditional correction and the instruction as a nearly constant offset, held, with contrasts that are free of these components, for Japanese dishes in the commercial models and Gemma 3; on US meals the persona contrast, too, was uniform. The present set also showed that the relation of the shift to baseline bias is a relation between models (Section 3.3). The expectation of the first report [13], that an object of known size in the photograph would reduce the error in amount, could not be examined properly in ACETADA: every photograph includes a marker for scale, but its dimensions were not given to the models, and the after-meal estimates reflected in part the meal that had been eaten (Section 3.1).

That VLMs may answer from prior knowledge in preference to what is in the image has been shown with counterfactual images and with ‘blind’ baselines in which the image is withheld [19–23,40,41]. Energy estimation is a case in which this is perhaps to be expected: a photograph has no absolute scale, and the amount has to come from somewhere other than the pixels. The name-only condition, which matched the image condition in accuracy, and the before/after pairs place a number on this for one task. In the evaluation that introduced ACETADA, supplying the list of food items and other context as text reduced the error of several models more consistently than reasoning prompts did, and an expert persona without nationality gave no evident gain on average [11]; both observations are in keeping with an estimate that is driven by what the food is named to be, and with the small shift we observed under the dietitian persona without nationality. In the study that proposed a model trained on paired before-and after-meal images, a commercial VLM asked for the weight consumed, either directly or as the difference of two estimates, showed a percentage error of about 39% on ACETADA, and in the authors’ own model text input alone gave a lower error than image input alone [24]. The present observations are in keeping with both, and describe for general-purpose VLMs how the difference comes to fail: the after-meal estimate was compressed toward the value given to the dish before the meal. Nationality and cultural prompting is known to move the values and opinions expressed by language models [25–29,42,43] and subjective numerical judgements [44–46]; the present results extend this to an estimate with a reference value and indicate the stage at which it enters. Work on persona directions in activation space and on the relation between pre-trained and post-trained models suggests that changes induced by training and by prompting can pass through shared representations [47–49]; our finding that a nationality contrast exists before post-training is compatible with this view but does not test it. In human observers, judgements of portion size depend on the reference portion the observer holds [30,31]; the behaviour of the models resembles this, without implying a common process. Finally, Gemma 3 lay on the line of the commercial models, including Gemini, while three other open-weight families did not; the difference therefore does not seem to be one of open against closed models [33].

#### Multi-model comparisons of prompts

Two recent studies compared many models under several prompts on Nutrition5k. Vedovelli et al. [66], with 40 vision–language models, found that model architecture accounted for most of the variance in accuracy and that prompt strategies had no significant effect after correction; Sterling et al. [67] benchmarked ten approaches for accuracy and cost. Both measured accuracy under prompts intended to improve the estimate, not the direction of the change that a description produces. The present results do not contradict them: the persona shift was small against the differences between models (Section 3.7). They add that a description can move the estimate in one direction across models even where its effect on accuracy is small.

### 4.4 Remaining hypotheses and experiments that would separate them

#### Origin of the family difference

The smaller slope in Qwen3-VL, InternVL3.5, and Mistral may originate in the pre-training corpus or in the degree to which post-training attenuates the contrast. Scoring pre-trained and instruction-tuned weights of a second family in the format of Section 2.3(d), a text-only probe of the amounts associated with dish names under each narrator, and steering along a persona direction would separate these. *Effect of post-training.* In Gemma 3, post-training increased the response to explicit instructions, and possibly to the visible amount (for before/after pairs, direction and correlation with the remaining fraction were somewhat higher in it than in pt with either the median or the mode of the scored distribution, but with the mode the ratios were alike, 0.75 and 0.73, so the evidence for this is weak) and reduced, and in the larger models made conditional, the response to the narrator. Whether this is a general pattern could be examined in a family that releases intermediate checkpoints. *Output language.* A condition with the Japanese persona and an instruction to answer in English, and its converse, would separate persona from language in the crop and name-only experiments. *Internal representations.* The locus described here is functional; whether the distributions under the two narrators diverge at particular layers has not been examined.

### 4.5 Potential clinical implications

#### Intended setting

The setting we have in mind is a small or medium-sized community hospital without a dedicated research budget or follow-up system, where photographs taken when meals are served and cleared are used to support the recording of intake by inpatients and dietary counselling. The estimate would serve as a first draft for the dietitian’s own visual assessment, and the judgement would remain with the dietitian.

#### What the findings imply for operation

Several points follow from the results. Because the estimate is taken from the identified dish, a step in which the identified dish is shown and can be corrected should be part of the design; when identification fails, accuracy falls and the persona behaves differently. The ratio of estimate to reference should be measured on reference meals of the facility, for which menu and amounts are known, and the estimates divided by its median: in the comparisons of Section 3.7 this lowered error where bias was large, whereas subtracting the mean bias did so less or not at all.

Whether a persona lowered error depended on the cuisine and on the model: for Japanese dishes, in the models that responded to it, the JP persona did as well as this division or better; on US meals it did not help or made error slightly worse; and in the open-weight families that can be run within a facility and responded little to the persona, only the division helped. A persona is therefore not proposed here as a general means of calibration; the finding concerns how the estimate is formed, and if a persona is used, its effect should be checked on reference meals of the facility with the model that is deployed.

Statements about amount should be kept out of the operational prompt, since they lowered or raised estimates irrespective of bias. Calibration should be done on the weights that are deployed, because the persona response changed under 4-bit weights although accuracy without a persona did not. A larger model of the same family should not be expected to respond better to the persona, with Qwen3-VL 32B as a possible exception. And intake should not be obtained by subtracting the after-meal from the before-meal estimate: in ACETADA this difference correlated with the energy consumed at only 0.12–0.42 and was too small by 330–590 kcal against a median consumption of 623 kcal, because trays on which 5% or less of the energy remained were given 39–75% of the before-meal estimate in the chat format; with a prompt that asks for the content of ‘this meal’, an after-meal photograph tends to be read as the meal that was eaten, and a prompt for such photographs would have to ask explicitly for what remains, which was not tried here.

#### Size of the errors

MedAPE per meal was 30–50%, and the systematic shift due to the persona was 20–50 kcal per meal; as it has the same direction at every meal, it amounts to 60–150 kcal over three meals, or about 3–9% of a daily intake of 1,600–2,000 kcal. This is not sufficient for the assessment of single meals; the conceivable use is to follow the average over several days and to give a starting point for the interview. The error of visual estimation by trained staff varies widely with conditions [3,30,50,51], and we do not compare the two here.

#### Safety

A systematic error that is constant within a person affects the following of change over time less than it affects judgements against a threshold, for which correction would be needed. Because a single sentence in the prompt moved estimates by tens of kilocalories, prompts should be under change control in the same way as other settings of a clinical system.

#### Path to validation

The present study is a retrospective application to existing image sets. The next steps would be the estimation of intake in data with weighed before- and after-meal amounts by a method other than subtraction of two independent estimates, for example a model trained on the difference between paired images [24], and then comparison with the weighing method within a facility.

### 4.6 Limitations

The after-meal photographs of ACETADA were given the same task prompt as the before-meal photographs; a prompt asking for the food that remains was not run, so that the reading of visible amount and the interpretation of the question are not separated in the before/after pairs. The JP and US descriptions differ in more than nationality: each names the cuisine with which the dietitian is familiar, and in the crop and name-only experiments the JP text also contains Japanese script; what is called the nationality contrast here is therefore the contrast between a Japanese and a US context as described, from which occupation and the mention of portions, but not cuisine, were separated by the control conditions. The comparison of prompt conditions with numerical calibration was made on random halves within each image set and is an internal check; whether a ratio measured on the meals of one facility holds at another was not examined. The crop and name-only experiments used one model and a persona text different from that of the chat format; this text contains the Japanese term for the profession in Japanese script, and under it the model wrote the dish name in Japanese in 99% and the basis in 95% of responses, so that persona and output language are not separated in these experiments. In the chat format this confound is limited to some models. Under *jp* the basis was written in Japanese in 0.3% or less of responses by Qwen3-VL 8B, and in the six commercial models of 2026 the proportion ranged from 0% (Gemini 3.1 Pro) to 98–99% (GPT-5.6 Sol), with 24–41% in GPT-5.6 Luna, 7–15% in Gemini 3.8 Flash, and under 1% in Claude Sonnet 5 and Claude Haiku 4.5 (2–6% with at least one Japanese word in an otherwise English basis); under *jp_home* the proportions were lower in the same order (72–79% in GPT-5.6 Sol, 16% or less elsewhere), and under the other six conditions 0.3% or less. The values were within a few points in the two runs (basis_jp_rate_run1_run2.csv). The contrast was present in models whose bases were in English (Claude Sonnet 5, step −0.17; Gemini 3.1 Pro, −0.11) as in those whose bases were in Japanese (GPT-5.6 Sol, −0.21), so that the language of the output alone does not seem to account for it; for GPT-5.6 Sol and Luna, however, persona and output language are not separated in the chat format either. The scoring format involves no generation, although its Japanese narrator sentence also contains the term in Japanese script. The comparison of pre-trained and post-trained weights was made in two families, Gemma 3 at three sizes on both image sets and InternVL3.5 at one size (8B) on NutriImage only, and the origin of the family difference was not identified; for InternVL3.5 the 30 candidates carried a small part of the probability of the scored distribution (median 0.03 before post-training), so that its medians describe the part of the distribution that the candidate grid covers; ‘pre-trained’ refers to the weights as released, whose training data may include instruction-like or distilled material, and the weights were obtained from a mirror (unsloth), described by its distributor as an unmodified copy of the official release; their identity with the official release was not verified. The ‘pre-trained’ checkpoints of the two families are not equivalent: Gemma 3 pt precedes post-training of the language model, whereas the InternVL3.5 ‘Pretrained’ checkpoint is initialized from a Qwen3 language model of unstated kind and has already been trained on multimodal dialogue data, so that the comparison in InternVL3.5 concerns the SFT and RL stages only (Section 2.3(d)). Both image sets, their reference values, and our previous studies are public, so that images, dish names, or reference values may have been present in the training data of the models examined; this would bear most directly on the name-only condition and on the pre-trained probe, and it could not be excluded, because no unpublished or newly composed images were used. The model judge was not valid for comparing names across languages (κ = 0.33 against human rating), so the strata of Section 3.2 that rest on it are descriptive only; the human rating was made by one author on 100 pairs; no repeated run without a persona was available, and the none–us comparison served as the reference for variation between requests. The same-dish pairs were few (20, of which 12 with an apparent difference), and whether the difference was apparent was judged by one author, who had seen the aggregate results.

The forms across images (Section 3.5) rest on one request per image. Repeated runs were available only for Claude Haiku 4.5 (both image sets) and Gemini 2.5 Flash (NutriImage); in these, cell means differed between runs by 1–8 kcal, whereas the change of the individual image correlated between runs at only 0.34–0.49 for the persona and the *smaller* instruction on NutriImage (on SNAPMe, 0.66– 0.73 by Pearson’s r and 0.40–0.42 by rank), so that cell means are stable and the shift of a single image is much less so. The 2025 run of Gemini 2.5 Flash contained null responses that were not at random, and only images valid in both runs were used. The reading of the contrast as a uniform part and a further part for Japanese dishes was formed after the results of the six models of the previous study had been seen; the way of taking the contrasts was settled there and applied to the other models without change. No intervals were computed for the steps of those six models. The form was not described for cells in which the mean contrast did not reach −10 kcal. NutriImage and SNAPMe differ not only in cuisine but also in how the photographs were taken, in the composition of a meal, and in the source of the reference values [56–59], so the difference in step between them cannot be attributed to the cuisine alone. Associations at the level of individual images were weak (|r| ≤ 0.21 for the Gemma 3 contrasts and ≤ 0.27 for the shifts of Claude Sonnet 4.6), and the descriptions of form rest on stratum means. Dependence on baseline error was distinguished from dependence on the size of the estimate for Gemma 3 27B it on NutriImage but not on SNAPMe, and for Claude Sonnet 4.6 with the full image and with the dish name only but not at the 25% crop, where the coefficient of the estimate (−0.18) exceeded that of the reference (+0.07) (regressions on the estimate under the US persona and the reference value together; Multimedia Appendix 1, S8).

The cells of the regressions are not independent (13 models in group A and 10 in group B), and the relation of the shift to baseline bias held between models but not within models across image sets. The narrator format differs from the chat format; the direction of the contrast agreed between them, but the absolute values depended on the format, and in the narrator format only the contrast between the two narrators, not the shift from no narrator, can be interpreted. The distributions of the pre-trained weights were flat, and their median is affected by the coarseness of the candidate grid. The commercial models were run at the vendors’ default temperature with one repetition, because two vendors’ interfaces did not accept a temperature of zero; the 2025 commercial results were obtained with a different output limit and a larger SNAPMe set; and extended thinking could not be fully disabled for the two Gemini models. Each family was represented by one generation and two to five models; the two groups were defined by inspection, and the difference between their slopes was not tested. The 4-bit comparison also differed in runtime and is not a comparison of precision alone. The two mixture-of-experts models were not interpreted. Reference values carry their own uncertainty, those of SNAPMe being self-reported, and the ACETADA images were reduced in size. Only energy was analysed.

### 4.7 Conclusions

In 26 commercial and open-weight VLMs examined under one protocol, energy estimates from food photographs behaved, in the model examined most closely, as if anchored on an amount associated with the identified dish and corrected only in part by the visible amount; the difference between before- and after-meal estimates did not give the energy consumed. A Japanese-dietitian persona lowered the estimates without a change in how often the dish was correctly identified, through a route distinct from that of explicit instructions about amount. Its contrast with a US persona was uniform across images of US meals and, in some models, larger for overestimated images of Japanese dishes; only in the latter case did the persona lower error as much as numerical calibration by subtraction, and never more than calibration by a ratio. In Gemma 3, the contrast was already present after pre-training, and in InternVL3.5 8B on Japanese dishes it was present before the SFT and RL stages; in both, the later stages of training were associated with a stronger response to explicit instructions and with a smaller contrast. These findings are at the level of behaviour and remain to be examined in the internal representations of the models.

## Supporting information

ultimedia Appendix 2. Prompt texts

Multimedia Appendix 3. Description of the reproducibility package

## Acknowledgments

Claude (Sonnet 4.6, Opus 4.8, and Fable 5.1; Anthropic, San Francisco, CA, USA) was used, under the authors’ direction, to draft and refactor analysis code, cross-check statistical outputs, and edit the English text. All analyses were independently re-executed and verified by the authors; no conclusion rests on an unverified LLM-generated analysis, and all study design decisions and interpretations are the authors’ own. Models of the same vendor were among those evaluated (Claude Haiku 4.5, Claude Sonnet 4.6, Claude Sonnet 5), and Claude Haiku 4.5 served as the judge of dish-name agreement; its agreement with human rating is given in Section 2.3(b).

## Data Availability

NutriImage is archived on Zenodo (concept doi:10.5281/zenodo.21944881; version 1.1, used in this study, doi:10.5281/zenodo.21961032). In that record, the reference values calculated by dietitians (CSV) and a README documenting provenance are openly available; the 887 images from which text was removed are available under restricted access for academic research through the access-request function of the repository, and their redistribution is not permitted. SNAPMe is openly available in Ag Data Commons (doi:10.15482/USDA.ADC/1528346; accessed 30 August 2026) and is described by Larke et al. [32]; ACETADA is available from its authors for non-commercial research. Per-cell tables, per-image outputs of the 26 models, prompts, aggregation scripts, the dated records of the expectations written before the runs of Sections 2.3(c)–(e), and the script that reconstructs served energy for ACETADA will be deposited on Zenodo as a reproducibility package (doi:10.5281/zenodo.22917651) and are described in Multimedia Appendix 3. That the labels of ACETADA refer to the amounts consumed is stated by its authors [11]; the reconstruction of served and remaining energy from these labels is the authors’ calculation.

## Authors’ Contributions

Conceptualization: SN. Methodology: SN. Software: SN. Validation: SN, AY. Formal analysis: SN. Investigation: SN. Resources: SN. Data curation: SN. Writing – original draft: SN. Writing – review and editing: SN, AY. Visualization: SN. Supervision: AY. Project administration: SN. All authors have read and agreed to the submitted version of the manuscript.

## Abbreviations

CI: confidence interval; it instruction-tuned
JP: condition with the Japanese-dietitian persona
MedAPE: median absolute percentage error
pt: pre-trained
US: United States (also the condition with the US-dietitian persona)
VLM: vision–language model

## Multimedia Appendices

Multimedia Appendix 1: Supplementary methods and results. Multimedia Appendix 2: Prompt texts. Multimedia Appendix 3: Description of the reproducibility package (Zenodo).

