## Supplemental Data 1 for "Effects of Dish Name, Visible Amount, Persona, and Explicit Instruction on Energy Estimates From Food Photographs by 26 Vision–Language Models: Comparative Evaluation Study"

Multimedia Appendix 1. Supplementary material to the article. Sections S1–S15 and Tables S1–S14 are cited from the main text as 'Multimedia Appendix 1'; tables and figures of the main text are cited here by their numbers in the current revision of the main text.

### **S1. Split-half check of the regression of persona shift on baseline bias**

Each image set was split at random into halves 200 times, with the same split for all cells of an image set; baseline bias was computed on one half and the JP shift on the other, and the cell-level regression was repeated. Unified reference values, images with reference energy above zero; the 48 analysed cells. For the NutriImage cells of the commercial models of the previous study, whose outputs are keyed by dish name, 880 of the 887 images could be matched to the image numbers through their reference values and were used.

| **Cells** | **r, all images** | **Slope, all images** | **r, split halves: median (2.5th to 97.5th percentile)** | **Slope, split halves** | **Intercept, split halves (kcal)** |
| --- | --- | --- | --- | --- | --- |
| commercial 2025 (12) | −0.81 | −0.25 | −0.81 (−0.86 to −0.72) | −0.25 (−0.29 to −0.20) | −22 (−24 to −20) |
| commercial 2026 (12) | −0.80 | −0.23 | −0.79 (−0.84 to −0.74) | −0.23 (−0.26 to −0.21) | −22 (−25 to −20) |
| commercial, both (24) | −0.80 | −0.24 | −0.80 (−0.84 to −0.74) | −0.24 (−0.26 to −0.21) | −22 (−24 to −20) |
| group A: commercial and Gemma 3 12B/27B (28) | −0.82 | −0.25 | −0.82 (−0.86 to −0.77) | −0.25 (−0.27 to −0.22) | −22 (−24 to −20) |
| Qwen3−VL dense (8) | −0.57 | −0.14 | −0.56 (−0.64 to −0.50) | −0.14 (−0.18 to −0.11) | −8 (−9 to −7) |
| InternVL3.5 dense (8) | −0.54 | −0.06 | −0.53 (−0.64 to −0.40) | −0.06 (−0.07 to −0.04) | −3 (−4 to −2) |
| Mistral (4) | −0.39 | −0.05 | −0.39 (−0.56 to −0.23) | −0.05 (−0.08 to −0.03) | 0 (−1 to 2) |
| group B: Qwen3−VL, InternVL3.5, Mistral (20) | −0.52 | −0.09 | −0.51 (−0.57 to −0.46) | −0.09 (−0.11 to −0.07) | −5 (−6 to −4) |
| all cells (48) | −0.77 | −0.24 | −0.77 (−0.80 to −0.72) | −0.24 (−0.27 to −0.21) | −17 (−19 to −15) |

The coefficients obtained on disjoint halves agree with those on all images, so that the relation of the JP shift to baseline bias is not produced by the estimate without a persona entering both. Script: s1_s5.py; values: s1_split_half.csv.

### **S2. BF16 and FP8 weights (Qwen3-VL)**

The five Qwen3-VL models were run with BF16 and with the official FP8 weights on both image sets (ten pairs of cells; unified reference values, kcal).

| **Model** | **Set** | **Baseline bias, BF16 / FP8** | **JP shift, BF16 / FP8** | **JP − US, BF16 / FP8** | **Larger − smaller, BF16 / FP8** | **MedAPE without a persona (%), BF16 / FP8** | **Images with the same estimate without a persona** |
| --- | --- | --- | --- | --- | --- | --- | --- |
| 2B | NI | +1.8 / +10.5 | +1.3 / +3.2 | +2.8 / +4.9 | +3.6 / +3.7 | 44.1 / 44.8 | 0.83 |
| 4B | NI | −19.6 / −16.5 | −2.9 / −6.6 | −13.1 / −11.3 | +21.9 / +20.8 | 38.1 / 37.4 | 0.86 |
| 8B | NI | +36.2 / +36.2 | −10.8 / −8.1 | −15.4 / −13.2 | +44.2 / +38.4 | 46.6 / 46.9 | 0.89 |
| 30B−A3B | NI | +17.0 / +18.0 | +11.9 / +4.7 | +12.4 / +9.2 | +16.1 / +15.8 | 36.5 / 36.9 | 0.80 |
| 32B | NI | +44.2 / +42.6 | −25.9 / −25.3 | −17.6 / −17.3 | +120.3 / +116.9 | 48.6 / 47.7 | 0.91 |
| 2B | SN | −63.7 / −56.5 | −1.0 / +1.3 | +2.4 / +3.1 | +3.3 / +2.2 | 39.6 / 39.1 | 0.82 |
| 4B | SN | −76.7 / −77.6 | −2.0 / −1.7 | −12.6 / −11.0 | +18.5 / +16.9 | 36.0 / 35.9 | 0.89 |
| 8B | SN | −16.3 / −16.1 | +8.4 / +7.8 | −4.9 / −6.0 | +32.0 / +25.9 | 35.0 / 35.3 | 0.87 |
| 30B−A3B | SN | −32.5 / −32.8 | −5.8 / −9.1 | −1.6 / −4.1 | +19.7 / +21.7 | 31.3 / 31.2 | 0.78 |
| 32B | SN | −17.0 / −16.1 | −14.2 / −14.3 | −9.2 / −10.3 | +120.0 / +116.1 | 32.5 / 32.6 | 0.92 |

The sign of JP − US agreed in 10 of 10 pairs, and the largest difference in JP − US was 3.2 kcal (30B-A3B, NutriImage). Differences in the JP shift (up to 7 kcal) and in baseline bias (up to 9 kcal) were larger, because the estimate without a persona, which enters both, differed between the two sets of weights in 8–22% of images. Script: s2_fp8.py; values: s2_bf16_fp8.csv.

Since the shifts of Qwen3-VL were small, this shows that the values did not change with FP8 in this family and does not bear on the slope of group A (from the main text, Section 3.6).

### **S3. MLX 4-bit weights on Apple silicon (Qwen3-VL 2B, 4B, 8B)**

Qwen3-VL 2B, 4B, and 8B were run with MLX 4-bit weights (mlx-community/Qwen3-VL-{2B,4B,8B}-Instruct-4bit; mlx-vlm 0.6.17; temperature 0) on an Apple M4 on the same image files and eight conditions as the H100 runs. Accuracy without a persona was close to that of BF16 in all three sizes (MedAPE within 2 points, ρ within 0.02), whereas the response to the persona was not the same: the JP − US contrast, negative in BF16 at 4B and 8B, was smaller at 4B and near zero or reversed at 8B under 4-bit weights, and the JP shift at 8B changed sign on both image sets; instruction following (larger − smaller) was retained in direction at all sizes but was smaller at 8B. The proportion of estimates on the 50 kcal grid also differed at 2B and 4B (Table S3). Values: mac_4bit_cells_doc01.csv; the same definitions as the master table.

| **Model, set** | **n** | **none: bias / MedAPE / ρ, 4-bit** | **none: bias / MedAPE / ρ, BF16** | **JP shift, 4-bit / BF16** | **JP − US, 4-bit / BF16** | **Larger − smaller, 4-bit / BF16** | **On the 50 kcal grid (none), 4-bit / BF16** |
| --- | --- | --- | --- | --- | --- | --- | --- |
| 2B, NI | 887 | +19 / 43.2 / 0.70 | +2 / 44.1 / 0.71 | −19.4 / +1.3 | −0.0 / +2.8 | +4.1 / +3.7 | 0.79 / 0.99 |
| 2B, SN | 1134 | −37 / 38.1 / 0.71 | −64 / 39.5 / 0.73 | −17.2 / −1.1 | −2.6 / +2.5 | +1.6 / +3.3 | 0.90 / 0.99 |
| 4B, NI | 887 | +1 / 37.0 / 0.76 | −20 / 38.1 / 0.77 | −9.1 / −2.9 | −8.1 / −13.1 | +18.1 / +21.9 | 0.77 / 0.96 |
| 4B, SN | 1134 | −52 / 36.1 / 0.78 | −77 / 36.0 / 0.77 | −18.1 / −2.0 | −11.9 / −12.6 | +15.7 / +18.5 | 0.82 / 0.96 |
| 8B, NI | 887 | +39 / 46.9 / 0.78 | +36 / 46.6 / 0.77 | +3.5 / −10.8 | −3.2 / −15.4 | +29.7 / +44.2 | 0.98 / 0.99 |
| 8B, SN | 1134 | +1 / 36.3 / 0.78 | −16 / 35.0 / 0.78 | +18.7 / +8.4 | +1.0 / −4.9 | +20.3 / +32.0 | 0.96 / 0.96 |

Table S3. MLX 4-bit against BF16 (vLLM, H100). n: images with a reference value and an estimate without a persona (4-bit); BF16 n is that of the master table (885–1131). The runtime (MLX) and image preprocessing differ from the H100 runs, so the comparison is not one of numerical precision alone.

The other five conditions, 4-bit against BF16 (mean shift from *none*, kcal):

| **Model, set** | **us, 4-bit / BF16** | **jp_home, 4-bit / BF16** | **us_home, 4-bit / BF16** | **neutral dietitian, 4-bit / BF16** | **smaller, 4-bit / BF16** | **larger, 4-bit / BF16** |
| --- | --- | --- | --- | --- | --- | --- |
| 2B, NI | −19.4 / −1.5 | −25.1 / −1.3 | −14.5 / +2.4 | +0.4 / +3.5 | −30.5 / −14.6 | −26.3 / −10.9 |
| 2B, SN | −14.7 / −3.4 | −20.8 / −6.4 | −9.7 / −2.2 | +4.0 / +3.1 | −34.2 / −18.6 | −32.6 / −15.3 |
| 4B, NI | −1.0 / +10.2 | −21.0 / −18.7 | −9.6 / −0.6 | −4.9 / −0.2 | −8.6 / −7.1 | +9.6 / +14.7 |
| 4B, SN | −6.2 / +10.6 | −36.7 / −22.7 | −23.5 / −7.9 | −10.4 / +2.7 | −14.6 / −6.5 | +1.1 / +12.1 |
| 8B, NI | +6.7 / +4.5 | −5.5 / −11.9 | +3.5 / +5.2 | −7.0 / −2.9 | +9.3 / −0.0 | +39.0 / +44.2 |
| 8B, SN | +17.7 / +13.2 | −7.9 / −8.7 | −2.4 / +3.7 | +0.6 / +6.1 | +14.2 / +7.7 | +34.5 / +39.7 |

### **S4. Model-specific behaviours**

*Range responses in Gemma 3 4B.* Gemma 3 4B returned energy as a range (for example, 'approximately 350–450 kcal') in 86% of NutriImage and 62% of SNAPMe responses without a persona, in 40–50% under *jp_home* and *neutral_dietitian*, and in none under *jp* and *us*; Gemma 3 12B and 27B and all other models gave no range responses. Ranges were taken at their midpoint. Because the persona changed the form of the response as well as its value, Gemma 3 4B was excluded from the regressions; at the midpoint its baseline bias and JP shift on NutriImage were +177 and −116 kcal (master table), and with the lower bound of the range +120 and −60 kcal (859 images; ni887_Gemma3-4B-BF16_L0.jsonl, whose energy_kcal field records the lower bound).

*Reversed instruction following in GPT-5.6 Luna.* Larger − smaller was −15 kcal (NutriImage) and −36 kcal (SNAPMe); the mean estimates on SNAPMe were 477 kcal under *smaller*, 449 kcal without a persona, and 441 kcal under *larger* (NutriImage: 376, 373, and 360 kcal). Its baseline bias was the largest of all models (+149 and +130 kcal), and its JP shift was accordingly the largest (−72 and −44 kcal).

*Thinking tokens in the Gemini models.* With an output limit of 256 tokens, 8 of 24 (Gemini 3.8 Flash) and 6 of 24 (Gemini 3.1 Pro) test responses were truncated before the JSON was complete, because thinking tokens counted against the limit and could not be fully disabled; a truncated response could leave a partial number (for example, 'energy_kcal: 1'). Responses without a closing brace were therefore treated as failures for all models, and the limit was raised to 2,048 tokens for the two Gemini models.

*Responses dominated by ties in the small open models.* In Qwen3-VL 2B and 4B and in InternVL3.5, 60–80% of images received the same estimate under *jp* as without a persona (Section S6).

### **S5. JP persona and smaller instruction within cells**

For each of the 48 analysed cells: the JP shift and the smaller shift (kcal), their ratio (given where the smaller shift was at least 10 kcal in size), the rank correlation across images between JP − none and smaller − none, and MedAPE under the three conditions (images with all three estimates; unified reference values).

| **Group** | **Family** | **Model** | **Set** | **n** | **JP shift** | **Smaller shift** | **Ratio** | **Rank correlation within images** | **MedAPE (%): none / jp / smaller** |
| --- | --- | --- | --- | --- | --- | --- | --- | --- | --- |
| A | Gemma 3 | 12B | NI | 886 | −34.5 | −44.2 | 0.78 | 0.45 | 42.5 / 35.8 / 39.6 |
| A | Gemma 3 | 12B | SN | 1131 | −6.4 | −40.8 | 0.16 | 0.44 | 34.0 / 33.5 / 35.9 |
| A | Gemma 3 | 27B | NI | 886 | −49.7 | −65.7 | 0.76 | 0.10 | 57.4 / 40.2 / 42.3 |
| A | Gemma 3 | 27B | SN | 1131 | −25.0 | −57.3 | 0.44 | 0.39 | 32.5 / 31.7 / 32.3 |
| A | commercial−2025 | GPT−4o | NI | 873 | −2.2 | −8.8 | — | 0.43 | 37.1 / 32.2 / 36.9 |
| A | commercial−2025 | GPT−4o | SN | 1131 | −2.0 | −12.7 | 0.15 | 0.51 | 34.3 / 34.1 / 35.8 |
| A | commercial−2025 | GPT−4o−mini | NI | 879 | −5.3 | −21.2 | 0.25 | 0.45 | 38.6 / 35.4 / 37.1 |
| A | commercial−2025 | GPT−4o−mini | SN | 1131 | −8.2 | −22.2 | 0.37 | 0.46 | 34.1 / 34.3 / 35.0 |
| A | commercial−2025 | Gemini Flash (2025) | NI | 879 | −61.4 | −61.7 | 0.99 | 0.44 | 59.3 / 37.8 / 40.9 |
| A | commercial−2025 | Gemini Flash (2025) | SN | 1131 | −35.8 | −70.2 | 0.51 | 0.40 | 39.7 / 34.2 / 32.5 |
| A | commercial−2025 | Gemini Pro (2025) | NI | 879 | −39.8 | −120.7 | 0.33 | 0.36 | 40.6 / 29.6 / 34.8 |
| A | commercial−2025 | Gemini Pro (2025) | SN | 1131 | −34.9 | −150.4 | 0.23 | 0.37 | 28.4 / 30.3 / 43.0 |
| A | commercial−2025 | Haiku (2025) | NI | 878 | −26.3 | −43.0 | 0.61 | 0.40 | 50.9 / 44.0 / 43.6 |
| A | commercial−2025 | Haiku (2025) | SN | 1131 | −34.6 | −46.2 | 0.75 | 0.48 | 36.2 / 39.0 / 38.3 |
| A | commercial−2025 | Sonnet (2025) | NI | 879 | −21.6 | −42.0 | 0.51 | 0.22 | 36.2 / 33.3 / 35.6 |
| A | commercial−2025 | Sonnet (2025) | SN | 1131 | −16.3 | −6.2 | — | 0.15 | 31.9 / 32.8 / 33.4 |
| A | commercial−2026 | GPT−5.6 Luna | NI | 886 | −72.2 | +2.2 | — | 0.43 | 97.2 / 51.4 / 94.0 |
| A | commercial−2026 | GPT−5.6 Luna | SN | 1131 | −44.4 | +28.2 | −1.57 | 0.38 | 50.3 / 42.9 / 59.5 |
| A | commercial−2026 | GPT−5.6 Sol | NI | 886 | −39.4 | −20.7 | 1.90 | 0.30 | 51.3 / 35.1 / 42.8 |
| A | commercial−2026 | GPT−5.6 Sol | SN | 1131 | −22.3 | −20.4 | 1.09 | 0.43 | 33.2 / 31.9 / 31.5 |
| A | commercial−2026 | Gemini 3.1 Pro | NI | 886 | −11.4 | −92.1 | 0.12 | 0.26 | 34.7 / 29.4 / 40.9 |
| A | commercial−2026 | Gemini 3.1 Pro | SN | 1131 | −1.6 | −110.1 | 0.01 | 0.29 | 30.7 / 31.0 / 50.5 |
| A | commercial−2026 | Gemini 3.8 Flash | NI | 886 | −2.8 | −60.3 | 0.05 | 0.23 | 28.5 / 26.6 / 34.0 |
| A | commercial−2026 | Gemini 3.8 Flash | SN | 1131 | −17.1 | −80.5 | 0.21 | 0.43 | 28.9 / 31.2 / 38.7 |
| A | commercial−2026 | Haiku 4.5 | NI | 885 | −28.2 | −43.2 | 0.65 | 0.43 | 50.4 / 45.5 / 46.5 |
| A | commercial−2026 | Haiku 4.5 | SN | 1130 | −37.6 | −46.4 | 0.81 | 0.46 | 35.6 / 38.9 / 38.0 |
| A | commercial−2026 | Sonnet 5 | NI | 886 | −26.3 | −80.3 | 0.33 | 0.25 | 40.8 / 32.1 / 37.0 |
| A | commercial−2026 | Sonnet 5 | SN | 1131 | −22.5 | −94.3 | 0.24 | 0.31 | 30.1 / 30.9 / 40.8 |
| B | InternVL3.5 | 14B | NI | 886 | −2.7 | −10.5 | 0.26 | 0.50 | 47.2 / 47.2 / 44.9 |
| B | InternVL3.5 | 14B | SN | 1131 | +2.3 | −14.3 | −0.16 | 0.46 | 36.6 / 36.0 / 36.0 |
| B | InternVL3.5 | 2B | NI | 886 | +8.1 | −6.4 | — | 0.31 | 42.9 / 44.0 / 42.9 |
| B | InternVL3.5 | 2B | SN | 1131 | +6.5 | −8.2 | — | 0.44 | 41.3 / 40.2 / 43.1 |
| B | InternVL3.5 | 4B | NI | 886 | −0.2 | −4.2 | — | 0.51 | 41.3 / 41.9 / 42.3 |
| B | InternVL3.5 | 4B | SN | 1131 | +1.3 | −4.8 | — | 0.48 | 44.0 / 43.0 / 44.6 |
| B | InternVL3.5 | 8B | NI | 886 | −7.5 | +4.8 | — | 0.46 | 49.9 / 48.2 / 50.4 |
| B | InternVL3.5 | 8B | SN | 1131 | −7.3 | +1.4 | — | 0.53 | 37.0 / 36.0 / 35.1 |
| B | Mistral | Mistral Small 3.2 | NI | 886 | +9.2 | −3.7 | — | 0.34 | 39.8 / 40.1 / 40.8 |
| B | Mistral | Mistral Small 3.2 | SN | 1131 | +7.6 | −8.5 | — | 0.30 | 40.1 / 38.5 / 41.2 |
| B | Mistral | Pixtral−12B | NI | 886 | −0.8 | −29.7 | 0.03 | 0.38 | 50.4 / 48.8 / 44.1 |
| B | Mistral | Pixtral−12B | SN | 1131 | −8.7 | −38.4 | 0.23 | 0.32 | 32.8 / 32.9 / 34.1 |
| B | Qwen3−VL | 2B | NI | 886 | +1.3 | −14.6 | −0.09 | 0.44 | 44.1 / 42.6 / 42.8 |
| B | Qwen3−VL | 2B | SN | 1131 | −1.0 | −18.6 | 0.05 | 0.50 | 39.6 / 37.4 / 39.4 |
| B | Qwen3−VL | 32B | NI | 886 | −25.9 | −44.6 | 0.58 | 0.37 | 48.6 / 39.2 / 40.5 |
| B | Qwen3−VL | 32B | SN | 1131 | −14.2 | −48.8 | 0.29 | 0.45 | 32.5 / 32.4 / 33.4 |
| B | Qwen3−VL | 4B | NI | 886 | −2.9 | −7.1 | — | 0.61 | 38.1 / 37.8 / 38.1 |
| B | Qwen3−VL | 4B | SN | 1131 | −2.0 | −6.5 | — | 0.63 | 36.0 / 36.2 / 35.9 |
| B | Qwen3−VL | 8B | NI | 886 | −10.8 | −0.0 | — | 0.58 | 46.6 / 42.6 / 44.9 |
| B | Qwen3−VL | 8B | SN | 1131 | +8.4 | +7.7 | — | 0.71 | 35.0 / 34.1 / 33.6 |

Ratio: from near zero (Gemini 3.1 Pro, SNAPMe) to 1.9 (GPT-5.6 Sol, NutriImage); negative where the two shifts had opposite signs (GPT-5.6 Luna on SNAPMe, in which the smaller instruction raised the estimates). Rank correlation within images: 0.10–0.51 in group A and 0.30–0.71 in group B (median over cells 0.43); in group B most estimates are multiples of 50 kcal and the paired differences have many ties. MedAPE rose by more than 3 points under the smaller instruction in 7 cells and under the JP persona in 1. Script: s1_s5.py; values: s5_jp_vs_smaller.csv.

### **S6. Output grid and ties**

Most open models returned energy in multiples of 50 kcal: the proportion of estimates without a persona on that grid was 0.84–0.99 in the dense open models (lowest InternVL3.5 30B-A3B 0.77–0.84 and Qwen3-VL 32B 0.68–0.73; Qwen3-VL 30B-A3B 0.26–0.33), and 0.06–0.60 in the commercial models of 2026 (Gemini 3.1 Pro 0.51–0.60, Claude Sonnet 5 0.21–0.22, the others 0.06–0.31) (Table S6a; grid50_none_by_cell.csv). Table S6 gives, by family, the range across cells of the proportion of images whose estimate under *jp* was identical to that without a persona.

| **Family** | **Model** | **NI n** | **NI** | **SN n** | **SN** |
| --- | --- | --- | --- | --- | --- |
| Gemma 3 | 12B | 887 | 0.95 | 1134 | 0.86 |
| Gemma 3 | 27B | 887 | 0.93 | 1134 | 0.84 |
| Gemma 3 | 4B | 887 | 0.94 | 1134 | 0.88 |
| InternVL3.5 | 14B | 887 | 0.94 | 1134 | 0.90 |
| InternVL3.5 | 2B | 887 | 0.98 | 1134 | 0.93 |
| InternVL3.5 | 30B-A3B (MoE) | 887 | 0.84 | 1134 | 0.77 |
| InternVL3.5 | 4B | 887 | 0.90 | 1134 | 0.89 |
| InternVL3.5 | 8B | 887 | 0.98 | 1134 | 0.93 |
| Mistral | Mistral Small 3.2 24B | 887 | 0.97 | 1134 | 0.89 |
| Mistral | Pixtral-12B | 887 | 0.98 | 1134 | 0.93 |
| Qwen3-VL | 30B-A3B (MoE) | 887 | 0.26 | 1134 | 0.33 |
| Qwen3-VL | 32B | 887 | 0.68 | 1134 | 0.73 |
| Qwen3-VL | 2B | 887 | 0.99 | 1134 | 0.99 |
| Qwen3-VL | 4B | 887 | 0.96 | 1134 | 0.96 |
| Qwen3-VL | 8B | 887 | 0.99 | 1134 | 0.96 |
| Commercial 2026 | GPT-5.6 Luna | 887 | 0.16 | 1134 | 0.30 |
| Commercial 2026 | GPT-5.6 Sol | 887 | 0.06 | 1134 | 0.09 |
| Commercial 2026 | Gemini 3.1 Pro | 887 | 0.59 | 1134 | 0.51 |
| Commercial 2026 | Gemini 3.8 Flash | 887 | 0.06 | 1134 | 0.06 |
| Commercial 2026 | Claude Haiku 4.5 | 886 | 0.07 | 1134 | 0.12 |
| Commercial 2026 | Claude Sonnet 5 | 887 | 0.22 | 1134 | 0.21 |
| Qwen3-VL (MLX 4-bit) | 2B | 887 | 0.79 | 1134 | 0.90 |
| Qwen3-VL (MLX 4-bit) | 4B | 887 | 0.78 | 1134 | 0.82 |
| Qwen3-VL (MLX 4-bit) | 8B | 887 | 0.98 | 1134 | 0.96 |

Table S6a. Proportion of estimates without a persona that are multiples of 50 kcal, by cell (n: responses with a numeric estimate). Open models: BF16 on H100; the last three rows: MLX 4-bit on Apple M4 (Section S3).

| **Family (cells)** | **Tie under jp, range** | **Median** |
| --- | --- | --- |
| Qwen3-VL (10) | 0.38–0.81 | 0.61 |
| Gemma 3 (6) | 0.14–0.59 | 0.35 |
| InternVL3.5 (10) | 0.69–0.80 | 0.73 |
| Mistral (4) | 0.63–0.73 | 0.72 |
| Commercial 2026 (12) | 0.11–0.43 | 0.29 |
| Commercial 2025 (12) | 0.04–0.44 | 0.32 |

### **S7. Mixture-of-experts models**

| **Model** | **Set** | **n** | **Baseline bias** | **JP shift** | **down / up / tie** | **JP − US** | **Larger − smaller** | **MedAPE none → JP (%)** |
| --- | --- | --- | --- | --- | --- | --- | --- | --- |
| Qwen3-VL 30B-A3B | NI | 886 | +17 | +12 | 0.22 / 0.31 / 0.47 | +12 | +16 | 36.5 → 39.0 |
| Qwen3-VL 30B-A3B | SN | 1131 | −32 | −6 | 0.32 / 0.15 / 0.53 | −2 | +20 | 31.3 → 32.4 |
| InternVL3.5 30B-A3B | NI | 886 | −24 | −17 | 0.28 / 0.03 / 0.69 | −12 | +11 | 39.4 → 41.6 |
| InternVL3.5 30B-A3B | SN | 1131 | −97 | −9 | 0.19 / 0.04 / 0.77 | −8 | +15 | 37.5 → 38.5 |

Both models have about 3B active parameters. Their instruction following was at the level of the 4B dense models of the same family, and neither lay on the line of its family in Figure 2. Reported without interpretation.

### **S8. Likelihood scoring of Gemma 3 pt and it, and of InternVL3.5 8B pre-trained and final: details**

Table S8a. Median and mode of the scored distribution (NutriImage, 859 images; kcal).

| **Model** | **Statistic** | **Bias (none)** | **MedAPE (%)** | **ρ** | **JP − US** | **JP<US / JP>US** | **Larger − smaller** |
| --- | --- | --- | --- | --- | --- | --- | --- |
| 4B pt | median | −5 | 40.5 | 0.71 | −64 | 0.82 / 0.01 | +59 |
| 4B pt | mode | −81 | 44.8 | 0.68 | −51 | 0.49 / 0.04 | +42 |
| 4B it | median | +56 | 46.2 | 0.68 | −22 | 0.40 / 0.12 | +202 |
| 4B it | mode | +53 | 44.4 | 0.69 | −21 | 0.34 / 0.10 | +202 |
| 12B pt | median | −13 | 41.8 | 0.63 | −44 | 0.66 / 0.05 | +40 |
| 12B pt | mode | −66 | 44.9 | 0.63 | −30 | 0.37 / 0.07 | +35 |
| 12B it | median | +85 | 55.1 | 0.71 | −21 | 0.38 / 0.15 | +161 |
| 12B it | mode | +80 | 51.1 | 0.70 | −20 | 0.29 / 0.12 | +154 |
| 27B pt | median | +42 | 44.0 | 0.72 | −107 | 0.91 / 0.02 | +53 |
| 27B pt | mode | −37 | 40.9 | 0.67 | −81 | 0.61 / 0.04 | +42 |
| 27B it | median | +74 | 51.0 | 0.71 | −35 | 0.52 / 0.14 | +221 |
| 27B it | mode | +73 | 49.3 | 0.70 | −33 | 0.46 / 0.16 | +219 |

Table S8b. Contrast or shift by stratum of baseline error, with the stratifying condition not entering the difference (kcal; n in parentheses).

| **Model / input** | **Set** | **Stratified by** | **< 0** | **0–50** | **50–150** | **> 150** | **Slope** | **r** |
| --- | --- | --- | --- | --- | --- | --- | --- | --- |
| Gemma 3 4B pt | NI | none | −59 (376) | −58 (169) | −67 (242) | −97 (72) | -0.05 | -0.15 |
| Gemma 3 4B it | NI | none | −16 (267) | −29 (173) | −20 (237) | −25 (182) | -0.02 | -0.05 |
| Gemma 3 12B pt | NI | none | −45 (370) | −38 (157) | −44 (266) | −56 (66) | -0.01 | -0.01 |
| Gemma 3 12B it | NI | none | −6 (198) | −9 (105) | −23 (336) | −37 (220) | -0.09 | -0.21 |
| Gemma 3 27B pt | NI | none | −105 (255) | −103 (156) | −105 (306) | −117 (142) | -0.02 | -0.04 |
| Gemma 3 27B it | NI | none | −16 (232) | −33 (136) | −39 (284) | −54 (207) | -0.10 | -0.21 |
| Gemma 3 4B pt | SN | none | −52 (559) | −45 (170) | −49 (248) | −73 (98) | 0.00 | 0.01 |
| Gemma 3 4B it | SN | none | −16 (521) | −13 (202) | −15 (191) | −29 (161) | -0.02 | -0.04 |
| Gemma 3 12B pt | SN | none | −34 (362) | −26 (163) | −29 (323) | −47 (227) | -0.02 | -0.08 |
| Gemma 3 12B it | SN | none | −7 (404) | −2 (189) | −10 (276) | −20 (206) | -0.02 | -0.08 |
| Gemma 3 27B pt | SN | none | −68 (361) | −58 (176) | −63 (306) | −84 (232) | -0.02 | -0.07 |
| Gemma 3 27B it | SN | none | −13 (458) | −6 (216) | −18 (211) | −49 (190) | -0.06 | -0.18 |
| Sonnet4.6 image full (jp none) | NI | us | −4 (344) | −14 (213) | −30 (196) | −44 (100) | -0.11 | -0.26 |
| Sonnet4.6 name only (jp none) | NI | us | −2 (309) | −2 (183) | −22 (243) | −39 (118) | -0.11 | -0.27 |
| Sonnet4.6 cut25 (jp none) | NI | us | −26 (436) | −30 (117) | −70 (160) | −83 (140) | -0.11 | -0.21 |

Gemma 3 rows: JP − US under scoring. Claude Sonnet 4.6 rows: JP − none, on the 853 images common to the three inputs (Table 5).

Table S8a-SN. Median and mode of the scored distribution on SNAPMe (Gemma 3 27B; 1,075 images; kcal).

| **Model** | **Statistic** | **Bias (none)** | **MedAPE (%)** | **ρ** | **JP − US** | **JP<US / JP>US** | **Larger − smaller** |
| --- | --- | --- | --- | --- | --- | --- | --- |
| 27B pt | median | +38 | 35.6 | 0.76 | −68 | 0.80 / 0.01 | +77 |
| 27B pt | mode | −26 | 31.4 | 0.75 | −51 | 0.42 / 0.05 | +68 |
| 27B it | median | +11 | 31.0 | 0.73 | −19 | 0.34 / 0.13 | +220 |
| 27B it | mode | +10 | 31.7 | 0.72 | −16 | 0.23 / 0.10 | +215 |

Medians recomputed from the per-image candidate log-probabilities (cand_logp, 30 candidates) reproduce the values used in Table 7; mode = candidate with the highest probability. The mode on SNAPMe is given for 27B only. Values: gemma_ptit_snapme_mode_27B.csv.

Figure S3. Scored distributions for one image under three conditions.

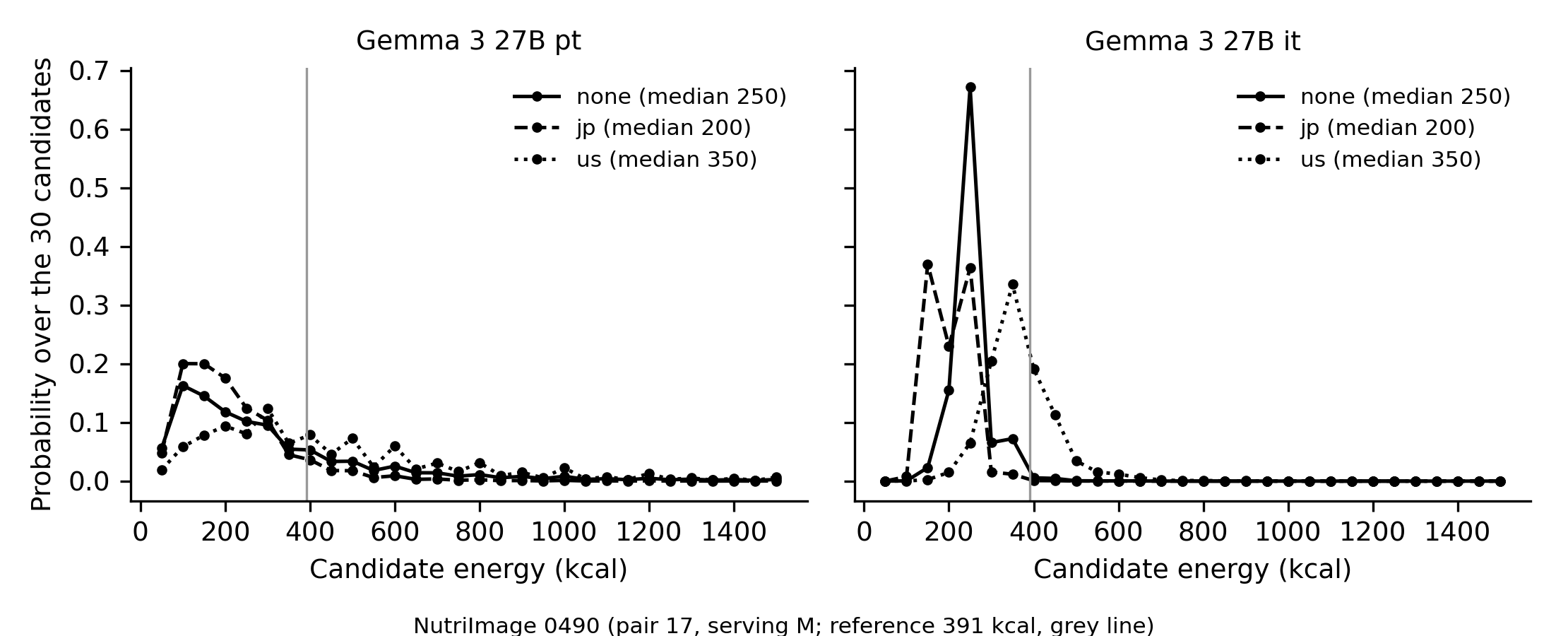

Probability over the 30 candidate values (50–1,500 kcal in steps of 50) for NutriImage 0490 (pair 17, serving M; reference 391 kcal, grey line) under none, jp, and us, for Gemma 3 27B pt (left) and it (right). Medians in the legend. The pt distribution is broad and moves as a whole; the it distribution is peaked and the persona moves the peak. Script: figS3.py (in the reproducibility package); source: ni887_Gemma3-27B-{pt,it}_probe.jsonl.

Table S8c. InternVL3.5 8B, pre-trained and final weights (NutriImage, 859 images; kcal).

| **Weights** | **Statistic** | **Bias (none)** | **MedAPE (%)** | **ρ** | **JP − US (95% CI)** | **JP<US / JP>US** | **Larger − smaller** |
| --- | --- | --- | --- | --- | --- | --- | --- |
| pre-trained | median, end-independent score | −27 | 41.9 | 0.67 | −57 (−60 to −54) | 0.76 / 0.01 | +44 |
| pre-trained | median, line-break score | −21 | 41.9 | 0.67 | −53 (−56 to −50) | 0.73 / 0.01 | +44 |
| pre-trained | expectation, end-independent score | +39 | 51.0 | 0.66 | −71 (−74 to −69) | 0.98 / 0.02 | +54 |
| final | median, end-independent score | +11 | 43.4 | 0.65 | −18 (−24 to −13) | 0.34 / 0.26 | +62 |
| final | median, line-break score | +65 | 54.3 | 0.60 | −29 (−35 to −23) | 0.38 / 0.18 | +65 |
| final | expectation, end-independent score | +44 | 49.7 | 0.66 | −23 (−28 to −18) | 0.58 / 0.42 | +70 |

End-independent score: log-likelihood of the number followed by the probability that the next token is not a digit; line-break score: as for Gemma 3 (Section 2.3(d)). Median of the probability placed on the 30 candidates: 0.03 (pre-trained) and 0.10 (final) with the end-independent score, 0.005 and 0.05 with the line-break score. JP − US by stratum of the error without a narrator (< 0, 0–50, 50–150, > 150 kcal; n 407, 189, 215, 48 in pt and 374, 156, 189, 140 in final): pre-trained −51, −51, −66, −92 (slope −0.05, r −0.17); final −2, −2, −8, −95 (slope −0.18, r −0.32). Bootstrap over images, 2,000 resamples. Script: iv_summary.py; values: internvl_ptit_ni_summary.csv.

Table S8d. Paired within-image differences between pre-trained and post-trained weights (kcal; post-trained minus pre-trained; bootstrap over images, 2,000 resamples).

| **Model** | **Set** | **n** | **JP − US, pt → post** | **Difference (95% CI)** | **Smaller in post** | **Larger − smaller, pt → post** | **Difference (95% CI)** | **Larger in post** |
| --- | --- | --- | --- | --- | --- | --- | --- | --- |
| Gemma 3 4B | NI | 859 | −64 → −22 | +42 (+38 to +47) | 0.59 | +59 → +202 | +143 (+138 to +148) | 0.94 |
| Gemma 3 4B | SN | 1075 | −52 → −17 | +35 (+29 to +41) | 0.57 | +69 → +205 | +136 (+131 to +141) | 0.90 |
| Gemma 3 12B | NI | 859 | −44 → −21 | +23 (+19 to +27) | 0.48 | +40 → +161 | +121 (+115 to +127) | 0.88 |
| Gemma 3 12B | SN | 1075 | −34 → −10 | +24 (+21 to +28) | 0.48 | +75 → +164 | +88 (+83 to +94) | 0.77 |
| Gemma 3 27B | NI | 859 | −107 → −35 | +71 (+66 to +77) | 0.75 | +53 → +221 | +168 (+162 to +176) | 0.93 |
| Gemma 3 27B | SN | 1075 | −68 → −19 | +49 (+45 to +53) | 0.68 | +77 → +220 | +143 (+135 to +151) | 0.85 |
| InternVL3.5 8B | NI | 859 | −57 → −18 | +38 (+32 to +44) | 0.63 | +44 → +62 | +18 (+14 to +21) | 0.39 |

Gemma 3: median of the line-break score; InternVL3.5 8B: median of the end-independent score. 'Smaller in post' and 'Larger in post': proportion of images in which the contrast is smaller, or larger − smaller is larger, after post-training. Script: paired_ptit.py; values: ptit_paired_diff.csv.

*Contrast of 27B regressed on the estimate and the reference value together (from the main text, Section 3.4).* Because images with large errors tend to be those with large estimates, the contrast of 27B was also regressed on the estimate without a narrator and on the reference value together. In it on NutriImage the two coefficients were of opposite sign and similar size (−0.090 and +0.103), as expected if the contrast depends on their difference, the error, and not on the size of the estimate alone; in pt neither coefficient differed from zero; in it on SNAPMe the coefficient of the estimate (−0.103) exceeded that of the reference (+0.037), so that on SNAPMe dependence on error was not separated from dependence on the size of the estimate. For Claude Sonnet 4.6 (JP − none regressed on the estimate under the US persona and on the reference value, 853 images) the coefficients were −0.106 (SE 0.016) and +0.117 (0.015) with the full image and −0.132 (0.015) and +0.105 (0.013) with the dish name only, of opposite sign and similar size as in Gemma 3 27B it on NutriImage; at the 25% crop they were −0.190 (0.024) and +0.074 (0.019), so that there dependence on error was not separated from dependence on the size of the estimate. Script: sonnet_recompute.py; values: table_input_ablation_sonnet_v2.csv.

### **S9. Same-dish pairs served at two sizes (NutriImage)**

| **Pair** | **Name given to the model (dish_name_en; larger / smaller where they differ)** | **Japanese name (romanised)** | **Sizes** | **Image, larger** | **Reference (kcal)** | **Image, smaller** | **Reference (kcal)** | **Ratio** | **Difference in the photographs** |
| --- | --- | --- | --- | --- | --- | --- | --- | --- | --- |
| 1 | Deep-fried tofu with starchy sauce | Ankake tofu | M / S | 0029 | 275.4 | 0030 | 138.0 | 2.00 | apparent |
| 2 | Tofu with nametake musuroom | Nametake tofu | M / S | 0453 | 131.8 | 0454 | 69.9 | 1.89 | apparent |
| 3 | Deep-fried tofu with kimchi | Kimchi agedashi tofu | M / S | 0312 | 309.8 | 0313 | 167.1 | 1.85 | apparent |
| 4 | Tofu with kimchi | Kimchi tofu | M / S | 0314 | 142.5 | 0315 | 72.2 | 1.97 | apparent |
| 5 | simmered tofu, chinese-style | Sutamina tofu | M / S | 0684 | 151.2 | 0685 | 75.8 | 2.00 | apparent |
| 6 | deep-fried chicken with chili sauce | Chikin chiri sosu | M / S | 0734 | 493.9 | 0735 | 315.6 | 1.56 | apparent |
| 7 | deep-fried chicken | Chikin karaage | M / S | 0720 | 516.2 | 0721 | 305.9 | 1.69 | not apparent |
| 8 | Minced meat cutlet | Minchi katsu | M / S | 0414 | 463.6 | 0415 | 311.6 | 1.49 | apparent |
| 9 | Boiled riceL / Boiled riceM | Raisu | L / M | 0564 | 545.4 | 0565 | 393.7 | 1.39 | not apparent |
| 10 | Boiled riceM / Boiled riceSS | Raisu | M / SS | 0565 | 393.7 | 0566 | 181.9 | 2.16 | not apparent |
| 11 | Deep-fried tofu with chinese sauce | Chuka agedashi tofu | M / S | 0810 | 357.2 | 0811 | 189.6 | 1.88 | apparent |
| 12 | Five-cereals boiled rice / Five-cereals boiled rice M | Gokoku gohan | M / S | 0138 | 461.2 | 0139 | 319.3 | 1.44 | apparent |
| 13 | Simmered pumpkin | Kabocha no sappari-ni | M / S | 0252 | 236.8 | 0253 | 145.7 | 1.62 | not apparent |
| 14 | Deep-fried tofu with amber sauce / Deep-fried-tofu with amber sauce | Agedashi tofu | M / S | 0004 | 267.5 | 0005 | 133.7 | 2.00 | apparent |
| 15 | Stir-fried chicken and asparagus / Stir-fride chicken and asparagus | Age-dori asupara itame | M / S | 0008 | 316.0 | 0009 | 216.3 | 1.46 | not apparent |
| 16 | Tofu with yuzu-miso | Yuzu-miso tofu | M / S | 0882 | 328.6 | 0883 | 187.2 | 1.76 | apparent |
| 17 | Deep-fried puffy tofu with minced chicken | Niku-miso tofu | M / S | 0490 | 390.7 | 0491 | 195.4 | 2.00 | apparent |
| 18 | Chicken and egg on rice | Oyakodon | M / S | 0540 | 691.7 | 0541 | 484.1 | 1.43 | not apparent |
| 19 | miso soup with pork and vegetables | Tonjiru | M / S | 0753 | 135.2 | 0754 | 83.4 | 1.62 | not apparent |
| 20 | Rice with salmon | Sake chirimen gohan | S / SS | 0600 | 276.5 | 0601 | 184.2 | 1.50 | reversed |

Classification by one author (S.N.) after the aggregate results had been seen, without sight of the outputs for individual pairs. Names are reproduced verbatim from the dataset, including misspellings, because they are the text given to the model in the name-only condition (full-width letters shown as ASCII). The two names of a pair were identical in 15 pairs and differed only in typography in pairs 2, 14, and 15; a size letter appears in the names of pairs 9 and 10 and in the name of the smaller serving of pair 12. Pairs 9 and 10 share image 0565.

Table S9b. Estimates of energy without a persona for the two servings of each pair (kcal; larger / smaller).

| **Pair** | **Reference** | **Sonnet 4.6 image** | **Sonnet 4.6 name only** | **Qwen3-VL 8B 4bit** | **GPT-5.6 Sol** | **GPT-5.6 Luna** | **Gemini 3.8 Flash** | **Gemini 3.1 Pro** | **Claude Sonnet 5** | **Claude Haiku 4.5** | **Gemma 3 27B it (probe median)** | **Gemma 3 27B pt (probe median)** |
| --- | --- | --- | --- | --- | --- | --- | --- | --- | --- | --- | --- | --- |
| 1 | 275 / 138 | 180 / 420 | 280 / 250 | 300 / 200 | 280 / 180 | 420 / 420 | 195 / 135 | 150 / 180 | 180 / 180 | 320 / 280 | 200 / 250 | 200 / 300 |
| 2 | 132 / 70 | 180 / 180 | 70 / 75 | 300 / 250 | 220 / 180 | 420 / 320 | 210 / 195 | 210 / 150 | 220 / 180 | 280 / 280 | 150 / 250 | 250 / 200 |
| 3 | 310 / 167 | 280 / 120 | 220 / 220 | 350 / 250 | 280 / 150 | 420 / 280 | 210 / 135 | 250 / 150 | 320 / 90 | 320 / 180 | 350 / 150 | 300 / 200 |
| 4 | 142 / 72 | 120 / 120 | 180 / 180 | 200 / 150 | 240 / 140 | 320 / 430 | 150 / 105 | 150 / 150 | 220 / 120 | 280 / 180 | 150 / 150 | 200 / 150 |
| 5 | 151 / 76 | 120 / 120 | 180 / 180 | 200 / 200 | 150 / 180 | 280 / 420 | 110 / 160 | 150 / 110 | 180 / 120 | 180 / 280 | 150 / 150 | 200 / 200 |
| 6 | 494 / 316 | 480 / 320 | 520 / 480 | 500 / 350 | 620 / 310 | 650 / 420 | 460 / 210 | 500 / 150 | 520 / 280 | 520 / 280 | 650 / 500 | 400 / 400 |
| 7 | 516 / 306 | 520 / 480 | 520 / 520 | 500 / 500 | 650 / 430 | 850 / 650 | 520 / 210 | 450 / 250 | 520 / 380 | 520 / 420 | 650 / 550 | 600 / 550 |
| 8 | 464 / 312 | 620 / 420 | 290 / 250 | 600 / 400 | 520 / 340 | 720 / 520 | 620 / 410 | 650 / 420 | 550 / 420 | 520 / 380 | 750 / 450 | 650 / 350 |
| 9 | 545 / 394 | 200 / 280 | 330 / 252 | 200 / 300 | 260 / 310 | 310 / 330 | 290 / 250 | 205 / 205 | 200 / 200 | 380 / 206 | 200 / 300 | 250 / 250 |
| 10 | 394 / 182 | 280 / 210 | 252 / 168 | 300 / 200 | 310 / 260 | 330 / 260 | 250 / 240 | 205 / 205 | 200 / 200 | 206 / 150 | 300 / 250 | 250 / 200 |
| 11 | 357 / 190 | 420 / 180 | 280 / 280 | 300 / 200 | 320 / 230 | 520 / 320 | 220 / 140 | 250 / 150 | 350 / 180 | 280 / 280 | 250 / 250 | 200 / 200 |
| 12 | 461 / 319 | 350 / 320 | 320 / 320 | 300 / 300 | 430 / 430 | 520 / 520 | 285 / 280 | 250 / 280 | 220 / 220 | 280 / 280 | 350 / 250 | 300 / 250 |
| 13 | 237 / 146 | 180 / 180 | 110 / 110 | 200 / 150 | 210 / 280 | 320 / 320 | 185 / 180 | 150 / 250 | 150 / 220 | 320 / 320 | 150 / 250 | 150 / 200 |
| 14 | 268 / 134 | 210 / 120 | 280 / 320 | 250 / 200 | 170 / 170 | 320 / 420 | 165 / 140 | 150 / 150 | 180 / 180 | 280 / 280 | 150 / 350 | 200 / 300 |
| 15 | 316 / 216 | 180 / 280 | 220 / 220 | 150 / 300 | 120 / 230 | 180 / 520 | 140 / 160 | 150 / 280 | 180 / 320 | 120 / 280 | 200 / 450 | 150 / 300 |
| 16 | 329 / 187 | 180 / 180 | 120 / 120 | 250 / 250 | 180 / 180 | 280 / 420 | 195 / 185 | 150 / 150 | 180 / 180 | 280 / 280 | 200 / 250 | 200 / 200 |
| 17 | 391 / 195 | 280 / 180 | 280 / 280 | 300 / 200 | 320 / 220 | 520 / 380 | 240 / 155 | 150 / 250 | 220 / 220 | 280 / 180 | 250 / 150 | 250 / 250 |
| 18 | 692 / 484 | 420 / 580 | 650 / 650 | 250 / 500 | 520 / 620 | 520 / 620 | 620 / 650 | 450 / 600 | 320 / 550 | 320 / 280 | 300 / 450 | 300 / 350 |
| 19 | 135 / 83 | 40 / 85 | 180 / 150 | 300 / 300 | 70 / 210 | 120 / 220 | 135 / 125 | 45 / 150 | 60 / 120 | 150 / 380 | 100 / 250 | 150 / 200 |
| 20 | 276 / 184 | 215 / 180 | 520 / 520 | 250 / 250 | 360 / 290 | 520 / 330 | 260 / 240 | 200 / 210 | 220 / 220 | 280 / 280 | 200 / 250 | 250 / 200 |

Claude Sonnet 4.6: image (full) and name-only conditions (Section 2.3(b)); Qwen3-VL 8B: MLX 4-bit chat run (Section S3); the six commercial models of 2026: run 1 of the main experiment. Gemma 3 27B: median of the scored distribution (Section S8). Values: s9_pairs_per_model.csv.

### **S10. Crop and name-only experiments: additional results**

*Looser criterion of identification.* When agreement of the main ingredient alone was required, 170 of 855 images (19.9%) were judged SAME at the 25% crop, and the JP shift at the crop was −22 kcal in these and −50 kcal in the 685 judged DIFF (strict criterion: 134 and 721 images, −20 and −49 kcal; cut_judge_loose_none.jsonl).

*Names returned without a persona and under a persona, and the shift by agreement of the names (Claude Sonnet 4.6; strict criterion; under jp one name is in Japanese).*

| **Images, persona** | **Names differ** | **Shift, all (kcal)** | **Names agree: shift (n)** | **Names differ: shift (n)** |
| --- | --- | --- | --- | --- |
| Full image, jp | 0.42 | −17 | −9 (494) | −28 (364) |
| Full image, us | 0.20 | −1 | −3 (683) | +3 (175) |
| 25% crop, jp | 0.73 | −44 | −16 (228) | −55 (628) |
| 25% crop, us | 0.35 | −2 | 0 (559) | −5 (299) |

| **JP shift by stability of the name** | **Full image** | **25% crop** |
| --- | --- | --- |
| Name stable under us, renamed under jp | −25 (257) | −52 (367) |
| Name stable under us, not renamed under jp | −9 (426) | −14 (192) |
| P(renamed under jp \| name differs under us) vs P(renamed under jp \| name stable under us) | 0.61 vs 0.38 | 0.88 vs 0.66 |

| **25% crop, by retention of identification (none, crop vs full)** | **Names agree (none vs jp)** | **Names differ** |
| --- | --- | --- |
| Identification retained (134) | −9 (74) | −34 (60) |
| Identification not retained (721) | −19 (153) | −57 (568) |

*Human rating of 100 full-image pairs (one author, against the original menu name).*

|  | **Name under jp correct** | **Name under jp not correct** |
| --- | --- | --- |
| Name without persona correct | 59 | 3 |
| Name without persona not correct | 6 | 32 |

Correct: 62 without a persona and 65 under jp (exact McNemar *P*=.51). The two names were taken to denote different dishes where only one was correct (9 pairs) and the same dish where both were correct (59); where neither was correct (32), the rater judged whether they denoted the same dish (different in 13). Different dishes in all: 22 of 100 (95% CI 15–31): 3 from a correct to an incorrect name, 6 from an incorrect to a correct name, 13 between two incorrect names. Model judge on the same 100 pairs: different in 46; agreement with the human rating 68%, κ = 0.33; of the 46, 18 were different dishes by human rating.

*JP shift in the rated pairs by human rating and model judge (96 pairs with reference energy above 20 kcal).*

| **Group** | **n** | **Mean (kcal)** | **95% CI** | **Median** | **Lower / higher / equal** | **US shift** |
| --- | --- | --- | --- | --- | --- | --- |
| Same dish (human); names judged the same | 47 | +3 | −9 to 15 | 0 | 0.30 / 0.32 / 0.38 | −6 |
| Same dish (human); names judged different (wording) | 27 | −28 | −45 to −13 | −10 | 0.59 / 0.07 / 0.33 | −1 |
| Different dishes (human) | 22 | −37 | −70 to −7 | −5 | 0.50 / 0.14 / 0.36 | −9 |

Mean error without a persona: −5, +45, and −41 kcal in the three groups. Without the pairs with an absolute difference of 100 kcal or more, the means were −1, −18, and −5 kcal. Difference between the second and the first group: −31 kcal (95% CI −52 to −11).

Expectations written before the none–us control: disagreement 20–30% at full size (observed 20%); US shift within ±5 kcal in both strata (observed −3 and +3 kcal). Human rating of the none–jp pairs (100 pairs at the 25% crop, drawn at random; the same criterion as the judge, rated by an author with the model judgement hidden; nonevsjp_validation_100_SN.xlsx): 78 pairs were rated the same dish and 22 different, against 54 and 46 by the judge (agreement 68%; in 28 pairs the judge said different where the rater said the same, mostly a Japanese rendering of the same dish, and in 4 the reverse). Against the menu name, the name was correct in 62 pairs without the persona and in 65 under it (both correct in 59). The JP shift at the crop was +3 kcal in the 47 pairs rated the same by both, −28 kcal in the 27 rated the same by the rater but different by the judge, and −37 kcal in the 22 rated different (96 pairs with estimates; nonevsjp_100_shift_by_group.csv).

*JP − none by stratum of the error under the US persona, 25% crop (from the main text, Section 3.2).* At the 25% crop the slope was the same as at full size (−0.11) but the shift was already −26 kcal in the lowest stratum (−26, −30, −70, −83 kcal; intercept −46 against −15 kcal), that is, the dependence on error was retained and a roughly constant lowering was added. At full size the values were −4, −14, −31, and −45 kcal across the four strata.

*Human rating of 100 pairs of names from the full image and the 25% crop (no persona; one author; the two names compared with each other, with the criterion given to the judge provided and the original menu name in view; the judge's output not seen).*

|  | **Judge: SAME** | **Judge: DIFF** |
| --- | --- | --- |
| Rater: SAME | 16 | 14 |
| Rater: DIFF | 2 | 68 |

Agreement 84 of 100; κ = 0.57. Names the same: rater 30, judge 18. Of the 16 disagreements, 14 were pairs that the rater found the same and the judge different. One pair (no. 75), first left undecided by the rater, was settled as DIFF. Source: judge_validation_100_final.csv.

*One-serving statement.* With the statement that the image shows one serving, identification retention was 59.4%, 37.3%, and 15.8% at the 60%, 40%, and 25% crops (without: 60.0%, 39.2%, 15.6%), and MedAPE without a persona was 33.1%, 39.1%, 46.3%, and 56.1% from the full image to the 25% crop (without: 34.4%, 39.9%, 47.1%, 56.1%).

*Name-only condition restricted to stable identification.* In the 511 of the 853 images judged SAME at the 60% crop, the name-only and image-based estimates correlated at ρ = 0.71 (all 853 images: 0.68), and the JP shift was −17.6 kcal with the image and −12.6 kcal with the name only (all 853 images: −17.2 and −13.0).

### **S11. ACETADA: strata by remaining fraction by weight**

| **Measure** | **Sonnet 4.6 (chat)** | **Qwen3-VL 8B (chat)** | **Gemma 3 27B it (chat)** | **Gemma 3 27B it (scoring)** | **Gemma 3 27B pt (scoring)** |
| --- | --- | --- | --- | --- | --- |
| Ratio after ÷ before, remaining weight ≤ 5% (n = 337) | 0.42 | 0.42 | 0.75 | 0.67 | 0.86 |
| 5–20% (258) | 0.45 | 0.50 | 0.80 | 0.75 | 0.85 |
| 20–50% (188) | 0.56 | 0.63 | 0.86 | 0.82 | 0.86 |
| > 50% (20) | 0.77 | 0.88 | 1.00 | 0.93 | 0.93 |
| ρ, ratio of estimates vs remaining fraction by weight | 0.24 | 0.48 | 0.21 | 0.29 | 0.07 |

*Mode in place of median (scoring format).* Gemma 3 27B pt: proportion with after < before 0.69 (median 0.78), overall ratio 0.75 (0.86), ρ with the remaining fraction by weight 0.21 (0.07). Gemma 3 27B it: 0.83 (0.87), 0.73 (0.75), 0.31 (0.29).

*Expectations written before the runs (from the main text, Section 3.1).* The expectations written before these runs referred to the remaining fraction by weight. Those for the direction in Sonnet 4.6 and Qwen3-VL 8B (≥ 0.8) and for the ratio in Gemma 3 27B pt (0.85; observed 0.86) were met; the direction in Gemma 3 27B it was read more often than expected (0.74 against 0.5–0.6); and the correlations were lower than expected for Sonnet 4.6 (0.24 against 0.5) and slightly so for Qwen3-VL 8B (0.48 against 0.5).

*What the models wrote about after-meal photographs (pairs with 5% or less of the served energy remaining; n = 344).* Claude Sonnet 4.6 (342 pairs with both estimates): the dish name of the after-meal photograph contained finished, remnants, empty, eaten, leftover, consumed, or residue in 298 (87%); for these the after-meal estimate had a median of 280 kcal, was 0 kcal in 7%, and was 100 kcal or more in 78%. Qwen3-VL 8B (343 pairs): the basis stated that the tray was empty or that no food remained in 51 (15%), and the estimate was then 0 kcal in 42 (82%); over all 343 after-meal photographs the estimate was 0 kcal in 13%. Gemma 3 27B it (chat): such a statement occurred in 2 photographs. Classification was by word search in the dish name (Claude Sonnet 4.6) and in the basis (Qwen3-VL 8B, Gemma 3 27B it), without human reading of all texts. Scoring format: median entropy of the distribution over the 30 candidates, 1.6 (it) and 3.1 (pt; maximum 3.4); median of the distribution for before- and after-meal photographs of these pairs, 600 and 400 kcal (it) and 650 and 550 kcal (pt); the lowest candidate (50 kcal) was the median in 1% (it) and 0% (pt) of after-meal photographs. Script: acetada_basis.py.

The remaining fraction by weight includes beverages. The expectations written before the runs referred to this fraction (main text, Section 3.1). Excluded records: two whose image paths duplicated those of another meal of the same participant and day (participant 3088, 2021-10-19, and 3057, 2021-08-03; the Dinner records), and, for energy measures, 12 meals in which recorded consumption exceeded the served amount.

### **S12. Repeated runs of the same conditions, and apparent slopes**

Table S12. Runs of the same conditions on separate occasions (chat format, no persona).

| **Runs compared** | **Images** | **Within-image SD without a persona (kcal)** | **Proportion of images with the same value** | **Between-image SD (kcal)** |
| --- | --- | --- | --- | --- |
| Claude Haiku 4.5, NutriImage (July/August 2026 and September 2026) | 853 | 33 | 0.67 | 145 |
| Claude Haiku 4.5, SNAPMe (June/August 2026 and September 2026) | 1,063 | 45 | 0.63 | 191 |
| Gemini 2.5 Flash, NutriImage (July and August 2026) | 714 | 70 | 0.12 | 238 |

*Reproduction of cell means and of the change of individual images.* Mean paired differences in the two runs: Claude Haiku 4.5, NutriImage, JP −26.9 and −28.1, smaller −44.0 and −44.3, larger +119.0 and +120.7 kcal; SNAPMe, JP −35.9 and −39.5 kcal; Gemini 2.5 Flash, NutriImage, JP −57.1 and −64.6 kcal. Correlation between runs of the change of the individual image (kcal): Claude Haiku 4.5, NutriImage, JP 0.46, smaller 0.49, larger 0.57, us 0.33; Gemini 2.5 Flash, NutriImage, JP 0.34, us 0.12–0.19; Claude Haiku 4.5, SNAPMe, JP 0.73, smaller 0.66, larger 0.48, us 0.67 by Pearson's r, and 0.42, 0.40, 0.53, and 0.31 by rank (mean shifts in the two runs: smaller −49.2 and −49.6, larger +116.0 and +113.5, us −4.0 and −4.5 kcal). The July run of Gemini 2.5 Flash contained null responses, which were not at random; only images valid in both runs were used.

*Apparent slopes in a difference from the estimate without a persona.* (a) Variation between requests. Under the US persona, whose true shift is close to zero, stratification by the error of *none* in the same run gave a gradient (log2; Claude Haiku 4.5, NutriImage, +0.10 to −0.08; Gemini 2.5 Flash, NutriImage, +0.07 and +0.04 in the lowest stratum to −0.05 and −0.11 in the highest, in the two runs), which became flat when the error of *none* was taken from the other run. About four tenths of the JP gradient was of this kind. (b) Deviations specific to the image in the *none* condition that recur between runs. For Claude Haiku 4.5 on SNAPMe, a gradient in the direction of that of JP remained under *us*, *us_home*, and *neutral_dietitian* even with the error of the other run (us: +0.04 to −0.18 and −0.24); that the change under *us*, with a mean of zero, was reproduced image by image (rank correlation 0.31 in kcal; r = 0.49 in log2) supports this. A contrast between two conditions other than *none*, stratified by the error of *none*, carries neither component, and is the form used in the main text (Section 3.5), as for Gemma 3 under scoring (Section 3.4).

*JP shift where the model underestimated (kcal).* With the error of *none* from the other run: Claude Haiku 4.5, −11 to −13; Gemini 2.5 Flash, −19 to −23. With the error of *none* from the same run: −6 to −9 and +7 to +11.

Runs compared (p42_repeats.py): Claude Haiku 4.5, the run of the previous study and the run of September 2026 of the present study, with the same model identifier and prompts (output limit 500 and 256 tokens); Gemini 2.5 Flash, the run of July 2026 of the previous study, which contained null responses, and a re-run of 15–16 August 2026. The run of the previous study for Claude Haiku 4.5 combines the none, jp, and us conditions of 4–5 July 2026 (NutriImage, cropped series) and 28 June 2026 (SNAPMe), as reported in that study [1], with the smaller, larger, jp_home, us_home, and neutral conditions of 15–19 August 2026, added during its revision. Images with reference energy above 20 kcal and a valid estimate in both runs. Checked against p42b_conditions.csv, p42b_shift_repro.csv, and p42b_form.csv.

### **S15. Model generation (from the main text, Section 3.6)**

Fitted separately, the 2025 and 2026 commercial cells gave nearly the same line (main text, Section 3.6), although baseline bias changed between generations within a vendor (e.g., Gemini Flash, +119 kcal in the 2025 model and −13 kcal in the 2026 model on NutriImage, with JP shifts of −60 and −3 kcal). Claude Haiku 4.5 was included in both groups and gave similar values on the two occasions (NutriImage: bias +46 and +44 kcal, JP shift −26 and −28 kcal; SNAPMe: −35 and −38 kcal, −34 and −38 kcal).

### **Table S13. MedAPE (%) under prompt conditions and after numerical calibration, all cells of 2026 (chat format): median over 200 random splits (2.5th–97.5th percentiles)**

| **Model** | **Set** | **n** | **Baseline bias (kcal)** | **none** | **jp** | **smaller** | **Mean bias subtracted** | **Divided by median ratio** | **jp − subtraction** | **jp − division** |
| --- | --- | --- | --- | --- | --- | --- | --- | --- | --- | --- |
| GPT-5.6 Luna | NI | 852 | +151 | 92.8 (81.1–101.0) | 49.9 (44.3–54.5) | 90.8 (81.6–100.4) | 46.7 (41.8–53.2) | 39.5 (37.3–42.5) | +2.8 (−4.6 to +8.8) | +10.4 (+3.6 to +14.9) |
| GPT-5.6 Sol | NI | 855 | +70 | 48.5 (44.9–53.4) | 34.4 (31.8–37.3) | 41.2 (37.7–45.4) | 41.7 (37.9–45.1) | 35.3 (32.8–37.9) | −7.5 (−10.7 to −4.1) | −1.1 (−3.7 to +2.1) |
| Claude Sonnet 5 | NI | 854 | +20 | 39.2 (36.0–42.6) | 31.3 (29.2–33.5) | 36.1 (33.8–38.0) | 36.8 (33.9–39.1) | 35.1 (32.9–37.9) | −5.3 (−7.4 to −2.7) | −3.8 (−6.0 to −1.6) |
| Gemini 3.1 Pro | NI | 854 | +3 | 34.3 (30.8–36.2) | 29.1 (27.7–30.7) | 40.8 (38.6–43.4) | 34.2 (31.6–36.4) | 32.6 (29.9–34.9) | −5.2 (−7.4 to −2.8) | −3.4 (−5.9 to −1.4) |
| Gemini 3.8 Flash | NI | 854 | −13 | 28.1 (26.5–30.5) | 26.6 (24.1–29.2) | 34.4 (32.2–36.1) | 29.0 (26.5–31.7) | 28.1 (25.8–30.3) | −2.3 (−5.5 to +0.3) | −1.4 (−3.3 to +0.7) |
| Claude Haiku 4.5 (2026 run) | NI | 852 | +44 | 49.1 (43.9–54.5) | 43.3 (39.0–46.9) | 44.4 (39.8–50.0) | 47.9 (44.5–52.5) | 43.5 (41.0–47.2) | −5.1 (−9.1 to −0.7) | −0.6 (−3.8 to +3.4) |
| Gemma 3 4B (range responses; reference only) | NI | 855 | +120 | 77.3 (67.2–85.3) | 45.2 (41.6–50.4) | 51.6 (47.3–56.0) | 52.4 (47.8–56.5) | 40.2 (37.0–43.6) | −6.9 (−12.5 to −1.2) | +5.2 (+0.6 to +11.1) |
| Gemma 3 12B | NI | 855 | +36 | 41.5 (38.4–43.8) | 34.7 (31.7–38.2) | 38.7 (36.8–41.5) | 39.9 (36.7–42.6) | 38.2 (36.0–40.6) | −5.5 (−8.6 to −1.1) | −3.8 (−6.7 to −0.5) |
| Gemma 3 27B | NI | 855 | +88 | 55.2 (50.8–63.1) | 38.9 (35.5–42.6) | 40.9 (37.5–44.2) | 46.2 (43.1–50.8) | 38.7 (36.6–41.8) | −7.3 (−11.1 to −3.5) | +0.2 (−3.5 to +4.0) |
| Qwen3-VL 2B | NI | 854 | −2 | 42.3 (39.0–45.7) | 41.7 (38.6–44.1) | 41.9 (38.7–44.1) | 42.7 (39.0–48.2) | 41.3 (38.5–43.9) | −1.3 (−5.1 to +0.8) | −0.1 (−2.9 to +3.2) |
| Qwen3-VL 4B | NI | 853 | −22 | 36.9 (34.9–39.5) | 36.6 (33.6–39.5) | 36.8 (34.3–39.2) | 37.1 (34.6–41.5) | 37.1 (34.4–39.7) | −0.8 (−3.9 to +1.8) | −0.6 (−2.9 to +1.6) |
| Qwen3-VL 8B | NI | 853 | +35 | 44.2 (41.4–48.7) | 41.4 (38.7–44.3) | 43.3 (39.5–47.5) | 38.6 (35.7–41.3) | 37.5 (35.2–39.5) | +2.6 (+0.3 to +5.1) | +3.9 (+0.8 to +6.1) |
| Qwen3-VL 32B | NI | 854 | +43 | 45.3 (41.7–51.2) | 37.1 (33.7–40.8) | 39.6 (36.4–42.2) | 42.3 (39.2–46.2) | 38.6 (35.5–41.5) | −5.1 (−7.9 to −1.9) | −1.3 (−4.4 to +2.0) |
| Qwen3-VL 30B-A3B | NI | 854 | +16 | 35.7 (33.0–39.0) | 38.1 (34.8–41.4) | 35.1 (32.3–39.0) | 36.8 (34.5–38.9) | 34.6 (32.5–36.8) | +1.4 (−1.4 to +4.1) | +3.4 (+1.0 to +6.0) |
| InternVL3.5 2B | NI | 850 | −44 | 41.3 (39.3–44.0) | 43.0 (40.6–45.3) | 42.0 (39.8–44.3) | 44.4 (40.5–48.3) | 41.9 (39.3–45.1) | −1.5 (−6.6 to +2.2) | +0.9 (−1.1 to +2.9) |
| InternVL3.5 4B | NI | 854 | −67 | 40.6 (37.6–42.6) | 40.9 (38.8–43.3) | 41.7 (39.0–43.9) | 45.1 (40.7–49.6) | 38.8 (35.7–41.6) | −4.2 (−8.7 to +0.9) | +2.3 (−1.6 to +5.3) |
| InternVL3.5 8B | NI | 854 | +34 | 48.1 (43.6–51.6) | 46.6 (41.8–51.3) | 48.4 (45.0–53.5) | 45.2 (42.3–48.2) | 43.1 (40.3–46.4) | +1.2 (−2.4 to +4.2) | +3.1 (−1.8 to +7.5) |
| InternVL3.5 14B | NI | 854 | +12 | 45.0 (41.9–49.2) | 45.6 (42.1–49.2) | 43.9 (40.5–47.3) | 43.1 (39.9–47.4) | 41.7 (38.6–44.9) | +2.3 (−0.4 to +4.7) | +4.1 (+0.8 to +6.7) |
| InternVL3.5 30B-A3B | NI | 854 | −26 | 38.2 (36.4–41.4) | 40.7 (38.3–42.7) | 39.5 (36.5–41.9) | 40.7 (36.8–44.7) | 38.5 (36.1–41.5) | 0.0 (−3.9 to +3.5) | +2.2 (−0.4 to +4.7) |
| Pixtral-12B | NI | 854 | +45 | 48.5 (43.2–55.2) | 47.0 (42.5–51.8) | 43.0 (39.4–46.8) | 44.7 (40.6–48.9) | 43.3 (40.4–46.2) | +1.9 (−1.7 to +5.4) | +3.5 (−1.1 to +8.5) |
| Mistral Small 3.2 24B | NI | 854 | −31 | 38.3 (35.8–41.5) | 39.3 (37.1–42.0) | 39.4 (36.6–42.6) | 39.8 (35.7–42.8) | 39.0 (35.9–41.8) | −0.4 (−3.0 to +2.8) | +0.2 (−1.7 to +2.2) |
| GPT-5.6 Luna | SN | 1074 | +134 | 49.4 (46.4–55.7) | 41.4 (38.2–46.0) | 58.4 (54.1–64.8) | 46.7 (43.5–50.9) | 32.5 (31.0–34.8) | −5.6 (−9.6 to −1.9) | +8.6 (+5.6 to +12.3) |
| GPT-5.6 Sol | SN | 1075 | +31 | 32.4 (29.2–35.0) | 30.9 (28.6–33.1) | 31.1 (28.5–33.2) | 33.2 (30.5–35.6) | 30.5 (27.9–32.8) | −2.5 (−4.9 to 0.0) | +0.2 (−1.7 to +2.0) |
| Claude Sonnet 5 | SN | 1074 | −41 | 28.7 (26.6–30.4) | 29.8 (27.7–31.4) | 40.5 (39.6–42.8) | 33.4 (30.9–37.1) | 28.6 (26.7–30.6) | −3.6 (−8.0 to −0.6) | +1.3 (−0.9 to +2.9) |
| Gemini 3.1 Pro | SN | 1074 | −56 | 29.8 (28.2–32.3) | 29.4 (28.2–31.9) | 50.2 (49.0–51.5) | 35.2 (32.3–37.5) | 30.1 (27.6–32.5) | −5.5 (−8.5 to −2.4) | −0.5 (−3.2 to +1.8) |
| Gemini 3.8 Flash | SN | 1074 | −50 | 27.9 (25.5–29.7) | 29.8 (28.1–32.0) | 38.4 (36.5–41.0) | 33.7 (30.7–36.5) | 28.1 (26.0–30.4) | −3.6 (−7.5 to −0.4) | +1.9 (−1.3 to +3.8) |
| Claude Haiku 4.5 (2026 run) | SN | 1055 | −41 | 34.4 (32.1–36.2) | 37.0 (34.7–39.3) | 36.8 (34.7–38.6) | 37.3 (34.1–40.6) | 34.0 (31.4–36.9) | −0.5 (−4.1 to +3.7) | +3.0 (+0.7 to +5.5) |
| Gemma 3 4B (range responses; reference only) | SN | 1075 | −21 | 34.7 (32.9–37.5) | 33.0 (31.0–35.6) | 34.0 (32.0–35.7) | 36.8 (33.5–39.6) | 34.9 (33.1–37.4) | −3.8 (−6.7 to −0.6) | −2.0 (−4.4 to 0.0) |
| Gemma 3 12B | SN | 1074 | −50 | 31.9 (29.9–34.4) | 31.1 (29.8–34.0) | 34.6 (32.7–36.7) | 38.3 (35.8–41.9) | 32.6 (30.6–35.3) | −7.0 (−10.6 to −3.6) | −1.2 (−3.7 to +0.7) |
| Gemma 3 27B | SN | 1074 | −8 | 30.4 (28.9–33.0) | 30.8 (28.8–32.6) | 30.8 (29.5–33.0) | 32.0 (29.6–36.0) | 30.5 (28.2–33.0) | −1.4 (−4.3 to +1.0) | +0.1 (−1.7 to +2.1) |
| Qwen3-VL 2B | SN | 1074 | −75 | 37.5 (36.0–40.2) | 35.7 (33.4–37.7) | 37.5 (35.7–39.5) | 41.5 (39.0–45.3) | 37.4 (35.0–40.3) | −5.8 (−9.4 to −2.8) | −1.7 (−4.3 to +0.1) |
| Qwen3-VL 4B | SN | 1057 | −84 | 34.0 (32.6–35.9) | 33.9 (31.6–36.0) | 33.2 (31.7–35.9) | 36.8 (34.0–40.6) | 32.9 (30.5–34.9) | −2.8 (−7.1 to +0.8) | +1.1 (−1.1 to +3.4) |
| Qwen3-VL 8B | SN | 1072 | −22 | 32.5 (30.1–35.0) | 31.7 (29.5–34.4) | 31.1 (29.0–33.8) | 35.3 (32.5–37.4) | 32.6 (30.3–34.8) | −3.5 (−5.8 to −1.2) | −0.9 (−2.7 to +0.7) |
| Qwen3-VL 32B | SN | 1070 | −22 | 30.9 (28.0–33.9) | 31.0 (28.6–32.8) | 32.0 (30.1–34.5) | 34.1 (31.0–37.0) | 31.2 (28.5–34.2) | −3.2 (−5.8 to 0.0) | −0.2 (−2.0 to +1.7) |
| Qwen3-VL 30B-A3B | SN | 1072 | −36 | 30.7 (28.5–32.9) | 31.1 (28.8–33.4) | 30.6 (28.6–32.8) | 33.4 (31.0–36.3) | 30.8 (28.5–32.8) | −2.3 (−5.0 to +1.2) | +0.5 (−1.4 to +2.0) |
| InternVL3.5 2B | SN | 1068 | −123 | 39.1 (37.0–41.3) | 38.3 (36.0–40.1) | 41.1 (38.7–43.3) | 41.4 (38.2–45.5) | 35.6 (33.6–37.5) | −3.3 (−8.3 to +1.0) | +2.6 (+0.4 to +4.3) |
| InternVL3.5 4B | SN | 1071 | −130 | 42.4 (40.1–44.5) | 41.0 (38.7–43.2) | 42.8 (40.5–45.1) | 40.7 (37.5–44.0) | 36.8 (34.2–40.0) | +0.4 (−3.6 to +5.2) | +4.3 (+0.8 to +8.0) |
| InternVL3.5 8B | SN | 1072 | −70 | 34.5 (32.1–37.1) | 34.6 (32.3–36.0) | 32.9 (30.6–35.7) | 41.5 (37.6–44.5) | 34.8 (33.0–37.0) | −6.9 (−10.5 to −3.0) | −0.3 (−2.3 to +1.3) |
| InternVL3.5 14B | SN | 1072 | −65 | 35.0 (32.8–37.0) | 34.3 (31.9–36.1) | 34.8 (32.8–36.4) | 39.1 (35.4–42.2) | 34.9 (32.7–36.9) | −5.0 (−8.2 to −1.7) | −0.6 (−2.5 to +1.2) |
| InternVL3.5 30B-A3B | SN | 1074 | −104 | 36.4 (34.2–38.6) | 37.5 (34.8–40.1) | 36.7 (34.0–38.9) | 40.5 (37.2–43.8) | 33.9 (31.9–36.5) | −3.0 (−7.7 to +0.9) | +3.7 (−0.1 to +6.1) |
| Pixtral-12B | SN | 1075 | −37 | 30.8 (29.2–32.9) | 31.4 (29.4–33.7) | 32.2 (30.2–34.3) | 36.4 (33.1–40.0) | 31.2 (29.2–34.0) | −5.1 (−8.2 to −1.9) | +0.2 (−1.6 to +1.9) |
| Mistral Small 3.2 24B | SN | 1072 | −116 | 38.2 (36.0–40.6) | 37.5 (36.0–40.0) | 39.6 (36.9–42.3) | 39.2 (36.3–43.4) | 37.0 (34.2–40.2) | −1.6 (−5.9 to +2.2) | +0.4 (−2.9 to +4.8) |

Images with reference energy above 20 kcal. In each of 200 random splits, the calibration constants were determined on one half and all columns evaluated on the other half; entries are the median over splits with the 2.5th and 97.5th percentiles. The last two columns are within-split differences between MedAPE under the JP persona and after subtraction of the mean bias, or division by the median ratio (median and percentiles). From p44ci_medape.csv and p44ci_open2_medape.csv (p44_medape_ci.py, which adds these columns to p44_confirm.py without changing the splits or the estimators). For Gemma 3 4B the estimate is the lower end of a range in most responses.

### **Table S14. Contrasts between conditions by stratum of the error without a persona (chat format)**

Table S14a. JP − US.

| **Model** | **Set** | **n** | **Mean contrast (kcal)** | **log2: < 0** | **0–50** | **50–150** | **> 150** | **Proportion lowered: < 0** | **0–50** | **50–150** | **> 150** | **n per stratum** | **Step (95% interval)** | **Form described** |
| --- | --- | --- | --- | --- | --- | --- | --- | --- | --- | --- | --- | --- | --- | --- |
| Gemini 2.5 Flash | NI | 858 | −61 | −0.10 | −0.38 | −0.36 | −0.40 | 0.54 | 0.69 | 0.70 | 0.79 | 157 / 158 / 249 / 294 | −0.28 | yes |
| Gemini 2.5 Pro | NI | 858 | −42 | −0.19 | −0.40 | −0.36 | −0.39 | 0.62 | 0.74 | 0.74 | 0.81 | 244 / 190 / 239 / 185 | −0.19 | yes |
| Claude Sonnet 4.6 | NI | 857 | −23 | −0.15 | −0.32 | −0.27 | −0.29 | 0.48 | 0.62 | 0.61 | 0.76 | 380 / 220 / 168 / 89 | −0.14 | yes |
| Claude Haiku 4.5 (2025 run) | NI | 855 | −32 | −0.14 | −0.24 | −0.28 | −0.26 | 0.41 | 0.56 | 0.57 | 0.55 | 291 / 132 / 224 / 208 | −0.12 | yes |
| GPT-4o | NI | 849 | 0 | +0.02 | −0.09 | −0.10 | −0.09 | 0.34 | 0.41 | 0.41 | 0.46 | 438 / 188 / 188 / 35 | −0.11 | no |
| GPT-4o-mini | NI | 856 | +5 | +0.05 | −0.06 | −0.10 | −0.02 | 0.21 | 0.32 | 0.39 | 0.32 | 371 / 199 / 223 / 63 | −0.11 | no |
| GPT-5.6 Luna | NI | 852 | −40 | −0.04 | −0.16 | −0.26 | −0.32 | 0.38 | 0.52 | 0.65 | 0.72 | 102 / 96 / 255 / 399 | −0.21 (−0.29 to −0.14) | yes |
| GPT-5.6 Sol | NI | 855 | −39 | −0.14 | −0.38 | −0.37 | −0.30 | 0.58 | 0.75 | 0.77 | 0.77 | 209 / 167 / 286 / 193 | −0.21 (−0.27 to −0.15) | yes |
| Claude Sonnet 5 | NI | 854 | −20 | −0.09 | −0.27 | −0.30 | −0.20 | 0.47 | 0.65 | 0.70 | 0.73 | 302 / 203 / 265 / 84 | −0.17 (−0.21 to −0.12) | yes |
| Gemini 3.1 Pro | NI | 854 | −22 | −0.16 | −0.26 | −0.27 | −0.28 | 0.51 | 0.60 | 0.56 | 0.67 | 380 / 197 / 216 / 61 | −0.11 (−0.18 to −0.03) | yes |
| Gemini 3.8 Flash | NI | 854 | −6 | −0.05 | −0.13 | −0.09 | −0.06 | 0.50 | 0.59 | 0.54 | 0.58 | 464 / 212 / 133 / 45 | −0.04 (−0.08 to 0.00) | no |
| Claude Haiku 4.5 (2026 run) | NI | 852 | −36 | −0.24 | −0.33 | −0.28 | −0.25 | 0.49 | 0.61 | 0.57 | 0.52 | 284 / 132 / 232 / 204 | −0.05 (−0.14 to +0.03) | yes |
| Gemma 3 4B (range responses; reference only) | NI | 855 | −40 | −0.16 | −0.30 | −0.31 | −0.24 | 0.46 | 0.56 | 0.62 | 0.54 | 163 / 101 / 240 / 351 | −0.12 (−0.20 to −0.04) | no |
| Gemma 3 12B | NI | 855 | −46 | −0.20 | −0.28 | −0.41 | −0.29 | 0.47 | 0.51 | 0.64 | 0.65 | 274 / 179 / 259 / 143 | −0.13 (−0.19 to −0.07) | yes |
| Gemma 3 27B | NI | 855 | −55 | −0.30 | −0.42 | −0.46 | −0.36 | 0.60 | 0.67 | 0.70 | 0.72 | 199 / 130 / 272 / 254 | −0.12 (−0.19 to −0.04) | yes |
| Qwen3-VL 2B | NI | 854 | +3 | +0.01 | +0.05 | +0.03 | −0.01 | 0.07 | 0.02 | 0.05 | 0.12 | 341 / 133 / 328 / 52 | +0.01 (−0.02 to +0.03) | no |
| Qwen3-VL 4B | NI | 853 | −13 | −0.11 | −0.13 | −0.14 | −0.06 | 0.29 | 0.28 | 0.35 | 0.28 | 434 / 179 / 204 / 36 | 0.00 (−0.03 to +0.03) | yes |
| Qwen3-VL 8B | NI | 853 | −16 | −0.08 | −0.11 | −0.09 | −0.09 | 0.30 | 0.24 | 0.31 | 0.35 | 259 / 152 / 327 / 115 | −0.01 (−0.05 to +0.02) | yes |
| Qwen3-VL 32B | NI | 854 | −18 | −0.08 | −0.11 | −0.22 | −0.18 | 0.30 | 0.35 | 0.48 | 0.48 | 254 / 162 / 281 / 157 | −0.09 (−0.14 to −0.03) | yes |
| Qwen3-VL 30B-A3B (MoE) | NI | 854 | +13 | +0.06 | +0.02 | +0.05 | +0.07 | 0.10 | 0.16 | 0.13 | 0.06 | 326 / 194 / 236 / 98 | −0.02 (−0.06 to +0.02) | no |
| InternVL3.5 2B | NI | 850 | +4 | +0.01 | +0.01 | +0.03 | +0.08 | 0.06 | 0.03 | 0.03 | 0.00 | 452 / 187 / 178 / 33 | +0.03 (0.00 to +0.06) | no |
| InternVL3.5 4B | NI | 854 | −8 | −0.02 | 0.00 | −0.09 | −0.13 | 0.20 | 0.13 | 0.27 | 0.60 | 546 / 176 / 122 / 10 | −0.03 (−0.07 to +0.02) | no |
| InternVL3.5 8B | NI | 854 | −22 | −0.09 | −0.09 | −0.08 | −0.11 | 0.33 | 0.29 | 0.29 | 0.37 | 275 / 124 / 304 / 151 | −0.01 (−0.04 to +0.03) | yes |
| InternVL3.5 14B | NI | 853 | −8 | −0.06 | −0.03 | −0.04 | −0.06 | 0.21 | 0.16 | 0.17 | 0.29 | 327 / 147 / 267 / 112 | +0.02 (−0.03 to +0.06) | no |
| InternVL3.5 30B-A3B (MoE) | NI | 854 | −12 | −0.10 | −0.15 | −0.18 | −0.19 | 0.18 | 0.24 | 0.30 | 0.37 | 443 / 169 / 188 / 54 | −0.07 (−0.13 to −0.01) | yes |
| Pixtral-12B | NI | 854 | −1 | +0.02 | +0.01 | −0.02 | −0.03 | 0.11 | 0.13 | 0.13 | 0.20 | 270 / 114 / 269 / 201 | −0.03 (−0.07 to 0.00) | no |
| Mistral Small 3.2 24B | NI | 854 | −18 | −0.15 | −0.28 | −0.16 | −0.11 | 0.30 | 0.37 | 0.34 | 0.34 | 456 / 179 / 169 / 50 | −0.04 (−0.09 to +0.01) | yes |
| Gemini 2.5 Flash | SN | 1199 | −39 | −0.14 | −0.15 | −0.22 | −0.13 | 0.56 | 0.61 | 0.65 | 0.61 | 356 / 204 / 252 / 387 | −0.02 | yes |
| Gemini 2.5 Pro | SN | 1202 | −26 | −0.13 | −0.13 | −0.17 | −0.15 | 0.64 | 0.62 | 0.67 | 0.63 | 486 / 266 / 224 / 226 | −0.02 | yes |
| Claude Sonnet 4.6 | SN | 1204 | −18 | −0.12 | −0.13 | −0.12 | −0.11 | 0.44 | 0.43 | 0.43 | 0.41 | 625 / 213 / 218 / 148 | +0.01 | yes |
| Claude Haiku 4.5 (2025 run) | SN | 1185 | −32 | −0.21 | −0.19 | −0.21 | −0.16 | 0.55 | 0.52 | 0.51 | 0.47 | 661 / 182 / 206 / 136 | +0.03 | yes |
| GPT-4o | SN | 1202 | −1 | −0.01 | −0.04 | +0.02 | −0.04 | 0.33 | 0.36 | 0.34 | 0.34 | 783 / 198 / 163 / 58 | −0.02 | no |
| GPT-4o-mini | SN | 1203 | −2 | −0.02 | −0.06 | −0.04 | −0.01 | 0.26 | 0.31 | 0.26 | 0.29 | 708 / 220 / 209 / 66 | −0.02 | no |
| GPT-5.6 Luna | SN | 1075 | −27 | −0.18 | −0.09 | −0.13 | −0.09 | 0.52 | 0.47 | 0.54 | 0.52 | 234 / 154 / 218 / 469 | +0.08 (0.00 to +0.16) | yes |
| GPT-5.6 Sol | SN | 1075 | −22 | −0.15 | −0.12 | −0.11 | −0.10 | 0.62 | 0.57 | 0.53 | 0.60 | 404 / 225 / 230 / 216 | +0.04 (−0.01 to +0.08) | yes |
| Claude Sonnet 5 | SN | 1074 | −15 | −0.11 | −0.12 | −0.12 | −0.13 | 0.45 | 0.41 | 0.48 | 0.48 | 582 / 224 / 187 / 81 | −0.01 (−0.05 to +0.03) | yes |
| Gemini 3.1 Pro | SN | 1072 | −14 | −0.10 | −0.05 | −0.04 | −0.01 | 0.50 | 0.47 | 0.51 | 0.36 | 628 / 211 / 175 / 58 | +0.07 (+0.01 to +0.13) | yes |
| Gemini 3.8 Flash | SN | 1074 | −18 | −0.10 | −0.05 | −0.15 | −0.10 | 0.62 | 0.55 | 0.68 | 0.70 | 654 / 196 / 157 / 67 | 0.00 (−0.04 to +0.05) | yes |
| Claude Haiku 4.5 (2026 run) | SN | 1053 | −35 | −0.24 | −0.24 | −0.20 | −0.23 | 0.52 | 0.56 | 0.55 | 0.56 | 593 / 176 / 168 / 116 | +0.01 (−0.05 to +0.08) | yes |
| Gemma 3 4B (range responses; reference only) | SN | 1075 | −28 | −0.13 | −0.17 | −0.16 | −0.13 | 0.43 | 0.49 | 0.40 | 0.44 | 531 / 152 / 244 / 148 | −0.03 (−0.06 to +0.01) | no |
| Gemma 3 12B | SN | 1073 | −21 | −0.12 | −0.10 | −0.19 | −0.09 | 0.32 | 0.34 | 0.34 | 0.28 | 598 / 180 / 216 / 79 | 0.00 (−0.05 to +0.04) | yes |
| Gemma 3 27B | SN | 1074 | −25 | −0.09 | −0.17 | −0.12 | −0.17 | 0.41 | 0.42 | 0.39 | 0.55 | 487 / 216 / 221 / 150 | −0.06 (−0.11 to −0.02) | yes |
| Qwen3-VL 2B | SN | 1074 | +2 | +0.01 | +0.04 | +0.02 | +0.04 | 0.04 | 0.03 | 0.02 | 0.02 | 602 / 146 / 276 / 50 | +0.02 (0.00 to +0.04) | no |
| Qwen3-VL 4B | SN | 1055 | −13 | −0.08 | −0.19 | −0.16 | −0.06 | 0.24 | 0.35 | 0.30 | 0.26 | 676 / 175 / 165 / 39 | −0.06 (−0.09 to −0.02) | yes |
| Qwen3-VL 8B | SN | 1072 | −6 | −0.02 | +0.01 | 0.00 | −0.03 | 0.16 | 0.12 | 0.14 | 0.20 | 494 / 177 / 289 / 112 | +0.01 (−0.01 to +0.03) | no |
| Qwen3-VL 32B | SN | 1070 | −10 | −0.07 | −0.02 | −0.03 | −0.07 | 0.30 | 0.20 | 0.29 | 0.35 | 520 / 169 / 251 / 130 | +0.03 (0.00 to +0.06) | no |
| Qwen3-VL 30B-A3B (MoE) | SN | 1072 | −2 | −0.01 | 0.00 | −0.02 | +0.01 | 0.25 | 0.26 | 0.29 | 0.25 | 584 / 177 / 214 / 97 | +0.01 (−0.03 to +0.05) | no |
| InternVL3.5 2B | SN | 1068 | +1 | 0.00 | −0.04 | 0.00 | +0.01 | 0.07 | 0.10 | 0.03 | 0.00 | 758 / 165 / 125 / 20 | −0.02 (−0.04 to 0.00) | no |
| InternVL3.5 4B | SN | 1070 | −9 | −0.05 | −0.02 | −0.07 | −0.04 | 0.24 | 0.19 | 0.17 | 0.31 | 815 / 133 / 109 / 13 | +0.01 (−0.04 to +0.05) | no |
| InternVL3.5 8B | SN | 1072 | −16 | −0.07 | −0.04 | −0.07 | −0.06 | 0.37 | 0.17 | 0.31 | 0.32 | 617 / 168 / 234 / 53 | +0.02 (−0.01 to +0.05) | yes |
| InternVL3.5 14B | SN | 1072 | +1 | +0.03 | +0.05 | +0.07 | +0.01 | 0.12 | 0.13 | 0.09 | 0.14 | 605 / 183 / 225 / 59 | +0.01 (−0.03 to +0.06) | no |
| InternVL3.5 30B-A3B (MoE) | SN | 1074 | −8 | −0.08 | −0.10 | −0.04 | −0.01 | 0.18 | 0.18 | 0.19 | 0.16 | 724 / 187 / 132 / 31 | +0.03 (−0.02 to +0.08) | no |
| Pixtral-12B | SN | 1075 | −4 | 0.00 | −0.04 | −0.02 | −0.06 | 0.15 | 0.14 | 0.10 | 0.23 | 551 / 174 / 252 / 98 | −0.03 (−0.07 to 0.00) | no |
| Mistral Small 3.2 24B | SN | 1072 | −13 | −0.09 | −0.18 | −0.17 | −0.06 | 0.28 | 0.43 | 0.35 | 0.27 | 766 / 174 / 110 / 22 | −0.08 (−0.15 to −0.02) | yes |

Table S14b. jp_home − us_home.

| **Model** | **Set** | **n** | **Mean contrast (kcal)** | **log2: < 0** | **0–50** | **50–150** | **> 150** | **Proportion lowered: < 0** | **0–50** | **50–150** | **> 150** | **n per stratum** | **Step (95% interval)** | **Form described** |
| --- | --- | --- | --- | --- | --- | --- | --- | --- | --- | --- | --- | --- | --- | --- |
| Gemini 2.5 Flash | NI | 858 | −57 | −0.15 | −0.41 | −0.34 | −0.34 | 0.55 | 0.71 | 0.71 | 0.77 | 157 / 158 / 249 / 294 | −0.22 | yes |
| Gemini 2.5 Pro | NI | 859 | −32 | −0.20 | −0.39 | −0.34 | −0.27 | 0.58 | 0.76 | 0.72 | 0.67 | 245 / 190 / 239 / 185 | −0.13 | yes |
| Claude Sonnet 4.6 | NI | 857 | −20 | −0.15 | −0.26 | −0.22 | −0.21 | 0.43 | 0.52 | 0.54 | 0.62 | 380 / 220 / 168 / 89 | −0.08 | yes |
| Claude Haiku 4.5 (2025 run) | NI | 854 | −35 | −0.16 | −0.28 | −0.30 | −0.24 | 0.42 | 0.50 | 0.59 | 0.52 | 290 / 132 / 224 / 208 | −0.11 | yes |
| GPT-4o | NI | 846 | −3 | 0.00 | −0.13 | −0.11 | −0.15 | 0.31 | 0.37 | 0.41 | 0.53 | 436 / 188 / 186 / 36 | −0.14 | no |
| GPT-4o-mini | NI | 857 | +2 | +0.04 | −0.07 | −0.06 | +0.02 | 0.23 | 0.30 | 0.30 | 0.17 | 371 / 200 / 223 / 63 | −0.07 | no |
| GPT-5.6 Luna | NI | 851 | −42 | −0.06 | −0.17 | −0.23 | −0.30 | 0.41 | 0.53 | 0.59 | 0.71 | 102 / 95 / 255 / 399 | −0.18 (−0.24 to −0.12) | yes |
| GPT-5.6 Sol | NI | 854 | −35 | −0.10 | −0.26 | −0.30 | −0.24 | 0.53 | 0.68 | 0.75 | 0.76 | 208 / 167 / 286 / 193 | −0.17 (−0.22 to −0.12) | yes |
| Claude Sonnet 5 | NI | 852 | −16 | −0.08 | −0.23 | −0.26 | −0.16 | 0.44 | 0.58 | 0.67 | 0.57 | 301 / 203 / 264 / 84 | −0.13 (−0.18 to −0.09) | yes |
| Gemini 3.1 Pro | NI | 853 | −19 | −0.10 | −0.23 | −0.25 | −0.22 | 0.48 | 0.61 | 0.61 | 0.59 | 380 / 196 / 216 / 61 | −0.14 (−0.21 to −0.06) | yes |
| Gemini 3.8 Flash | NI | 854 | −9 | −0.05 | −0.15 | −0.08 | −0.12 | 0.42 | 0.56 | 0.50 | 0.64 | 464 / 212 / 133 / 45 | −0.07 (−0.12 to −0.02) | no |
| Claude Haiku 4.5 (2026 run) | NI | 853 | −36 | −0.17 | −0.28 | −0.27 | −0.27 | 0.41 | 0.59 | 0.54 | 0.56 | 285 / 132 / 232 / 204 | −0.10 (−0.17 to −0.04) | yes |
| Gemma 3 4B (range responses; reference only) | NI | 855 | −32 | −0.11 | −0.08 | −0.10 | −0.16 | 0.40 | 0.36 | 0.41 | 0.53 | 163 / 101 / 240 / 351 | 0.00 (−0.05 to +0.05) | no |
| Gemma 3 12B | NI | 855 | −44 | −0.17 | −0.19 | −0.32 | −0.29 | 0.39 | 0.40 | 0.53 | 0.64 | 274 / 179 / 259 / 143 | −0.09 (−0.15 to −0.04) | yes |
| Gemma 3 27B | NI | 855 | −50 | −0.24 | −0.32 | −0.38 | −0.29 | 0.53 | 0.62 | 0.73 | 0.70 | 199 / 130 / 272 / 254 | −0.09 (−0.14 to −0.03) | yes |
| Qwen3-VL 2B | NI | 854 | −4 | −0.02 | 0.00 | −0.03 | −0.06 | 0.11 | 0.06 | 0.12 | 0.23 | 341 / 133 / 328 / 52 | −0.01 (−0.03 to +0.01) | no |
| Qwen3-VL 4B | NI | 853 | −19 | −0.14 | −0.17 | −0.18 | −0.13 | 0.34 | 0.35 | 0.40 | 0.44 | 434 / 179 / 204 / 36 | −0.02 (−0.06 to +0.01) | yes |
| Qwen3-VL 8B | NI | 853 | −18 | −0.12 | −0.11 | −0.10 | −0.10 | 0.36 | 0.25 | 0.32 | 0.36 | 259 / 152 / 327 / 115 | +0.01 (−0.02 to +0.05) | yes |
| Qwen3-VL 32B | NI | 854 | −22 | −0.08 | −0.14 | −0.24 | −0.21 | 0.20 | 0.30 | 0.48 | 0.44 | 254 / 162 / 281 / 157 | −0.12 (−0.16 to −0.06) | yes |
| Qwen3-VL 30B-A3B (MoE) | NI | 854 | −4 | −0.04 | −0.16 | −0.06 | −0.01 | 0.23 | 0.36 | 0.26 | 0.15 | 326 / 194 / 236 / 98 | −0.04 (−0.07 to 0.00) | no |
| InternVL3.5 2B | NI | 850 | 0 | −0.01 | −0.01 | −0.01 | +0.04 | 0.06 | 0.06 | 0.06 | 0.03 | 452 / 187 / 178 / 33 | +0.02 (0.00 to +0.05) | no |
| InternVL3.5 4B | NI | 854 | −10 | −0.06 | −0.04 | −0.08 | −0.15 | 0.20 | 0.15 | 0.25 | 0.70 | 546 / 176 / 122 / 10 | −0.01 (−0.04 to +0.03) | yes |
| InternVL3.5 8B | NI | 854 | −20 | −0.07 | −0.09 | −0.09 | −0.11 | 0.27 | 0.28 | 0.27 | 0.36 | 275 / 124 / 304 / 151 | −0.02 (−0.05 to +0.01) | yes |
| InternVL3.5 14B | NI | 854 | −5 | −0.05 | −0.03 | +0.01 | −0.05 | 0.16 | 0.15 | 0.12 | 0.28 | 328 / 147 / 267 / 112 | +0.03 (−0.01 to +0.07) | no |
| InternVL3.5 30B-A3B (MoE) | NI | 854 | −18 | −0.15 | −0.18 | −0.10 | −0.19 | 0.29 | 0.27 | 0.27 | 0.39 | 443 / 169 / 188 / 54 | −0.01 (−0.06 to +0.04) | yes |
| Pixtral-12B | NI | 854 | −17 | −0.07 | −0.07 | −0.11 | −0.10 | 0.26 | 0.25 | 0.29 | 0.34 | 270 / 114 / 269 / 201 | −0.02 (−0.05 to +0.01) | yes |
| Mistral Small 3.2 24B | NI | 854 | −15 | −0.09 | −0.16 | −0.09 | −0.14 | 0.25 | 0.26 | 0.24 | 0.42 | 456 / 179 / 169 / 50 | −0.04 (−0.08 to 0.00) | yes |
| Gemini 2.5 Flash | SN | 1197 | −22 | −0.08 | −0.08 | −0.14 | −0.12 | 0.55 | 0.50 | 0.60 | 0.56 | 355 / 204 / 252 / 386 | −0.03 | yes |
| Gemini 2.5 Pro | SN | 1201 | −20 | −0.14 | −0.17 | −0.13 | −0.10 | 0.58 | 0.56 | 0.58 | 0.53 | 485 / 266 / 224 / 226 | 0.00 | yes |
| Claude Sonnet 4.6 | SN | 1204 | −8 | −0.09 | −0.08 | −0.06 | −0.05 | 0.35 | 0.33 | 0.28 | 0.33 | 625 / 213 / 218 / 148 | +0.03 | no |
| Claude Haiku 4.5 (2025 run) | SN | 1192 | −29 | −0.18 | −0.13 | −0.18 | −0.17 | 0.47 | 0.36 | 0.51 | 0.43 | 666 / 183 / 207 / 136 | +0.02 | yes |
| GPT-4o | SN | 1202 | −2 | 0.00 | +0.01 | 0.00 | +0.03 | 0.33 | 0.33 | 0.36 | 0.43 | 783 / 198 / 163 / 58 | +0.01 | no |
| GPT-4o-mini | SN | 1202 | −1 | −0.01 | +0.02 | −0.03 | −0.01 | 0.26 | 0.29 | 0.24 | 0.23 | 708 / 219 / 209 / 66 | 0.00 | no |
| GPT-5.6 Luna | SN | 1074 | −35 | −0.15 | −0.19 | −0.11 | −0.13 | 0.52 | 0.48 | 0.48 | 0.57 | 234 / 153 / 218 / 469 | 0.00 (−0.06 to +0.07) | yes |
| GPT-5.6 Sol | SN | 1074 | −22 | −0.12 | −0.14 | −0.06 | −0.10 | 0.55 | 0.57 | 0.51 | 0.58 | 404 / 225 / 229 / 216 | +0.02 (−0.02 to +0.06) | yes |
| Claude Sonnet 5 | SN | 1074 | −14 | −0.12 | −0.12 | −0.10 | −0.16 | 0.46 | 0.47 | 0.45 | 0.48 | 583 / 224 / 186 / 81 | 0.00 (−0.05 to +0.04) | yes |
| Gemini 3.1 Pro | SN | 1071 | −18 | −0.12 | −0.03 | −0.02 | −0.03 | 0.57 | 0.43 | 0.50 | 0.46 | 626 / 211 / 175 / 59 | +0.10 (+0.04 to +0.14) | yes |
| Gemini 3.8 Flash | SN | 1074 | −14 | −0.09 | −0.02 | −0.11 | −0.12 | 0.56 | 0.48 | 0.60 | 0.49 | 654 / 196 / 157 / 67 | 0.00 (−0.04 to +0.05) | yes |
| Claude Haiku 4.5 (2026 run) | SN | 1060 | −30 | −0.23 | −0.17 | −0.14 | −0.16 | 0.51 | 0.42 | 0.43 | 0.47 | 598 / 178 / 168 / 116 | +0.07 (+0.01 to +0.14) | yes |
| Gemma 3 4B (range responses; reference only) | SN | 1075 | −30 | −0.13 | −0.11 | −0.09 | −0.15 | 0.44 | 0.41 | 0.39 | 0.57 | 531 / 152 / 244 / 148 | +0.01 (−0.02 to +0.05) | no |
| Gemma 3 12B | SN | 1073 | −26 | −0.12 | −0.12 | −0.22 | −0.14 | 0.32 | 0.29 | 0.38 | 0.43 | 598 / 180 / 216 / 79 | −0.04 (−0.08 to −0.01) | yes |
| Gemma 3 27B | SN | 1074 | −34 | −0.14 | −0.15 | −0.18 | −0.19 | 0.53 | 0.49 | 0.59 | 0.71 | 487 / 216 / 221 / 150 | −0.03 (−0.06 to 0.00) | yes |
| Qwen3-VL 2B | SN | 1074 | −4 | −0.03 | −0.02 | −0.02 | −0.01 | 0.11 | 0.08 | 0.06 | 0.08 | 602 / 146 / 276 / 50 | +0.01 (−0.01 to +0.03) | no |
| Qwen3-VL 4B | SN | 1059 | −16 | −0.11 | −0.13 | −0.12 | −0.13 | 0.34 | 0.29 | 0.30 | 0.38 | 676 / 176 / 168 / 39 | −0.02 (−0.06 to +0.01) | yes |
| Qwen3-VL 8B | SN | 1072 | −13 | −0.04 | −0.04 | −0.05 | −0.07 | 0.23 | 0.16 | 0.21 | 0.33 | 494 / 177 / 289 / 112 | −0.01 (−0.03 to +0.01) | yes |
| Qwen3-VL 32B | SN | 1072 | −14 | −0.08 | −0.07 | −0.06 | −0.10 | 0.23 | 0.21 | 0.24 | 0.32 | 522 / 169 / 251 / 130 | +0.01 (−0.02 to +0.04) | yes |
| Qwen3-VL 30B-A3B (MoE) | SN | 1072 | −10 | −0.04 | −0.08 | −0.06 | −0.04 | 0.30 | 0.30 | 0.31 | 0.30 | 584 / 177 / 214 / 97 | −0.02 (−0.06 to +0.02) | no |
| InternVL3.5 2B | SN | 1067 | 0 | −0.01 | −0.03 | −0.01 | 0.00 | 0.07 | 0.08 | 0.04 | 0.00 | 757 / 165 / 125 / 20 | −0.01 (−0.03 to +0.01) | no |
| InternVL3.5 4B | SN | 1070 | −9 | −0.04 | −0.03 | −0.04 | −0.08 | 0.24 | 0.20 | 0.14 | 0.38 | 815 / 133 / 109 / 13 | 0.00 (−0.04 to +0.06) | no |
| InternVL3.5 8B | SN | 1072 | −13 | −0.06 | −0.04 | −0.09 | −0.05 | 0.27 | 0.15 | 0.29 | 0.30 | 617 / 168 / 234 / 53 | 0.00 (−0.03 to +0.03) | yes |
| InternVL3.5 14B | SN | 1072 | 0 | +0.02 | +0.04 | +0.08 | −0.02 | 0.10 | 0.11 | 0.09 | 0.14 | 605 / 183 / 225 / 59 | +0.02 (−0.02 to +0.06) | no |
| InternVL3.5 30B-A3B (MoE) | SN | 1074 | −15 | −0.10 | −0.11 | −0.09 | −0.10 | 0.30 | 0.25 | 0.24 | 0.26 | 724 / 187 / 132 / 31 | 0.00 (−0.05 to +0.04) | yes |
| Pixtral-12B | SN | 1075 | −13 | −0.07 | −0.09 | −0.06 | −0.09 | 0.25 | 0.20 | 0.20 | 0.32 | 551 / 174 / 252 / 98 | −0.01 (−0.04 to +0.02) | yes |
| Mistral Small 3.2 24B | SN | 1073 | −12 | −0.08 | −0.15 | −0.12 | −0.05 | 0.23 | 0.33 | 0.25 | 0.18 | 767 / 174 / 110 / 22 | −0.06 (−0.10 to −0.01) | yes |

Table S14c. smaller − US.

| **Model** | **Set** | **n** | **Mean contrast (kcal)** | **log2: < 0** | **0–50** | **50–150** | **> 150** | **Proportion lowered: < 0** | **0–50** | **50–150** | **> 150** | **n per stratum** | **Step (95% interval)** | **Form described** |
| --- | --- | --- | --- | --- | --- | --- | --- | --- | --- | --- | --- | --- | --- | --- |
| Gemini 2.5 Flash | NI | 858 | −62 | −0.23 | −0.34 | −0.26 | −0.32 | 0.69 | 0.69 | 0.68 | 0.74 | 157 / 158 / 249 / 294 | −0.08 | yes |
| Gemini 2.5 Pro | NI | 858 | −125 | −0.78 | −0.80 | −0.77 | −0.83 | 0.97 | 0.97 | 0.99 | 0.98 | 244 / 190 / 239 / 185 | −0.02 | yes |
| Claude Sonnet 4.6 | NI | 857 | −44 | −0.30 | −0.35 | −0.33 | −0.39 | 0.73 | 0.74 | 0.71 | 0.82 | 380 / 220 / 168 / 89 | −0.06 | yes |
| Claude Haiku 4.5 (2025 run) | NI | 856 | −49 | −0.26 | −0.30 | −0.32 | −0.32 | 0.55 | 0.59 | 0.67 | 0.65 | 292 / 132 / 224 / 208 | −0.05 | yes |
| GPT-4o | NI | 850 | −7 | −0.05 | +0.02 | −0.05 | −0.03 | 0.37 | 0.30 | 0.33 | 0.53 | 438 / 187 / 189 / 36 | +0.03 | no |
| GPT-4o-mini | NI | 857 | −11 | −0.07 | −0.07 | −0.12 | −0.10 | 0.36 | 0.33 | 0.42 | 0.41 | 371 / 200 / 223 / 63 | −0.03 | yes |
| GPT-5.6 Luna | NI | 852 | +35 | +0.16 | +0.22 | +0.21 | +0.13 | 0.15 | 0.10 | 0.10 | 0.16 | 102 / 96 / 255 / 399 | +0.03 (−0.07 to +0.11) | no |
| GPT-5.6 Sol | NI | 855 | −21 | −0.09 | −0.14 | −0.10 | −0.11 | 0.61 | 0.58 | 0.58 | 0.65 | 209 / 167 / 286 / 193 | −0.02 (−0.06 to +0.02) | yes |
| Claude Sonnet 5 | NI | 854 | −76 | −0.51 | −0.54 | −0.55 | −0.55 | 0.95 | 0.97 | 0.96 | 0.99 | 302 / 203 / 265 / 84 | −0.03 (−0.06 to 0.00) | yes |
| Gemini 3.1 Pro | NI | 854 | −105 | −0.81 | −0.85 | −0.91 | −0.86 | 0.96 | 0.95 | 0.98 | 1.00 | 380 / 197 / 216 / 61 | −0.06 (−0.12 to 0.00) | yes |
| Gemini 3.8 Flash | NI | 854 | −65 | −0.49 | −0.48 | −0.52 | −0.50 | 0.95 | 0.97 | 0.98 | 0.98 | 464 / 212 / 133 / 45 | −0.01 (−0.05 to +0.03) | yes |
| Claude Haiku 4.5 (2026 run) | NI | 853 | −52 | −0.32 | −0.33 | −0.30 | −0.35 | 0.63 | 0.64 | 0.62 | 0.65 | 285 / 132 / 232 / 204 | −0.01 (−0.07 to +0.06) | yes |
| Gemma 3 4B (range responses; reference only) | NI | 855 | −25 | −0.16 | −0.12 | −0.09 | −0.09 | 0.49 | 0.45 | 0.42 | 0.42 | 163 / 101 / 240 / 351 | +0.06 (−0.01 to +0.13) | no |
| Gemma 3 12B | NI | 855 | −57 | −0.31 | −0.26 | −0.33 | −0.36 | 0.62 | 0.47 | 0.58 | 0.79 | 274 / 179 / 259 / 143 | 0.00 (−0.05 to +0.05) | yes |
| Gemma 3 27B | NI | 855 | −72 | −0.37 | −0.25 | −0.27 | −0.38 | 0.84 | 0.62 | 0.69 | 0.90 | 199 / 130 / 272 / 254 | +0.07 (+0.03 to +0.12) | yes |
| Qwen3-VL 2B | NI | 854 | −14 | −0.09 | −0.04 | −0.05 | −0.14 | 0.26 | 0.16 | 0.20 | 0.42 | 341 / 133 / 328 / 52 | +0.01 (−0.01 to +0.04) | yes |
| Qwen3-VL 4B | NI | 853 | −18 | −0.11 | −0.08 | −0.12 | −0.09 | 0.37 | 0.28 | 0.35 | 0.42 | 434 / 179 / 204 / 36 | +0.02 (−0.01 to +0.04) | yes |
| Qwen3-VL 8B | NI | 853 | −5 | −0.02 | +0.02 | +0.01 | −0.02 | 0.19 | 0.14 | 0.13 | 0.17 | 259 / 152 / 327 / 115 | +0.03 (−0.01 to +0.06) | no |
| Qwen3-VL 32B | NI | 854 | −38 | −0.18 | −0.12 | −0.19 | −0.25 | 0.54 | 0.42 | 0.54 | 0.76 | 254 / 162 / 281 / 157 | −0.01 (−0.05 to +0.04) | yes |
| Qwen3-VL 30B-A3B (MoE) | NI | 854 | −6 | −0.01 | 0.00 | −0.04 | −0.04 | 0.18 | 0.18 | 0.25 | 0.26 | 326 / 194 / 236 / 98 | −0.02 (−0.05 to +0.01) | no |
| InternVL3.5 2B | NI | 850 | −11 | −0.10 | −0.14 | −0.09 | −0.06 | 0.22 | 0.22 | 0.16 | 0.21 | 452 / 187 / 178 / 33 | 0.00 (−0.04 to +0.03) | yes |
| InternVL3.5 4B | NI | 854 | −12 | −0.08 | −0.01 | −0.11 | −0.11 | 0.28 | 0.15 | 0.34 | 0.50 | 546 / 176 / 122 / 10 | +0.02 (−0.03 to +0.07) | yes |
| InternVL3.5 8B | NI | 854 | −9 | −0.04 | −0.03 | +0.02 | −0.03 | 0.23 | 0.15 | 0.10 | 0.19 | 275 / 124 / 304 / 151 | +0.02 (0.00 to +0.05) | no |
| InternVL3.5 14B | NI | 853 | −15 | −0.07 | −0.05 | −0.10 | −0.10 | 0.29 | 0.22 | 0.30 | 0.44 | 327 / 147 / 267 / 112 | −0.02 (−0.05 to +0.01) | yes |
| InternVL3.5 30B-A3B (MoE) | NI | 854 | −4 | 0.00 | −0.02 | −0.02 | −0.03 | 0.10 | 0.11 | 0.14 | 0.15 | 443 / 169 / 188 / 54 | −0.02 (−0.06 to +0.03) | no |
| Pixtral-12B | NI | 854 | −31 | −0.14 | −0.09 | −0.14 | −0.19 | 0.49 | 0.32 | 0.42 | 0.62 | 270 / 114 / 269 / 201 | 0.00 (−0.04 to +0.05) | yes |
| Mistral Small 3.2 24B | NI | 854 | −32 | −0.21 | −0.17 | −0.21 | −0.28 | 0.47 | 0.34 | 0.49 | 0.76 | 456 / 179 / 169 / 50 | −0.01 (−0.05 to +0.03) | yes |
| Gemini 2.5 Flash | SN | 1200 | −75 | −0.23 | −0.27 | −0.36 | −0.31 | 0.65 | 0.65 | 0.76 | 0.76 | 357 / 204 / 252 / 387 | −0.09 | yes |
| Gemini 2.5 Pro | SN | 1202 | −148 | −0.77 | −0.77 | −0.80 | −0.87 | 0.98 | 0.98 | 0.99 | 1.00 | 486 / 266 / 224 / 226 | −0.04 | yes |
| Claude Sonnet 4.6 | SN | 1204 | −7 | −0.04 | −0.01 | +0.03 | −0.08 | 0.37 | 0.31 | 0.30 | 0.40 | 625 / 213 / 218 / 148 | +0.02 | no |
| Claude Haiku 4.5 (2025 run) | SN | 1193 | −47 | −0.23 | −0.32 | −0.32 | −0.21 | 0.63 | 0.58 | 0.63 | 0.60 | 666 / 185 / 206 / 136 | −0.06 | yes |
| GPT-4o | SN | 1202 | −13 | −0.08 | −0.09 | −0.07 | −0.07 | 0.42 | 0.38 | 0.38 | 0.48 | 783 / 198 / 163 / 58 | 0.00 | yes |
| GPT-4o-mini | SN | 1203 | −16 | −0.08 | −0.08 | −0.09 | −0.07 | 0.40 | 0.36 | 0.42 | 0.36 | 708 / 220 / 209 / 66 | 0.00 | yes |
| GPT-5.6 Luna | SN | 1074 | +49 | +0.18 | +0.25 | +0.14 | +0.17 | 0.11 | 0.10 | 0.11 | 0.09 | 233 / 154 / 218 / 469 | 0.00 (−0.05 to +0.06) | no |
| GPT-5.6 Sol | SN | 1075 | −20 | −0.10 | −0.10 | −0.10 | −0.06 | 0.59 | 0.55 | 0.60 | 0.58 | 404 / 225 / 230 / 216 | +0.01 (−0.02 to +0.04) | yes |
| Claude Sonnet 5 | SN | 1075 | −91 | −0.52 | −0.56 | −0.56 | −0.65 | 0.96 | 0.95 | 0.97 | 1.00 | 583 / 224 / 187 / 81 | −0.06 (−0.10 to −0.02) | yes |
| Gemini 3.1 Pro | SN | 1072 | −127 | −0.82 | −0.85 | −0.84 | −1.01 | 0.96 | 0.99 | 0.97 | 0.98 | 628 / 211 / 175 / 58 | −0.07 (−0.16 to 0.00) | yes |
| Gemini 3.8 Flash | SN | 1074 | −84 | −0.47 | −0.46 | −0.49 | −0.56 | 0.96 | 0.97 | 0.98 | 0.96 | 654 / 196 / 157 / 67 | −0.03 (−0.07 to 0.00) | yes |
| Claude Haiku 4.5 (2026 run) | SN | 1059 | −45 | −0.23 | −0.30 | −0.27 | −0.23 | 0.59 | 0.66 | 0.60 | 0.61 | 597 / 178 / 168 / 116 | −0.04 (−0.10 to +0.02) | yes |
| Gemma 3 4B (range responses; reference only) | SN | 1075 | −2 | −0.01 | +0.02 | +0.02 | +0.07 | 0.33 | 0.26 | 0.24 | 0.31 | 531 / 152 / 244 / 148 | +0.04 (0.00 to +0.08) | no |
| Gemma 3 12B | SN | 1073 | −56 | −0.32 | −0.29 | −0.36 | −0.32 | 0.65 | 0.56 | 0.63 | 0.73 | 598 / 180 / 216 / 79 | −0.01 (−0.05 to +0.04) | yes |
| Gemma 3 27B | SN | 1074 | −59 | −0.24 | −0.22 | −0.26 | −0.31 | 0.74 | 0.65 | 0.71 | 0.85 | 487 / 216 / 221 / 150 | −0.03 (−0.07 to +0.01) | yes |
| Qwen3-VL 2B | SN | 1074 | −16 | −0.10 | −0.03 | −0.05 | −0.05 | 0.33 | 0.23 | 0.20 | 0.20 | 602 / 146 / 276 / 50 | +0.06 (+0.03 to +0.08) | yes |
| Qwen3-VL 4B | SN | 1058 | −18 | −0.08 | −0.14 | −0.12 | −0.09 | 0.34 | 0.35 | 0.34 | 0.31 | 677 / 176 / 166 / 39 | −0.03 (−0.07 to +0.01) | yes |
| Qwen3-VL 8B | SN | 1072 | −6 | −0.02 | 0.00 | −0.01 | −0.03 | 0.16 | 0.14 | 0.13 | 0.18 | 494 / 177 / 289 / 112 | +0.01 (−0.01 to +0.03) | no |
| Qwen3-VL 32B | SN | 1071 | −46 | −0.21 | −0.16 | −0.19 | −0.28 | 0.73 | 0.53 | 0.71 | 0.75 | 521 / 169 / 251 / 130 | 0.00 (−0.04 to +0.04) | yes |
| Qwen3-VL 30B-A3B (MoE) | SN | 1073 | −8 | −0.01 | +0.02 | −0.02 | −0.03 | 0.30 | 0.28 | 0.23 | 0.34 | 585 / 177 / 214 / 97 | −0.01 (−0.05 to +0.04) | no |
| InternVL3.5 2B | SN | 1068 | −13 | −0.10 | −0.10 | −0.15 | −0.12 | 0.28 | 0.20 | 0.24 | 0.40 | 758 / 165 / 125 / 20 | −0.02 (−0.05 to +0.02) | yes |
| InternVL3.5 4B | SN | 1070 | −16 | −0.10 | −0.04 | −0.07 | −0.08 | 0.34 | 0.28 | 0.21 | 0.54 | 815 / 133 / 109 / 13 | +0.04 (−0.01 to +0.09) | yes |
| InternVL3.5 8B | SN | 1073 | −8 | −0.03 | −0.02 | +0.04 | +0.01 | 0.23 | 0.12 | 0.14 | 0.11 | 617 / 169 / 234 / 53 | +0.04 (0.00 to +0.07) | no |
| InternVL3.5 14B | SN | 1072 | −16 | −0.09 | −0.01 | −0.10 | −0.07 | 0.36 | 0.25 | 0.32 | 0.39 | 605 / 183 / 225 / 59 | +0.02 (−0.02 to +0.07) | yes |
| InternVL3.5 30B-A3B (MoE) | SN | 1074 | −5 | −0.02 | −0.03 | 0.00 | −0.09 | 0.13 | 0.12 | 0.13 | 0.26 | 724 / 187 / 132 / 31 | −0.02 (−0.06 to +0.02) | no |
| Pixtral-12B | SN | 1075 | −35 | −0.13 | −0.13 | −0.17 | −0.20 | 0.54 | 0.41 | 0.50 | 0.63 | 551 / 174 / 252 / 98 | −0.03 (−0.09 to +0.02) | yes |
| Mistral Small 3.2 24B | SN | 1073 | −31 | −0.15 | −0.15 | −0.24 | −0.20 | 0.53 | 0.53 | 0.61 | 0.73 | 767 / 174 / 110 / 22 | −0.05 (−0.11 to +0.02) | yes |

Strata: error of the estimate without a persona (estimate − reference), < 0, 0–50, 50–150, > 150 kcal. log2: mean of log2 of the ratio of the two estimates. Step: mean of the three strata with positive error minus the stratum with negative error (strata with fewer than 30 images not used). Form described: mean contrast −10 kcal or beyond. Intervals: percentile, 2,000 resamples of images; none for the six models of the previous study. From p43_contrast.csv, p44_cells.csv, and p44open2_cells.csv.

### **Table S1. Accuracy and shift by condition, all 54 cells (energy)**

Table S1a. MedAPE (%) by condition and Spearman's ρ without a persona.

| **Model** | **Set** | **ρ none** | **none** | **jp** | **us** | **jp_home** | **us_home** | **neutral** | **smaller** | **larger** |
| --- | --- | --- | --- | --- | --- | --- | --- | --- | --- | --- |
| Qwen3-VL 2B | NI | 0.71 | 44.1 | 42.6 | — | 42.5 | — | — | — | — |
| Qwen3-VL 2B | SN | 0.73 | 39.5 | 37.4 | — | — | — | — | — | — |
| Qwen3-VL 4B | NI | 0.77 | 38.1 | 37.8 | 38.0 | 37.9 | 37.4 | 38.6 | 38.1 | 37.2 |
| Qwen3-VL 4B | SN | 0.77 | 36.0 | 36.2 | 34.9 | 39.2 | 36.9 | 35.1 | 35.9 | 34.7 |
| Qwen3-VL 8B | NI | 0.77 | 46.6 | 42.6 | 45.9 | 42.2 | 44.7 | 44.2 | 44.9 | 59.9 |
| Qwen3-VL 8B | SN | 0.78 | 35.0 | 34.1 | 34.8 | 31.8 | 33.7 | 34.3 | 33.6 | 36.7 |
| Qwen3-VL 32B | NI | 0.75 | 48.6 | 39.2 | 44.4 | 36.9 | 45.7 | 46.5 | 40.5 | 79.4 |
| Qwen3-VL 32B | SN | 0.77 | 32.5 | 32.4 | 30.9 | 31.7 | 30.9 | 31.6 | 33.4 | 41.9 |
| Qwen3-VL 30B-A3B | NI | 0.78 | 36.5 | 39.0 | 37.4 | 39.8 | 40.9 | 37.1 | 35.7 | 39.8 |
| Qwen3-VL 30B-A3B | SN | 0.79 | 31.3 | 32.4 | 32.1 | 32.9 | 32.5 | 32.9 | 31.3 | 31.9 |
| Gemma 3 4B | NI | 0.62 | 109.7 | 47.9 | 60.9 | 59.2 | 63.1 | 49.1 | 53.9 | 64.7 |
| Gemma 3 4B | SN | 0.74 | 39.0 | 34.7 | 34.5 | 35.2 | 35.2 | 34.4 | 35.5 | 34.8 |
| Gemma 3 12B | NI | 0.73 | 42.5 | 35.8 | 44.2 | 39.0 | 47.4 | 44.3 | 39.6 | 71.8 |
| Gemma 3 12B | SN | 0.78 | 34.0 | 33.5 | 31.7 | 34.1 | 33.5 | 31.9 | 35.9 | 36.1 |
| Gemma 3 27B | NI | 0.73 | 57.4 | 40.2 | 56.6 | 44.0 | 63.2 | 53.5 | 42.3 | 135.4 |
| Gemma 3 27B | SN | 0.79 | 32.5 | 31.7 | 32.0 | 31.2 | 32.0 | 32.0 | 32.3 | 47.8 |
| InternVL3.5 2B | NI | 0.66 | 42.9 | 44.0 | — | 43.9 | 43.0 | 43.1 | 42.9 | 42.8 |
| InternVL3.5 2B | SN | 0.73 | 41.3 | 40.2 | 41.1 | 41.8 | 41.8 | — | — | — |
| InternVL3.5 4B | NI | 0.71 | 41.3 | 41.9 | 40.9 | 40.8 | 40.1 | 40.8 | 42.3 | 40.9 |
| InternVL3.5 4B | SN | 0.73 | 44.0 | 43.0 | 41.7 | 43.1 | 42.6 | 42.0 | 44.6 | 42.6 |
| InternVL3.5 8B | NI | 0.69 | 50.1 | 48.3 | 52.9 | 47.8 | 52.9 | 48.4 | 50.4 | 54.1 |
| InternVL3.5 8B | SN | 0.74 | 37.0 | 36.0 | 35.1 | 35.9 | 35.3 | 35.9 | 35.1 | 35.9 |
| InternVL3.5 14B | NI | 0.68 | 47.2 | 47.2 | 47.4 | 47.1 | 45.0 | 45.5 | 44.9 | 48.1 |
| InternVL3.5 14B | SN | 0.76 | 36.6 | 36.0 | 35.7 | 36.1 | 36.1 | 36.0 | 36.0 | 36.2 |
| InternVL3.5 30B-A3B | NI | 0.71 | 39.4 | 41.6 | 40.7 | 40.4 | 39.0 | 40.1 | 40.5 | 39.3 |
| InternVL3.5 30B-A3B | SN | 0.76 | 37.5 | 38.5 | 37.5 | 38.2 | 35.6 | 37.5 | 37.8 | 36.4 |
| Mistral Pixtral-12B | NI | 0.66 | 50.4 | 48.8 | 48.5 | 50.4 | 55.8 | 55.0 | 44.1 | 79.0 |
| Mistral Pixtral-12B | SN | 0.78 | 32.8 | 32.9 | 33.2 | 32.7 | 33.1 | 32.8 | 34.1 | 35.1 |
| Mistral Small-3.2-24B | NI | 0.71 | 39.8 | 40.1 | 41.1 | 40.5 | 40.7 | 42.2 | 40.8 | 43.3 |
| Mistral Small-3.2-24B | SN | 0.77 | 40.1 | 38.5 | 38.7 | 38.6 | 37.8 | 40.1 | 41.2 | 36.6 |
| Haiku 4.5 | NI | 0.60 | 50.4 | 45.5 | 51.7 | 43.9 | 51.5 | 49.1 | 46.5 | 105.1 |
| Haiku 4.5 | SN | 0.72 | 35.6 | 38.9 | 36.9 | 39.8 | 34.8 | — | 38.0 | 45.2 |
| Sonnet 5 | NI | 0.79 | 40.8 | 32.1 | 38.0 | — | 38.0 | 37.5 | 37.0 | 76.7 |
| Sonnet 5 | SN | 0.80 | 30.1 | 30.9 | 30.2 | — | 30.3 | 30.2 | 40.8 | 46.1 |
| GPT-5.6 Luna | NI | 0.72 | 97.2 | 51.4 | 73.3 | 62.0 | 85.9 | 78.9 | 94.0 | 84.1 |
| GPT-5.6 Luna | SN | 0.79 | 50.3 | 42.9 | 46.1 | 43.4 | 48.0 | 48.9 | 59.5 | 49.7 |
| GPT-5.6 Sol | NI | 0.77 | 51.3 | 35.1 | 48.5 | 37.9 | 51.5 | 46.8 | 42.8 | 83.1 |
| GPT-5.6 Sol | SN | 0.80 | 33.2 | 31.9 | 32.2 | 33.2 | 32.6 | 31.8 | 31.5 | 43.9 |
| Gemini 3.8 Flash | NI | 0.86 | 28.5 | 26.6 | 28.4 | 27.9 | 30.0 | 28.3 | 34.0 | 65.5 |
| Gemini 3.8 Flash | SN | 0.81 | 28.9 | 31.2 | 28.3 | 31.2 | 28.3 | 29.2 | 38.7 | 42.3 |
| Gemini 3.1 Pro | NI | 0.80 | 34.7 | 29.4 | 35.2 | 28.4 | 34.2 | 30.8 | 40.9 | 167.2 |
| Gemini 3.1 Pro | SN | 0.81 | 30.7 | 31.0 | — | 32.0 | 27.4 | 29.1 | 50.5 | 102.6 |
| Claude Haiku 4.5 (2025 run) | NI | 0.60 | 50.8 | 43.7 | 51.9 | 42.5 | 50.4 | 47.3 | 43.5 | 105.7 |
| Claude Haiku 4.5 (2025 run) | SN | 0.72 | 36.2 | 39.0 | 36.2 | 38.8 | 35.9 | 36.2 | 38.3 | 45.2 |
| Claude Sonnet 4.6 (2025 run) | NI | 0.78 | 36.2 | 33.3 | 36.5 | 33.9 | 37.3 | 34.7 | 35.6 | 60.6 |
| Claude Sonnet 4.6 (2025 run) | SN | 0.79 | 31.9 | 32.8 | 31.0 | 32.6 | 32.1 | 31.9 | 33.4 | 37.8 |
| GPT-4o | NI | 0.78 | 37.0 | 32.1 | 36.4 | 34.0 | 36.0 | 37.0 | 36.6 | 36.2 |
| GPT-4o | SN | 0.79 | 34.3 | 34.1 | 33.0 | 33.7 | 33.4 | 32.8 | 35.8 | 33.5 |
| GPT-4o-mini | NI | 0.76 | 38.6 | 35.0 | 37.4 | 35.7 | 38.2 | 36.0 | 37.0 | 39.1 |
| GPT-4o-mini | SN | 0.79 | 34.1 | 34.3 | 33.2 | 34.0 | 32.9 | 33.1 | 35.0 | 32.9 |
| Gemini 2.5 Flash | NI | 0.78 | 57.8 | 38.0 | 60.1 | 40.4 | 64.5 | 55.7 | 41.1 | 133.0 |
| Gemini 2.5 Flash | SN | 0.76 | 39.7 | 34.2 | 37.3 | 36.3 | 37.7 | 37.7 | 32.5 | 70.3 |
| Gemini 2.5 Pro | NI | 0.82 | 40.9 | 29.7 | 41.4 | 33.8 | 43.4 | 40.4 | 34.7 | 197.2 |
| Gemini 2.5 Pro | SN | 0.80 | 28.4 | 30.3 | 28.4 | 31.0 | 28.7 | 27.8 | 43.0 | 103.7 |

Table S1b. Mean paired difference from the none condition (kcal) by condition.

| **Model** | **Set** | **Baseline bias** | **jp** | **us** | **jp_home** | **us_home** | **neutral** | **smaller** | **larger** | **Wilcoxon p, jp** |
| --- | --- | --- | --- | --- | --- | --- | --- | --- | --- | --- |
| Qwen3-VL 2B | NI | +2 | +1 | −1 | −1 | +2 | +4 | −15 | −11 | 0.097 |
| Qwen3-VL 2B | SN | −64 | −1 | −3 | −6 | −2 | +3 | −19 | −15 | 0.232 |
| Qwen3-VL 4B | NI | −20 | −3 | +10 | −19 | −1 | 0 | −7 | +15 | 0.019 |
| Qwen3-VL 4B | SN | −77 | −2 | +11 | −23 | −8 | +3 | −6 | +12 | 0.186 |
| Qwen3-VL 8B | NI | +36 | −11 | +5 | −12 | +5 | −3 | 0 | +44 | <0.001 |
| Qwen3-VL 8B | SN | −16 | +8 | +13 | −9 | +4 | +6 | +8 | +40 | <0.001 |
| Qwen3-VL 32B | NI | +44 | −26 | −8 | −31 | −9 | −3 | −45 | +76 | <0.001 |
| Qwen3-VL 32B | SN | −17 | −14 | −5 | −21 | −8 | −1 | −49 | +71 | <0.001 |
| Qwen3-VL 30B-A3B | NI | +17 | +12 | −1 | +11 | +15 | +2 | −6 | +10 | <0.001 |
| Qwen3-VL 30B-A3B | SN | −32 | −6 | −4 | −3 | +6 | +1 | −12 | +8 | <0.001 |
| Gemma 3 4B | NI | +177 | −116 | −75 | −99 | −81 | −117 | −100 | −72 | <0.001 |
| Gemma 3 4B | SN | +20 | −54 | −26 | −48 | −29 | −52 | −27 | −23 | <0.001 |
| Gemma 3 12B | NI | +38 | −35 | +11 | −16 | +28 | +15 | −44 | +90 | <0.001 |
| Gemma 3 12B | SN | −45 | −6 | +14 | −3 | +22 | +11 | −41 | +85 | <0.001 |
| Gemma 3 27B | NI | +88 | −50 | +5 | −34 | +15 | −7 | −66 | +146 | <0.001 |
| Gemma 3 27B | SN | −5 | −25 | −1 | −28 | +4 | −10 | −57 | +115 | <0.001 |
| InternVL3.5 2B | NI | −42 | +8 | +4 | +7 | +7 | +4 | −6 | −3 | <0.001 |
| InternVL3.5 2B | SN | −112 | +7 | +6 | +1 | +1 | +5 | −8 | −4 | <0.001 |
| InternVL3.5 4B | NI | −64 | 0 | +7 | +3 | +13 | +4 | −4 | +5 | 0.787 |
| InternVL3.5 4B | SN | −120 | +1 | +10 | −2 | +7 | +5 | −5 | +5 | 0.097 |
| InternVL3.5 8B | NI | +37 | −7 | +14 | −4 | +15 | −5 | +5 | +20 | <0.001 |
| InternVL3.5 8B | SN | −61 | −7 | +8 | −7 | +5 | −1 | +1 | +15 | <0.001 |
| InternVL3.5 14B | NI | +14 | −3 | +5 | −2 | +3 | −1 | −10 | +8 | 0.009 |
| InternVL3.5 14B | SN | −58 | +2 | +1 | +2 | +3 | 0 | −14 | +5 | 0.003 |
| InternVL3.5 30B-A3B | NI | −24 | −17 | −5 | −5 | +12 | −2 | −9 | +2 | <0.001 |
| InternVL3.5 30B-A3B | SN | −97 | −9 | −1 | −2 | +12 | 0 | −6 | +9 | <0.001 |
| Mistral Pixtral-12B | NI | +47 | −1 | 0 | +1 | +18 | +12 | −30 | +57 | 0.572 |
| Mistral Pixtral-12B | SN | −31 | −9 | −5 | −11 | +2 | +5 | −38 | +47 | <0.001 |
| Mistral Small-3.2-24B | NI | −28 | +9 | +28 | +13 | +28 | +3 | −4 | +49 | <0.001 |
| Mistral Small-3.2-24B | SN | −108 | +8 | +21 | +8 | +19 | +2 | −8 | +36 | <0.001 |
| Haiku 4.5 | NI | +44 | −28 | +8 | −31 | +4 | −8 | −43 | +120 | <0.001 |
| Haiku 4.5 | SN | −38 | −38 | −4 | −32 | −3 | −9 | −46 | +111 | <0.001 |
| Sonnet 5 | NI | +21 | −26 | −6 | −24 | −8 | −5 | −80 | +104 | <0.001 |
| Sonnet 5 | SN | −37 | −22 | −8 | −28 | −14 | −7 | −94 | +118 | <0.001 |
| GPT-5.6 Luna | NI | +149 | −72 | −32 | −56 | −14 | −20 | +2 | −13 | <0.001 |
| GPT-5.6 Luna | SN | +130 | −44 | −19 | −41 | −8 | −11 | +28 | −8 | <0.001 |
| GPT-5.6 Sol | NI | +70 | −39 | 0 | −26 | +9 | −4 | −21 | +76 | <0.001 |
| GPT-5.6 Sol | SN | +32 | −22 | −1 | −10 | +12 | −2 | −20 | +73 | <0.001 |
| Gemini 3.8 Flash | NI | −13 | −3 | +3 | +3 | +11 | +1 | −60 | +127 | <0.001 |
| Gemini 3.8 Flash | SN | −46 | −17 | −1 | −11 | +2 | −3 | −80 | +140 | <0.001 |
| Gemini 3.1 Pro | NI | +3 | −11 | +11 | −3 | +16 | +4 | −92 | +311 | <0.001 |
| Gemini 3.1 Pro | SN | −52 | −2 | +12 | +15 | +33 | +9 | −110 | +334 | 0.029 |
| Claude Haiku 4.5 (2025 run) | NI | +46 | −26 | +5 | −32 | +3 | −11 | −43 | +118 | <0.001 |
| Claude Haiku 4.5 (2025 run) | SN | −38 | −35 | −4 | −29 | 0 | −9 | −46 | +113 | <0.001 |
| Claude Sonnet 4.6 (2025 run) | NI | +6 | −22 | +1 | −17 | +3 | −4 | −42 | +93 | <0.001 |
| Claude Sonnet 4.6 (2025 run) | SN | −19 | −16 | +1 | −16 | −9 | −5 | −6 | +75 | <0.001 |
| GPT-4o | NI | −22 | −2 | −2 | −3 | +1 | +6 | −9 | +12 | 0.071 |
| GPT-4o | SN | −74 | −2 | −1 | −1 | +1 | +6 | −13 | +12 | 0.206 |
| GPT-4o-mini | NI | 0 | −5 | −10 | −2 | −4 | −3 | −21 | +13 | <0.001 |
| GPT-4o-mini | SN | −62 | −8 | −6 | −5 | −3 | −2 | −22 | +13 | <0.001 |
| Gemini 2.5 Flash | NI | +119 | −60 | −1 | −48 | +7 | −6 | −61 | +150 | <0.001 |
| Gemini 2.5 Flash | SN | +84 | −36 | 0 | −26 | −2 | +3 | −70 | +151 | <0.001 |
| Gemini 2.5 Pro | NI | +63 | −40 | +2 | −31 | +1 | −7 | −120 | +298 | <0.001 |
| Gemini 2.5 Pro | SN | +18 | −35 | −10 | −24 | −6 | −11 | −150 | +274 | <0.001 |

Dashes: condition not run (Gemini 2.5 Pro, NutriImage: no smaller/larger run in the previous study). Wilcoxon p: two-sided signed-rank test of jp against none, for reference only (Section 2.5). The smaller and larger shifts of the 2025 group were recomputed from the per-image outputs of the previous study on the same image sets as the other columns (fill_master2.py).
