## Supplementary material for "Effects of Dish Name, Visible Amount, Persona, and Explicit Instruction on Energy Estimates From Food Photographs by 26 Vision–Language Models: Comparative Evaluation Study": ultimedia Appendix 2. Prompt texts

Nakagawa S, Yamamoto A.

### **Multimedia Appendix 2. Prompt texts**

*A.1 Chat format (2.3a).* Task prompt, placed after the image:

Look at this food image. Estimate the nutritional content based solely on visual appearance—color, shape, ingredients, and portion size. Do NOT read any text, labels, or numbers visible in the image. Use only visual information. Provide a brief one-line explanation of your reasoning. Respond ONLY in JSON format: {energy_kcal: value, protein_g: value, fat_g: value, carb_g: value, salt_g: value, basis: one-line reasoning}

| **Condition** | **Description placed before the task prompt** |
| --- | --- |
| none | (none) |
| jp | You are a Japanese registered dietitian (kanri-eiyoshi). You are deeply familiar with standard single-serving portion sizes of everyday Japanese meals. |
| us | You are a United States registered dietitian. You are deeply familiar with standard single-serving portion sizes of everyday American meals. |
| jp_home | You are a Japanese homemaker who prepares family meals every day. You are familiar with the usual serving sizes of everyday Japanese home cooking, but you have no formal nutrition training. |
| us_home | You are an American homemaker who prepares family meals every day. You are familiar with the usual serving sizes of everyday American home cooking, but you have no formal nutrition training. |
| neutral_dietitian | You are a registered dietitian. You have professional knowledge of food composition and nutrition, with no particular national or cultural specialization. |
| smaller | You tend to assume smaller single-serving portion sizes than usual when estimating meals. |
| larger | You tend to assume larger single-serving portion sizes than usual when estimating meals. |

*A.2 Crop and name-only experiments (2.3b, c).* Claude Sonnet 4.6 (claude-sonnet-4-6), max_tokens 400, temperature not specified; the image (JPEG, base64) and the user prompt in one user turn; the persona, when used, as the system prompt (none: no system prompt):

| **Condition** | **System prompt** |
| --- | --- |
| jp | You are a Japanese registered dietitian (管理栄養士) with extensive knowledge of traditional and modern Japanese cuisine, ingredients, and cooking methods. You are highly familiar with Japanese ingredients including various types of dashi, mirin, sake, soy sauce, and traditional vegetables and fish. |
| us | You are an American registered dietitian with expertise in Western cuisine and nutrition. You are familiar with common American and European food ingredients and cooking methods. |

User prompt with an image (cut_estimate.py; line breaks as in the script; the bracketed line was present only in the one-serving conditions, followed by a blank line):

Look at this food image.

Estimate the nutritional content of this meal based solely on its visual appearance —

color, shape, ingredients, and portion size.

Do NOT read any text, labels, or numbers visible in the image.

Use only visual information.

[one-serving line, when used: Assume this is one standard serving.]

Please also provide a brief one-line explanation of your reasoning.

Respond ONLY in JSON format (no other text):

{"dish_name": "name of the dish", "energy_kcal": value, "protein_g": value, "fat_g": value, "carb_g": value, "salt_g": value, "basis": "one-line reasoning"}

User prompt with the dish name only ({name_en} is the English dish name of the reference table):

A dish called "{name_en}" is served.

[one-serving line, when used: Assume this is one standard serving.]

Estimate its nutritional content.

Please also provide a brief one-line explanation of your reasoning.

Respond ONLY in JSON format (no other text):

{"dish_name": "name of the dish", "energy_kcal": value, "protein_g": value, "fat_g": value, "carb_g": value, "salt_g": value, "basis": "one-line reasoning"}

Judge prompt for the criterion of the main text (main ingredient and cooking method). Claude Haiku 4.5 (claude-haiku-4-5-20251001), max_tokens 10, one user turn, no system prompt; {a} is the name returned for the full image and {b} the name returned for the crop, in the same condition. The same prompt is used by cut_judge_names.py (no persona; 60%, 40%, and 25% crops; output cut_judge_none.jsonl and cut_judge_one.jsonl) and by cut_judge_names_persona.py (jp and us; 25% crop; output cut_judge_loose_none_jp.jsonl and cut_judge_loose_none_us.jsonl, whose file names carry 'loose' from the script they were derived from although the criterion is this one):

Two descriptions of a dish are given. Decide whether they refer to the same dish.

A: {a}

B: {b}

Judge by the MAIN INGREDIENT and the COOKING METHOD only.

Answer SAME if the main ingredient and the cooking method match, even when the

side ingredients, garnishes, sauces, or level of detail differ. Examples:

"Braised Chicken Wings with Peas and Onions" vs "Braised Chicken with Peas and Cabbage" = SAME

"Pan-seared fish fillet with cream sauce" vs "Fish fillet" = SAME

"Stir-fried Chicken with garlic shoots" vs "Stir-fried Chicken with Green Peppers" = SAME

Answer DIFFERENT only when the main ingredient or the cooking method differs. Examples:

"Scrambled Eggs" vs "Potato Chips" = DIFFERENT

"Stir-fried Chicken" vs "Stir-fried Pork" = DIFFERENT

"Grilled Fish" vs "Simmered Fish" = DIFFERENT

Respond with exactly one word: SAME or DIFFERENT

Judge prompt for the looser criterion (main ingredient only; Multimedia Appendix 1, S10; cut_judge_loose.py, same model and settings, no persona, three crops; output cut_judge_loose_none.jsonl). The prompt is reproduced as run, including the second paragraph, which was carried over from the prompt above and still mentions the cooking method:

Two descriptions of a dish are given. Decide whether they refer to the same dish.

A: {a}

B: {b}

Judge by the MAIN INGREDIENT only. Ignore differences in cooking method.

Answer SAME if the main ingredient and the cooking method match, even when the

side ingredients, garnishes, sauces, or level of detail differ. Examples:

"Braised Chicken Wings with Peas and Onions" vs "Braised Chicken with Peas and Cabbage" = SAME

"Pan-seared fish fillet with cream sauce" vs "Fish fillet" = SAME

"Stir-fried Chicken with garlic shoots" vs "Stir-fried Chicken with Green Peppers" = SAME

Answer DIFFERENT only when the main ingredient differs. Examples:

"Scrambled Eggs" vs "Potato Chips" = DIFFERENT

"Stir-fried Chicken" vs "Stir-fried Pork" = DIFFERENT

"Grilled Fish" vs "Simmered Fish" = SAME (same ingredient, method ignored)

Respond with exactly one word: SAME or DIFFERENT

Judge prompt for the comparison of names across conditions (none vs jp and none vs us; full image and 25% crop; Section 2.3(b); name_agreement.py, of which cut25_name_agreement.py is the earlier jp-only form with the same prompt; same model and settings; output cut_judge_nonevsjp_full.jsonl, cut_judge_nonevsjp_cut25.jsonl, cut_judge_nonevsus_full.jsonl, cut_judge_nonevsus_cut25.jsonl). {a} is the name returned without a persona and {b} the name returned under the persona. The criterion is that of the main text; unlike the prompt above, this one gives no examples and lists language among the differences to be ignored:

Two descriptions of a dish are given. One may be in Japanese. Decide whether they refer to the same dish.

A: {a}

B: {b}

Judge by the MAIN INGREDIENT and the COOKING METHOD only. Answer SAME if the main ingredient and the cooking method match, even when side ingredients, garnishes, sauces, language, or level of detail differ. Answer DIFFERENT only when the main ingredient or the cooking method differs.

Respond with exactly one word: SAME or DIFFERENT

*A.3 Narrator format (2.3d).* Document: narrator sentence, line break, image token, line break, 'Energy (kcal):'. Candidates: 50 to 1,500 kcal in steps of 50 (30 values), each scored as ' {value}' followed by a line break.

| **Condition** | **First sentence(s) of the document** |
| --- | --- |
| none | Estimated nutritional content of the dish in the photo. |
| jp | A Japanese registered dietitian (管理栄養士), familiar with standard single-serving portion sizes of everyday Japanese meals, looked at the photo and estimated its nutritional content. |
| us | An American registered dietitian, familiar with standard single-serving portion sizes of everyday meals in the United States, looked at the photo and estimated its nutritional content. |
| smaller | Estimated nutritional content of the dish in the photo. The portion is smaller than it looks. |
| larger | Estimated nutritional content of the dish in the photo. The portion is larger than it looks. |
