## Supplementary material for "Effects of Dish Name, Visible Amount, Persona, and Explicit Instruction on Energy Estimates From Food Photographs by 26 Vision–Language Models: Comparative Evaluation Study": Multimedia Appendix 3. Description of the reproducibility package

Nakagawa S, Yamamoto A.

**Multimedia Appendix 3. Description of the reproducibility package**

The reproducibility package is deposited on Zenodo (doi:10.5281/zenodo.22917651) under the authors' account. It contains the files from which every table, figure, and number of the main text and of Multimedia Appendix 1 can be regenerated. The NutriImage images themselves are not included (restricted access; Data Availability Statement); SNAPMe and ACETADA images are obtained from their providers.

### **1. Per-cell tables**

master_cells_energy_v2_20260921.csv — the 54 cells (model × image set) with baseline bias, MedAPE, ρ, the eight condition shifts, JP − US, larger − smaller, and the Wilcoxon p-values (Tables 1, 4, S1; Figures 2, 3). Derived tables: table_input_ablation_sonnet_v2.csv (Table 5), acetada_before_after_5models.csv (Table 6), gemma_ptit_ni_summary.csv, gemma_ptit_snapme_summary.csv, gemma_ptit_snapme_mode_27B.csv, internvl_ptit_ni_summary.csv (Table 7, S8), ptit_paired_diff.csv, p43_contrast.csv, p44ci_medape.csv, p44ci_open2_medape.csv (Tables 8, 9, S13, S14), s2_bf16_fp8.csv (S2), mac_4bit_cells_doc01.csv (S3), grid50_none_by_cell.csv (S6a), error_stratified_independent_v2.csv (S8b), s9_pairs_per_model.csv and ni_size_pairs.csv (S9), s10_names_v2.csv, s10_name_stability_v2.csv, s10_retention_x_names_v2.csv (S10), p42b_conditions.csv, p42b_shift_repro.csv, p42b_form.csv (S12), basis_jp_rate_run1_run2.csv (Section 4.6), nutriimage_dims.csv and snapme_dims.csv (Section 2.2).

### **2. Per-image outputs**

Chat format: the per-image outputs of the 26 models (image, condition, parsed estimates, basis) — one JSON-lines file per model and image set for the models run for this article, with both runs of the six commercial models of 2026, and one JSON file per model, image set, and condition for the six commercial models of the previous study (NutriImage restricted to the 887 images of version 1.1); the Qwen3-VL runs in FP8 and in MLX 4-bit (Multimedia Appendix 1, S2, S3); and the before- and after-meal photographs of ACETADA with Qwen3-VL 8B, Gemma 3 27B, and Claude Sonnet 4.6 (Table 6). Likelihood scoring: the candidate log-probabilities of Gemma 3 (pt, it; 4B, 12B, 27B) for NutriImage and SNAPMe and of Gemma 3 27B (pt, it) for ACETADA, and of InternVL3.5 8B (pre-trained, final) for NutriImage and SNAPMe; the SNAPMe files of InternVL3.5 were run as planned (Section 5) but are not analysed in this article. Crop and name-only experiments: the per-image results of Claude Sonnet 4.6 (full, 60%, 40%, 25% crop, name only; none, jp, us; one-serving variants) and the judge files (cut_judge_none.jsonl, cut_judge_one.jsonl, cut_judge_loose_none.jsonl, cut_judge_loose_none_jp.jsonl, cut_judge_loose_none_us.jsonl, cut_judge_nonevsjp_{full,cut25}.jsonl, cut_judge_nonevsus_{full,cut25}.jsonl). Human ratings: judge_validation_100_final.csv, nonevsjp_validation_100_SN.xlsx, nonevsjp_100_human_categories.csv, nonevsjp_100_shift_by_group.csv.

### **3. Prompts and run configuration**

The prompt texts are reproduced in Multimedia Appendix 2 and are included as the scripts that issued them (commercial_run.py, cut_estimate.py, cut_judge_names.py, cut_judge_names_persona.py, cut_judge_loose.py, name_agreement.py, the vLLM and Transformers scoring scripts of the persona:v5–v11 images, pilot_p23_local_v4.py for MLX). Model identifiers, precision, runtime versions, and dates are in Table 3.

### **4. Aggregation and figure scripts**

rebuild_master.py, fill_master.py, fill_master2.py (master table); sonnet_recompute.py (Table 5, Figure 3A–B, S8b, S10); s1_s5.py, s2_fp8.py, iv_summary.py, paired_ptit.py, p44_medape_ci.py, p42c_run1_vs_run2.py, acetada_basis.py, basis_jp_rate.py, image_dims.py; fig1.py, fig2.py, fig3.py, fig3e.py (Figures 2–4); the docx builders (mk25t.js, sup39.js, build_submit.py) with the patch scripts recording every revision.

### **5. Dated records of expectations**

The expectations written before the runs of Sections 2.3(c)–(e) (Gemma 3 pt/it scoring, InternVL3.5 pre-trained/final, the repeated run of the six commercial models) are included as dated Markdown files (internvl_ptit_runplan_20260921.md, internvl_step3_runplan_20260922.md, repeat_run_predictions_20260921.md), unchanged after the runs.

### **6. ACETADA served energy**

acetada_basis.py reconstructs the served and remaining energy from the consumed-amount labels of ACETADA (Section 2.2; Table 6); the reconstruction is the authors' calculation.
